# Scaling genetic discovery for organ volumes using machine learning-assisted imputation and bias-corrected GWAS

**DOI:** 10.1101/2025.11.07.25339752

**Authors:** Afreen Naz, Brandon Whitcher, Altayeb Ahmed, Marjola Thanaj, Elena P Sorokin, Jimmy D Bell, E Louise Thomas, Madeleine Cule, Hanieh Yaghootkar

**Author notes:** **Corresponding authors:** Hanieh Yaghootkar.

## Abstract

**Background:** MRI-derived organ and tissue volumes are powerful endophenotypes for studying complex disease, but their availability is limited by cost and throughput. We present a scalable framework that combines machine learning-based phenotypic imputation with probabilistic GWAS (POP-GWAS) to enable robust genetic discovery for imaging-derived phenotypes (IDPs).

**Results:** Using 37,589 UK Biobank MRI scans and 382 biomarkers, we imputed nine IDPs—including volumes of fat depots, muscle, pancreas, and lung—across ∼450,000 individuals. The POP-GWAS framework integrated measured and imputed traits, correcting for imputation uncertainty and increasing effective sample size by up to 200%. We identified 452 independent loci associated with the nine IDPs.

This approach uncovered new insights into the architecture and disease relevance of organ volumes. For example, genetically higher abdominal subcutaneous fat was associated with higher risks of diabetes, polycystic ovary syndrome, cardiovascular disease, gout, osteoarthritis, asthma, psoriasis; higher visceral fat with cholelithiasis and reflux; higher muscle volume with aortic aneurysm, atrial fibrillation, thrombotic events, osteoarthritis, but a lower risk of depression; higher lung volume with higher risks of aortic aneurysm, but a lower risk of heart disease and reflux; higher pancreas volume with lower risk of diabetes. Tissue enrichment analyses revealed organ-specific patterns, e.g., brain tissue for fat traits and pancreatic for pancreas volume.

**Conclusions:** Our study demonstrates that machine learning-assisted GWAS enables scalable discovery in imaging genetics. This framework advances understanding of organ-specific biology and provides a blueprint for leveraging the remaining >60,000 UK Biobank MRI scans to accelerate genetic discovery and uncover mechanisms of disease.

## Introduction

Genome-wide association studies (GWAS) have transformed our understanding of the genetic architecture of complex traits and diseases by identifying thousands of trait-associated loci. However, the power of GWAS is fundamentally limited by the availability and quality of phenotypic data. While routine measurements such as body mass index (BMI) and blood biomarkers are widely available, they often provide only coarse-grained representations of biological processes. In contrast, imaging-derived phenotypes (IDPs), particularly organ and tissue volumes derived from magnetic resonance imaging (MRI), offer high-resolution insights into human biology but remain underutilised due to their cost, complexity, and limited availability at scale.

Organ and tissue volumes—such as those of the liver, pancreas, lungs, kidneys, muscle, and adipose tissue—are clinically and biologically important endophenotypes [1]. Their variation has been implicated in the pathogenesis of a wide range of diseases, including type 2 diabetes, cardiovascular disease, chronic kidney disease, respiratory conditions, and several cancers [2]. For instance, increased visceral fat is associated with a higher risk of diabetes and cardiovascular disease, while changes in kidney volume have been connected to chronic kidney disease and hypertension [3, 4]. However, the high cost of MRI scanning limits the availability of these phenotypes in biobanks, constraining statistical power and reducing the ability to uncover novel biological mechanisms through GWAS [5].

To overcome the limitations of sparse imaging phenotypes, we developed a scalable framework that combines machine learning-based phenotypic imputation with a probabilistic genome-wide association approach (POP-GWAS) [6]. This strategy enables robust discovery from traits that are only available in a subset of individuals, by imputing high-cost phenotypes using routine biomarkers and correcting for uncertainty in downstream genetic analyses [6].

In this study, we combine machine learning-based phenotypic imputation with POP-GWAS to perform the largest GWAS to date of MRI-derived organ and tissue volumes in the UK Biobank. Using 37,589 MRI scans and 382 routine biomarkers, we imputed nine IDPs—including volumes of abdominal subcutaneous and visceral fat, muscle, pancreas, kidneys, spleen, lungs, and liver—across approximately 450,000 participants. We then applied POP-GWAS to integrate directly measured and imputed traits, improving power by up to 200% and enabling robust genetic discovery. We report 452 independent loci associated with organ and tissue volumes, many of which are novel, and demonstrate concordance between imputed and measured effect sizes. Our results uncover new biological insights into tissue-specific genetic regulation and highlight disease-relevant pathways. This approach unlocks the potential of imaging genetics at population scale, enabling deeper insights into the molecular underpinnings of organ morphology, function, and disease susceptibility.

## Results

### Study population

The UK Biobank imaging cohort included 42,963 participants with abdominal MRI scans. Of these, 37,705 individualswith high-quality organ and tissue volume measurements were designated as the imaging cohort for genetic analyses and model training. An additional 5,258 participants were held out as a test set for independent validation of the imputation models and were not included in GWAS analyses. The remaining 370,568 participants without MRI scans comprised the non-imaging cohort, in which IDPs were imputed using machine learning models trained in the imaging cohort (**Supplementary Figure 1)**.

Each MRI scan provided nine IDPs representing volumes of abdominal subcutaneous fat, visceral fat, iliopsoas muscle (left and right), total muscle, liver, kidneys (left and right), lungs, pancreas, and spleen. Detailed demographic and clinical characteristics for the imaging and non-imaging cohorts are summarised in **Supplementary Table 1**.

### Prediction of IDPs

To extend genetic discovery beyond participants with imaging data, we developed machine learning models to impute nine IDPs in 370,568 UK Biobank participants without MRI scans. Imputation models were trained using 37,705 participants with high-quality MRI-derived organ volume measurements, and validated in an independent test set of 5,258 individuals.

Imputation was based on 382 routinely collected biomarkers spanning anthropometric traits, clinical biochemistry, blood counts, and lifestyle variables. These features were available across the full cohort. Trait-specific feature selection using the Boruta algorithm was applied to remove redundant or uninformative predictors. The number of retained features varied across IDPs, ranging from 69 (spleen) to 101 (abdominal subcutaneous fat), reflecting the biological complexity and predictability of each trait. A detailed breakdown of the UK Biobank field IDs used for each IDP—including anthropometric, biochemical, haematological, lifestyle, and impedance variables—is provided in **Supplementary Table 2**. Fat and muscle depots relied more heavily on anthropometric and impedance-derived measures, while internal organs (e.g., liver, kidney, pancreas) included a greater proportion of biochemical markers.

Out-of-sample validation in the test cohort (N = 5,258) demonstrated strong concordance between imputed and directly measured MRI traits. Prediction accuracy varied by organ, with the strongest performance observed for abdominal subcutaneous fat (R² = 0.90) and total muscle volume (R² = 0.93), followed by visceral fat, iliopsoas muscle, and liver volume (R² > 0.60). Smaller and morphologically variable organs such as the pancreas and spleen were more challenging to predict (R² ∼0.19–0.47), though imputation accuracy remained sufficient for downstream genetic analysis. Full performance metrics and R² values across the four evaluated modelling approaches are reported in **Supplementary Table 3** and **Supplementary Figure 2**.

The consistency of correlation patterns between imputed and measured IDPs, and the preservation of their multivariate structure, support the robustness of the imputation framework and its suitability for large-scale genetic discovery.

### Genetic architecture of organ and tissue volume measurements

To maximise discovery power while correcting for bias introduced by machine learning-based imputation, we applied POP-GWAS—a three-stage framework that uses the directly measured imaging cohort to calibrate association statistics from the much larger imputed dataset. This enabled robust and well-calibrated GWAS across all nine imputed IDPs.

We performed GWAS in 370,568 non-imaged UK Biobank participants using imputed IDPs and then corrected SNP effect sizes using bias estimates derived from the imaging cohort. This strategy substantially increased effective sample sizes—by up to 200%—relative to the original imaging subset. To avoid circularity and double-counting, we used the imaging cohort only for estimating the imputation bias in POP-GWAS and not for meta-analysis or joint association testing. The corrected GWAS results from the non-imaging cohort (with imputed IDPs) are treated as the primary discovery set and used for all downstream analyses.

Abdominal subcutaneous fat and total muscle volume showed the greatest gains (effective n = 112,710 and 98,463, respectively), while pancreas volume had the smallest increase (effective n = 37,607) (**Supplementary table 4**). SNP-based heritability estimates of the imputed traits ranged from 0.17 for pancreas volume to 0.30 for iliopsoas muscle volume (**Supplementary table 4**). We observed elevated genomic inflation factors (λ) for several traits, including abdominal subcutaneous fat (λ = 1.58), which may reflect both true polygenicity and residual confounding. However, the LDSC intercepts were close to 1, indicating that the majority of the observed inflation was attributable to polygenic signal rather than confounding (**Supplementary table 4**). Nevertheless, we recommend interpreting GWAS results in the context of effect size distributions and replication across traits.

In total, we identified 452 independent genome-wide significant loci (P < 5 × 10□□) associated with the nine IDPs (**Figure 1; Supplementary Figures 3; Supplementary Table 5**). These included 65 loci for abdominal subcutaneous fat, 32 for visceral fat, 25 for iliopsoas muscle, 116 for total muscle, 47 for kidney, 34 for liver, 29 for lung, 28 for pancreas, and 72 for spleen volume. Internal validation using directly measured traits in the imaging cohort revealed that the majority of loci (>90%) replicated at nominal significance with concordant direction of effect, indicating high reliability of the imputation-based GWAS (**Supplementary table 5, Supplementary figure 4**).

**Figure 1.**
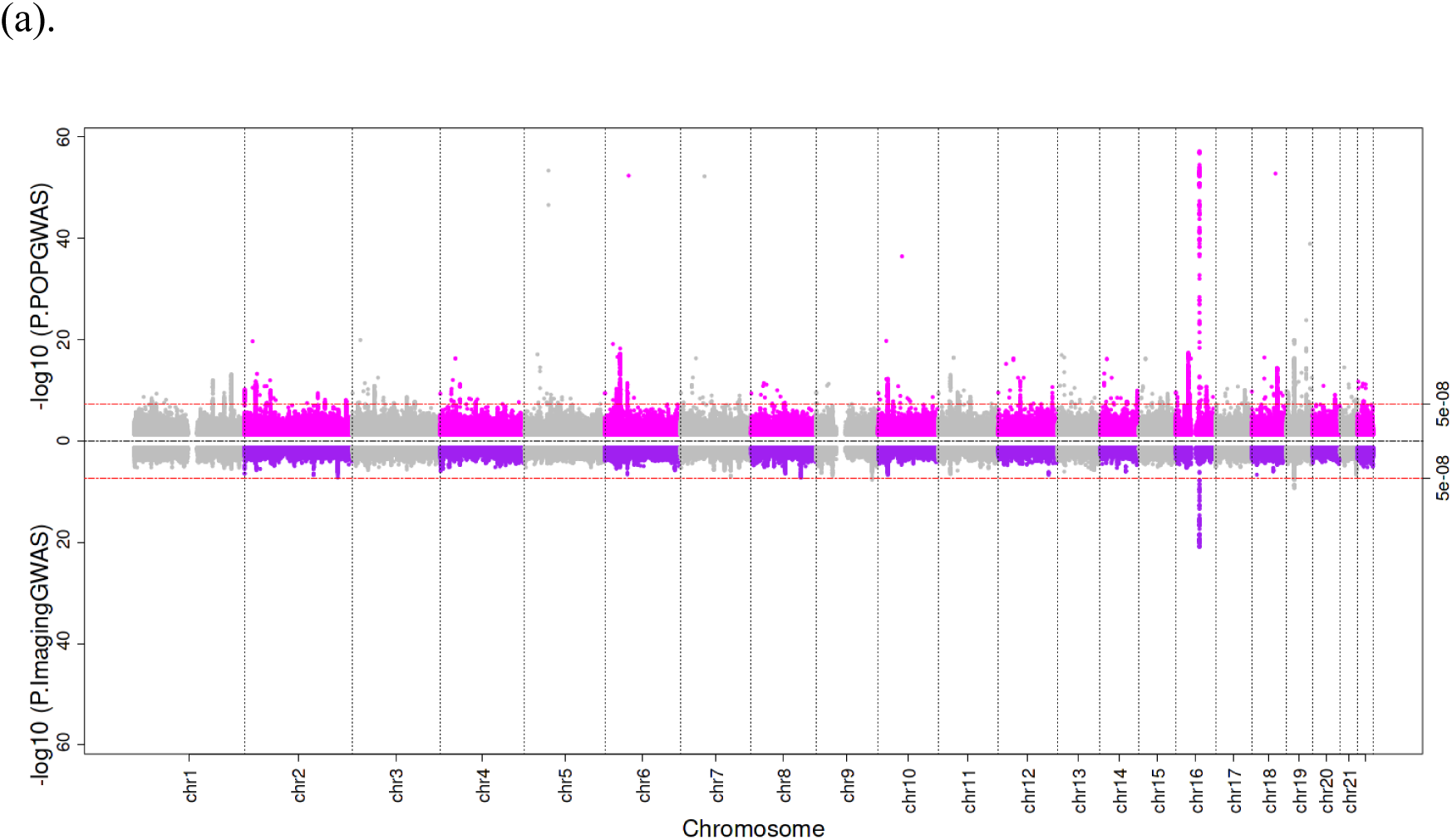
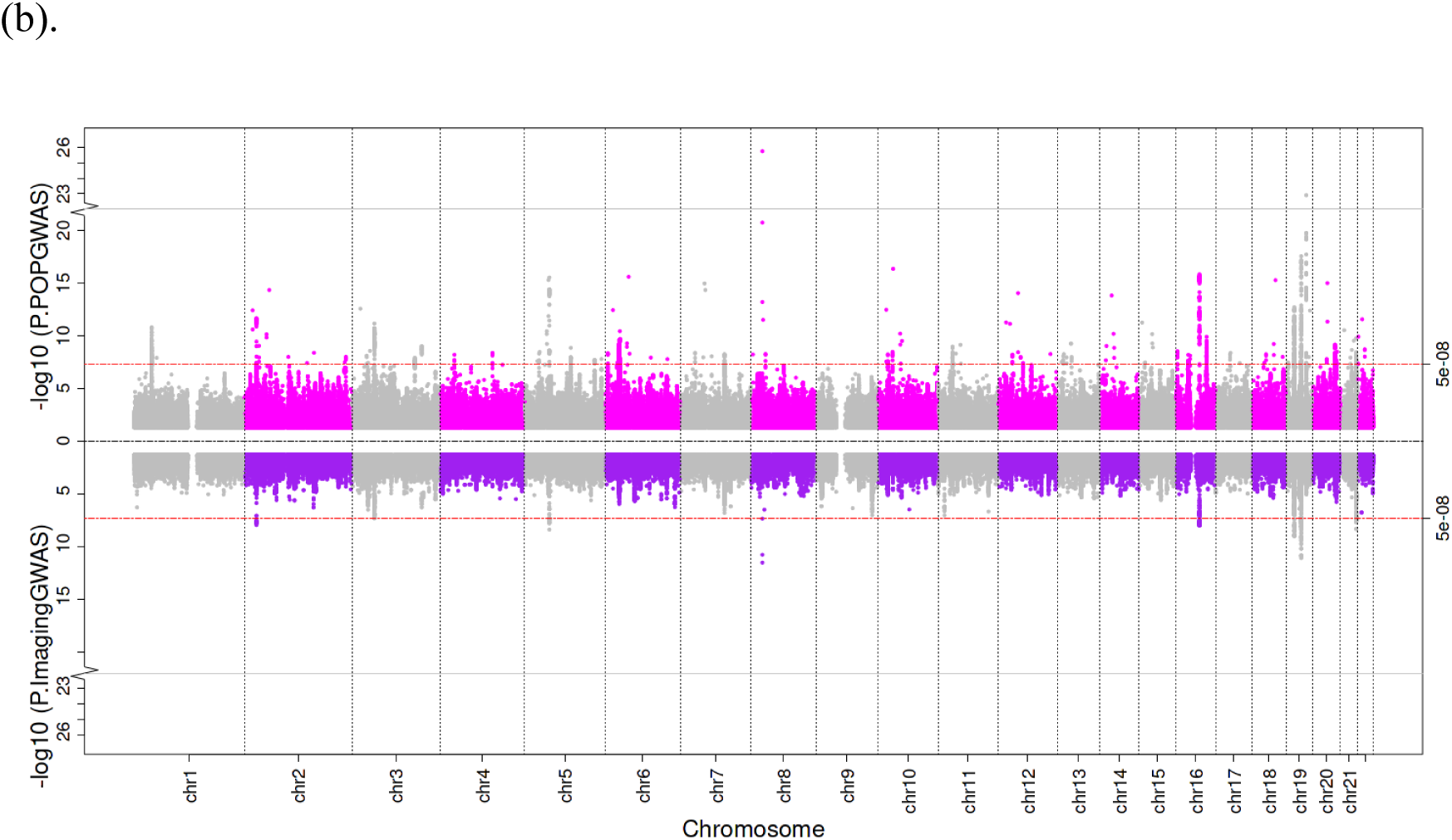
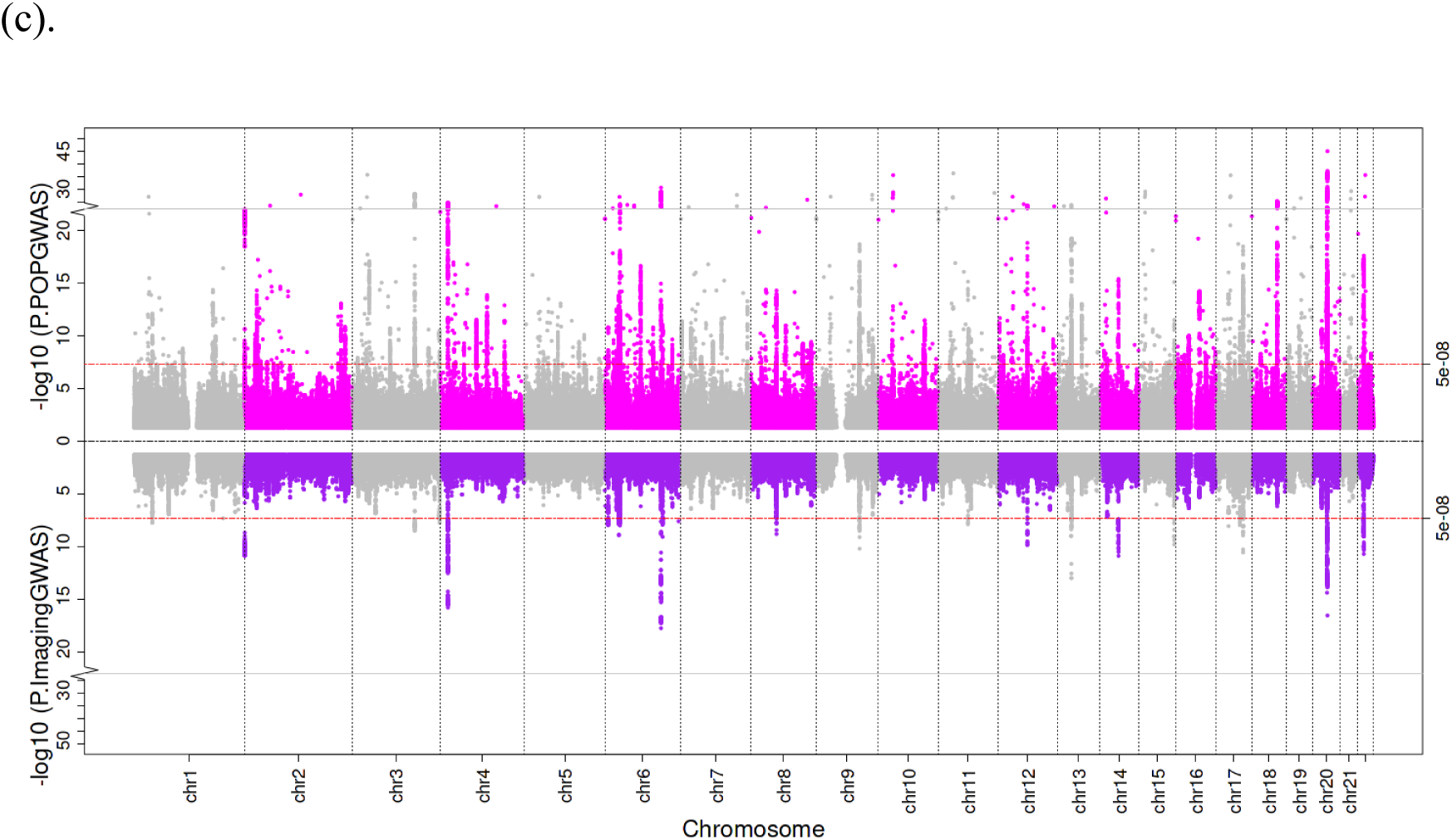
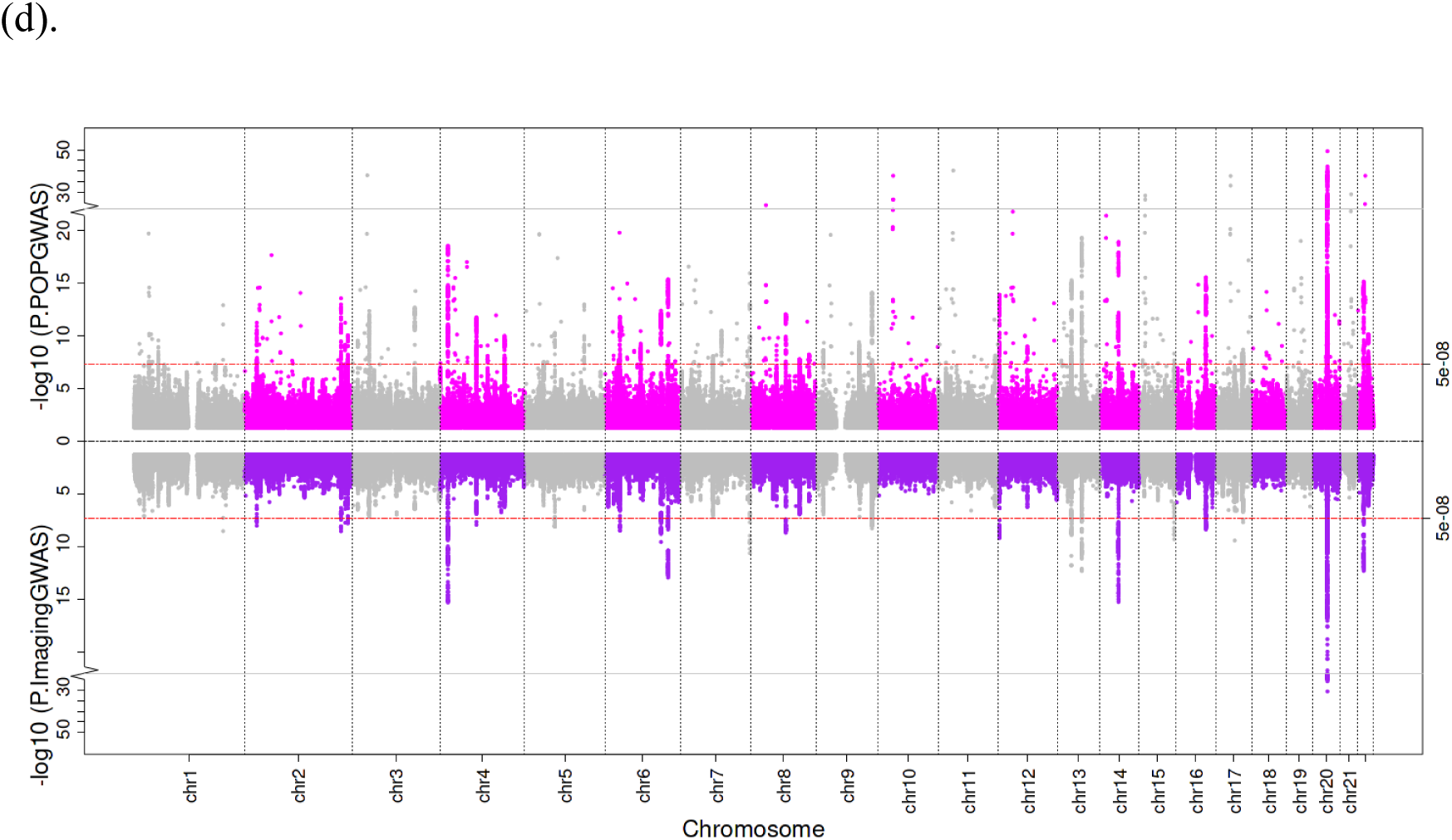
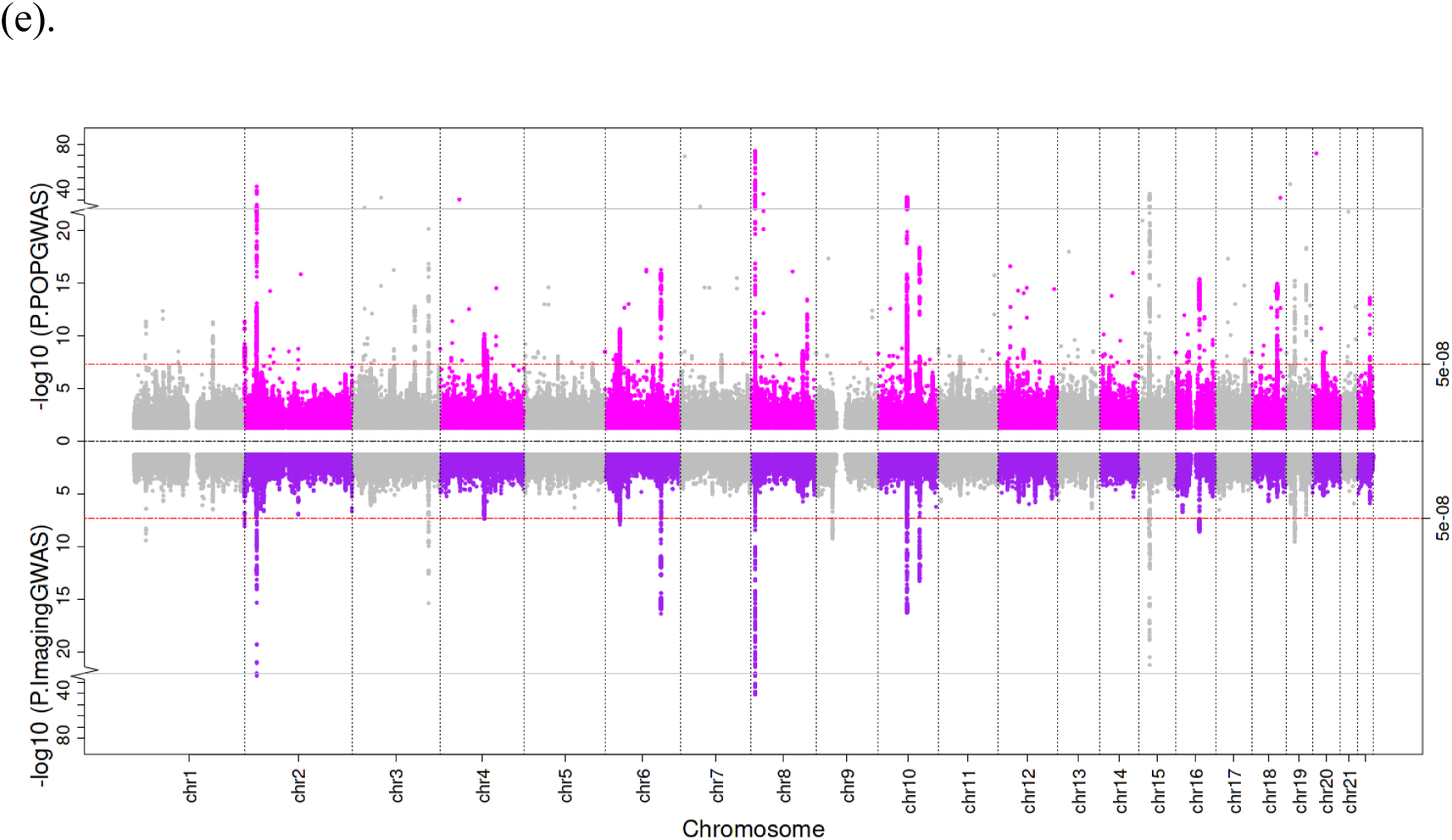
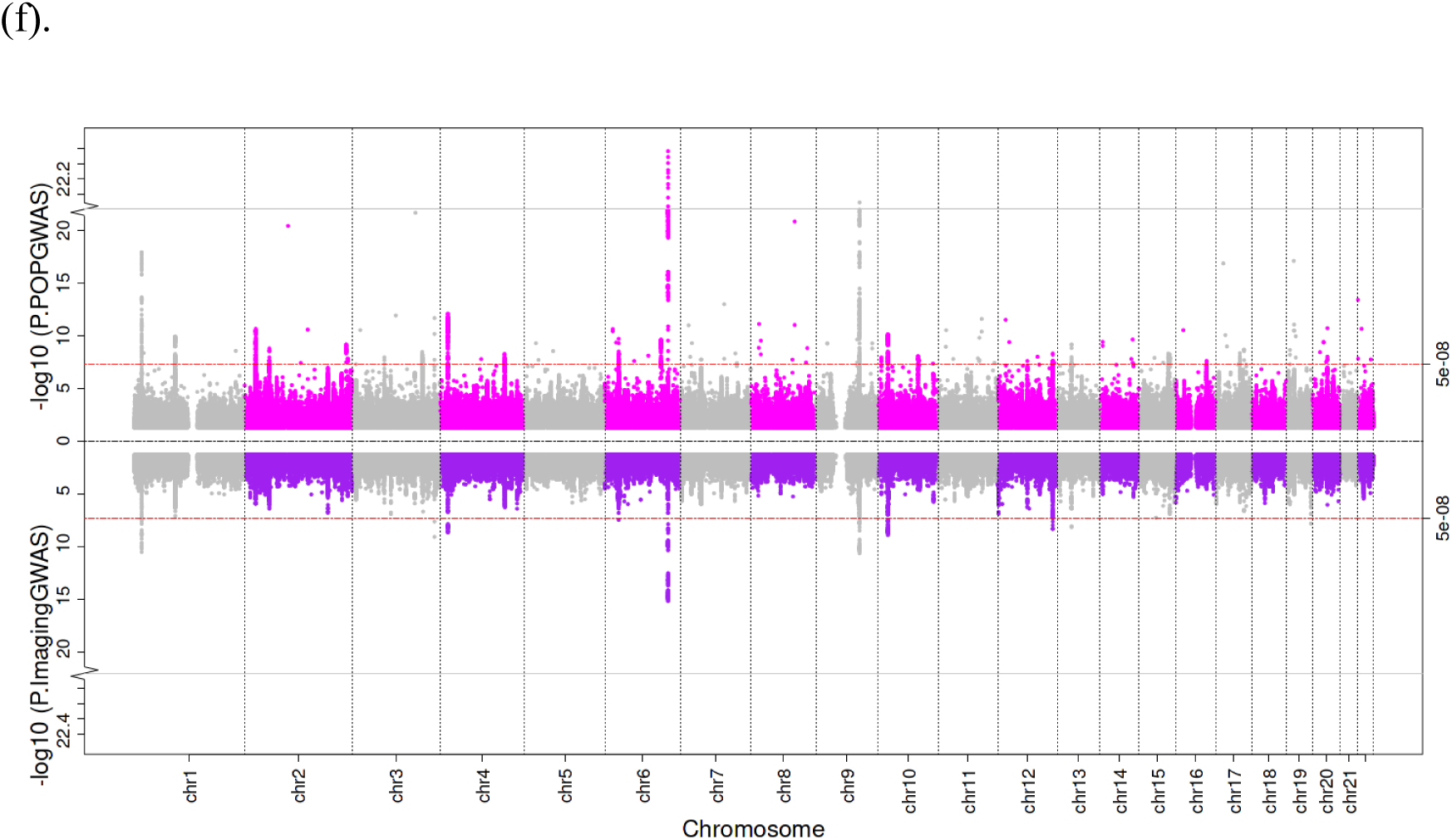
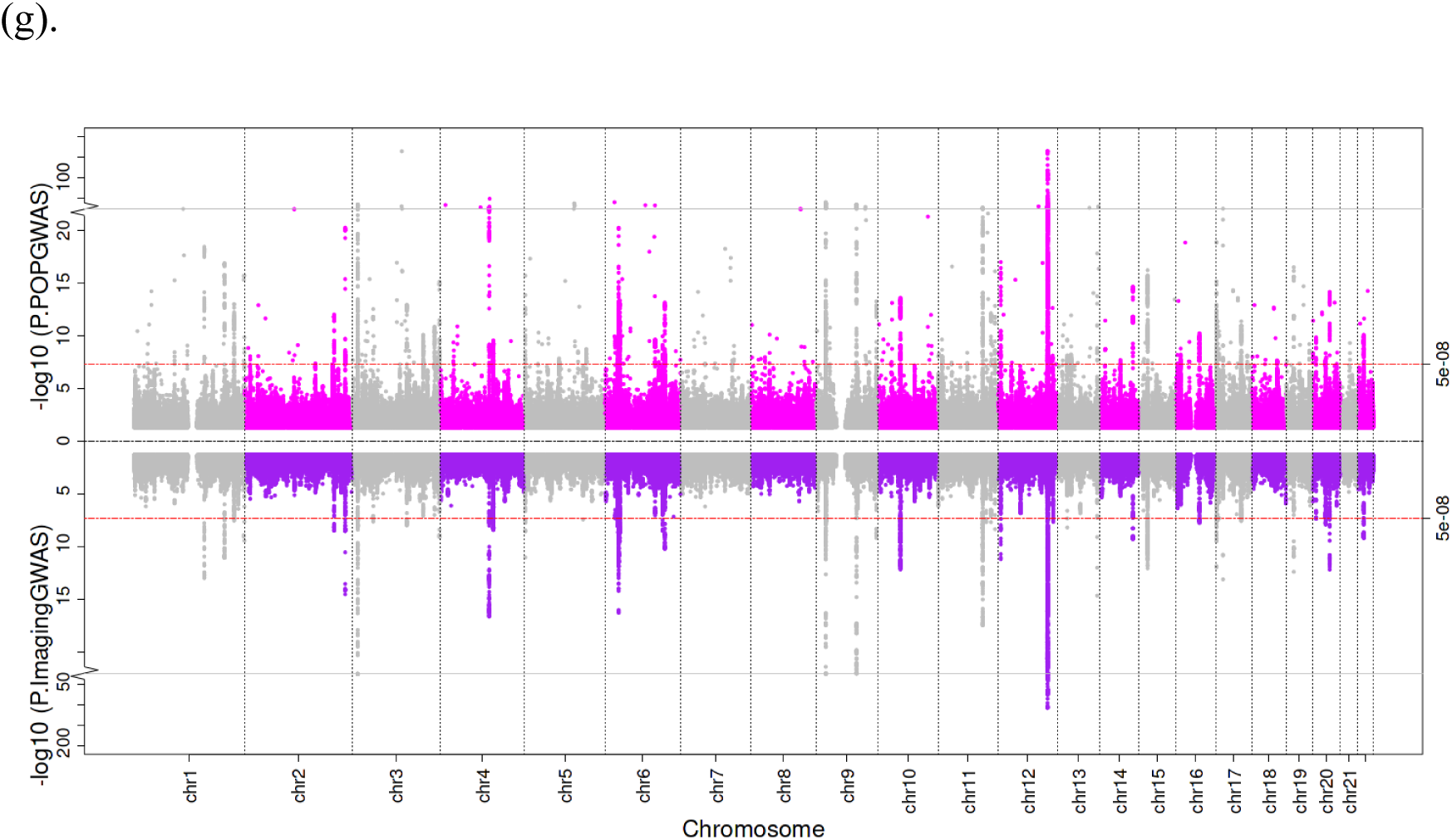
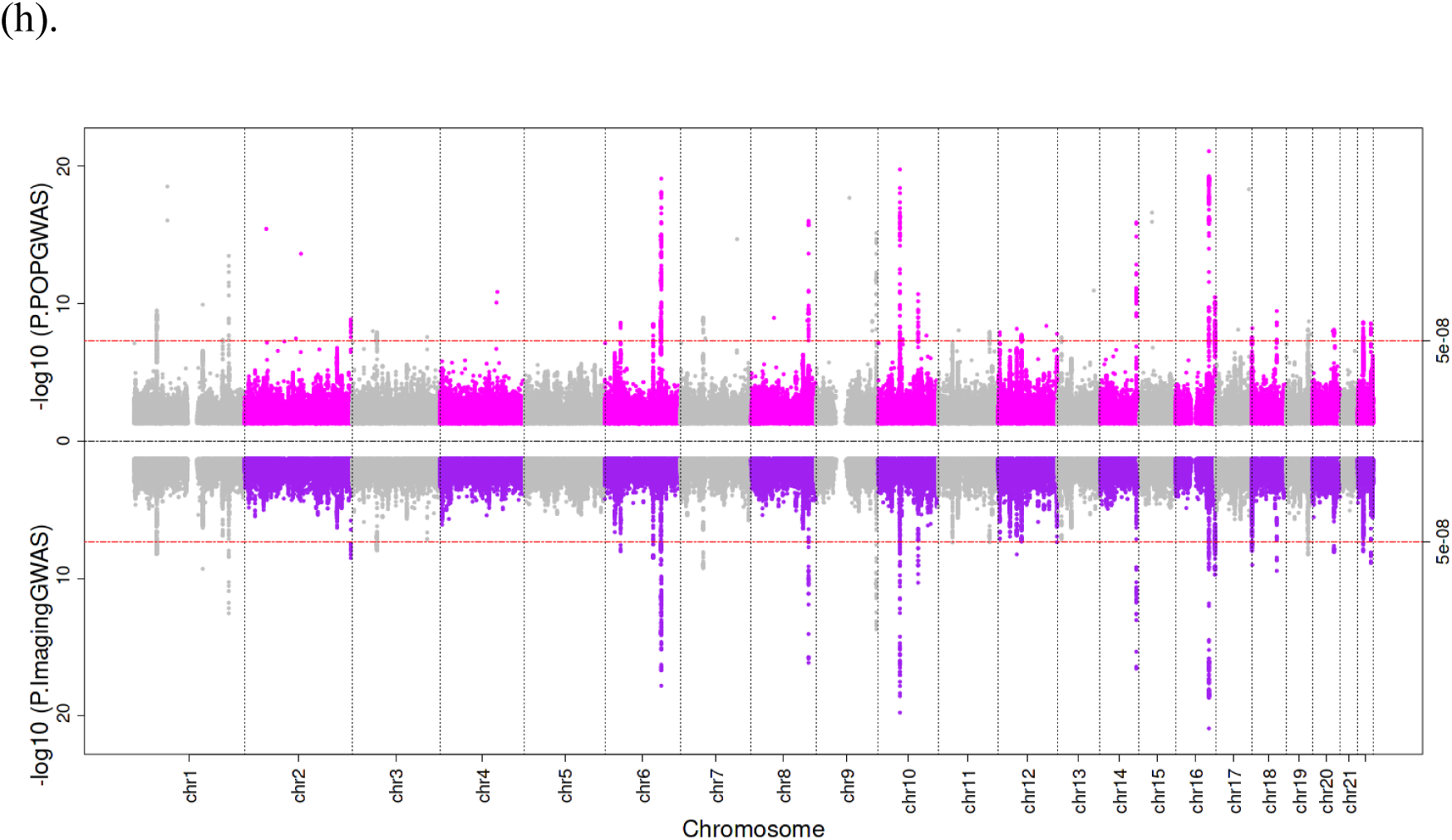
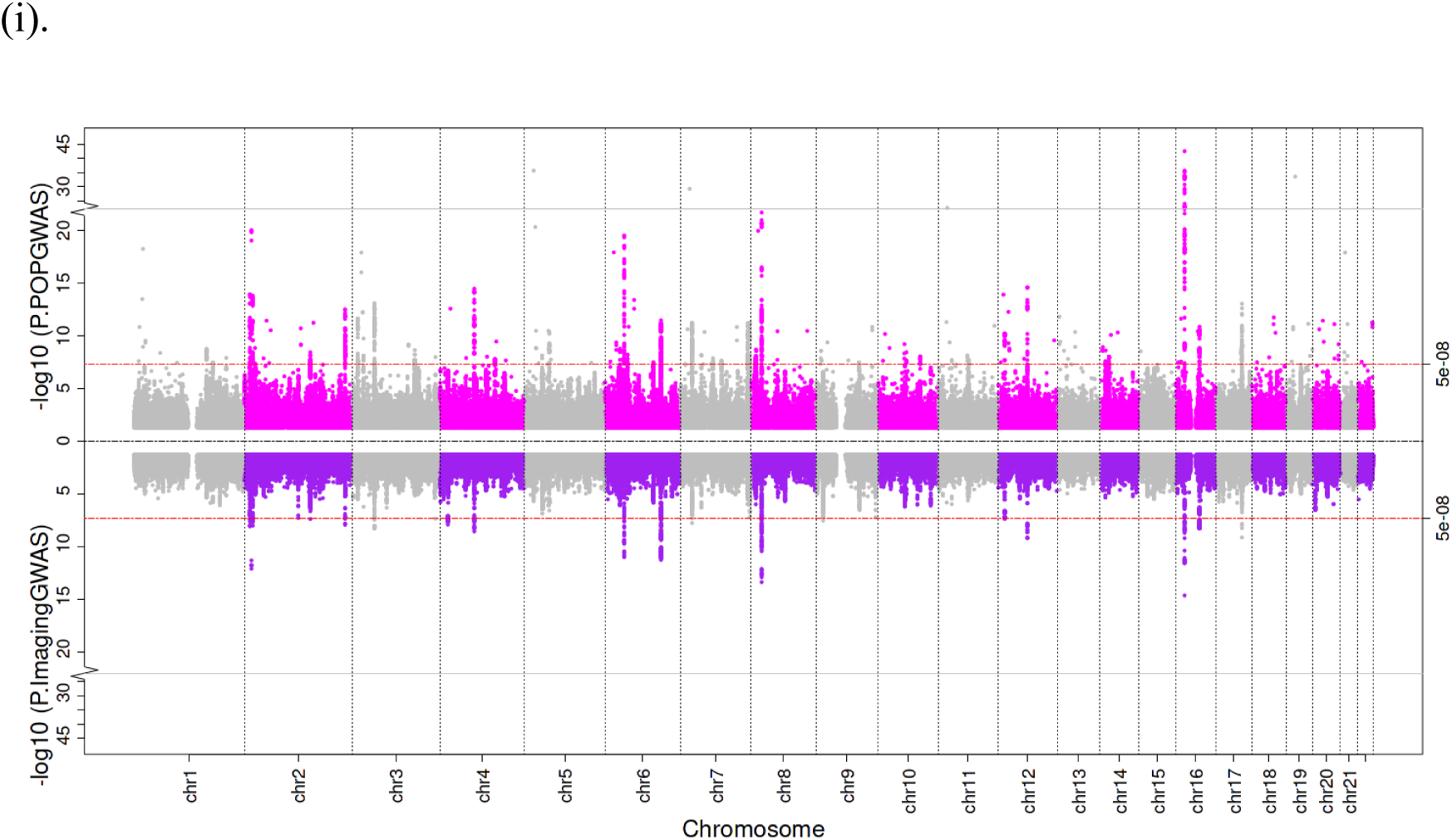
Miami plots of genome-wide association results for nine imputed IDPs. For each organ or tissue volume, the upper panel displays -log10 P-values from the POP-GWAS (non-imaging cohort), and the lower panel shows the corresponding values from the GWAS of directly measured MRI-derived traits (imaging cohort), plotted by genomic position across chromosomes (x-axis). The red dashed line indicates the genome-wide significance threshold (P = 5 × 10□□). Panels: (a) Abdominal subcutaneous fat, (b) Visceral fat, (c) Total muscle volume, (d) Iliopsoas muscle volume, (e) Liver volume, (f) Lung volume, (g) Spleen volume, (h) Pancreas volume, (i) Kidney volume.

Among the significant associations, several loci of interest were identified for each IDP (**Supplementary tables 6-14**). SNP-to-gene mapping and expression quantitative trait loci (eQTL) annotations were performed using FUMA v1.5.2, based on GTEx v8 data. For abdominal subcutaneous fat, rs935166 was associated with *KHK* expression in subcutaneous adipose tissue and its protein levels; rs2777888 was associated with *SEMA3F* expression in both visceral and subcutaneous adipose tissue and its protein levels; and rs56186137 was associated with *SULT1A1* expression in subcutaneous adipose tissue and its protein levels. For visceral fat, rs1701704 was identified as an eQTL for *SUOX* in both subcutaneous and visceral fat, with significant associations to its plasma protein levels. For total muscle volume, rs9894577 was identified as an eQTL for *HEXIM1*, which was also associated with its plasma protein levels; rs143384 was associated with both total muscle and iliopsoas muscle volume is an eQTL for *PROCR* and associated with its protein levels.

We also identified novel loci with no previously reported associations to human traits or diseases. These include rs4819362, associated with both abdominal subcutaneous and visceral fat, linked to the expression of *GATD3* in these fat depots and its plasma protein levels; rs2762599 (near *SFMBT2*) associated with lung volume; rs930005 (near *AP000797*) and rs72868785 (near *ADCYAP1*) associated with pancreas volume; rs4789614 (an intronic variant in *SDK2*) associated with spleen volume; and rs117635090 (near *LRFN5*) associated with total muscle volume.

Gene set enrichment analysis (**Supplementary tables 15–23**) identified several significant pathways associated with organ volumes. For total muscle volume, pathways involved in peptide hormone binding and kinase binding highlighted the importance of hormonal signaling and kinase activity in muscle mass regulation. For iliopsoas muscle volume, significant associations included skeletal system development. Liver volume was linked to pathways regulating RNA biosynthesis and overall biosynthetic activity, reflecting its role in maintaining metabolic homeostasis. Lung volume was associated with ligand-receptor interactions, Hedgehog signaling, and the PERK-mediated unfolded protein response, emphasizing the role of cellular signaling and stress responses. For spleen volume, pathways related to different cancers.

Tissue enrichment analysis revealed organ-specific transcriptional signatures, including the association of abdominal subcutaneous and visceral fat with brain tissue, liver volume with ovarian tissue, spleen volume with both spleen and whole blood, and pancreas volume with pancreatic tissue (**Supplementary figure 5**).

### Genetic correlation analyses

To understand the shared genetic architecture between organ volumes, we first examined genetic correlations among the nine tissue and organ volume measures. Most of the IDPs were positively correlated (**figure 2**). The highest correlation was observed between total muscle volume and iliopsoas muscle volume (rg = 0.82), followed by abdominal subcutaneous fat and visceral fat (rg = 0.79), reflecting expected anatomical and functional similarities. In contrast, lung volume showed negative genetic correlations with abdominal subcutaneous fat (rg = –0.32) and visceral fat (rg = –0.28).

**Figure 2.**
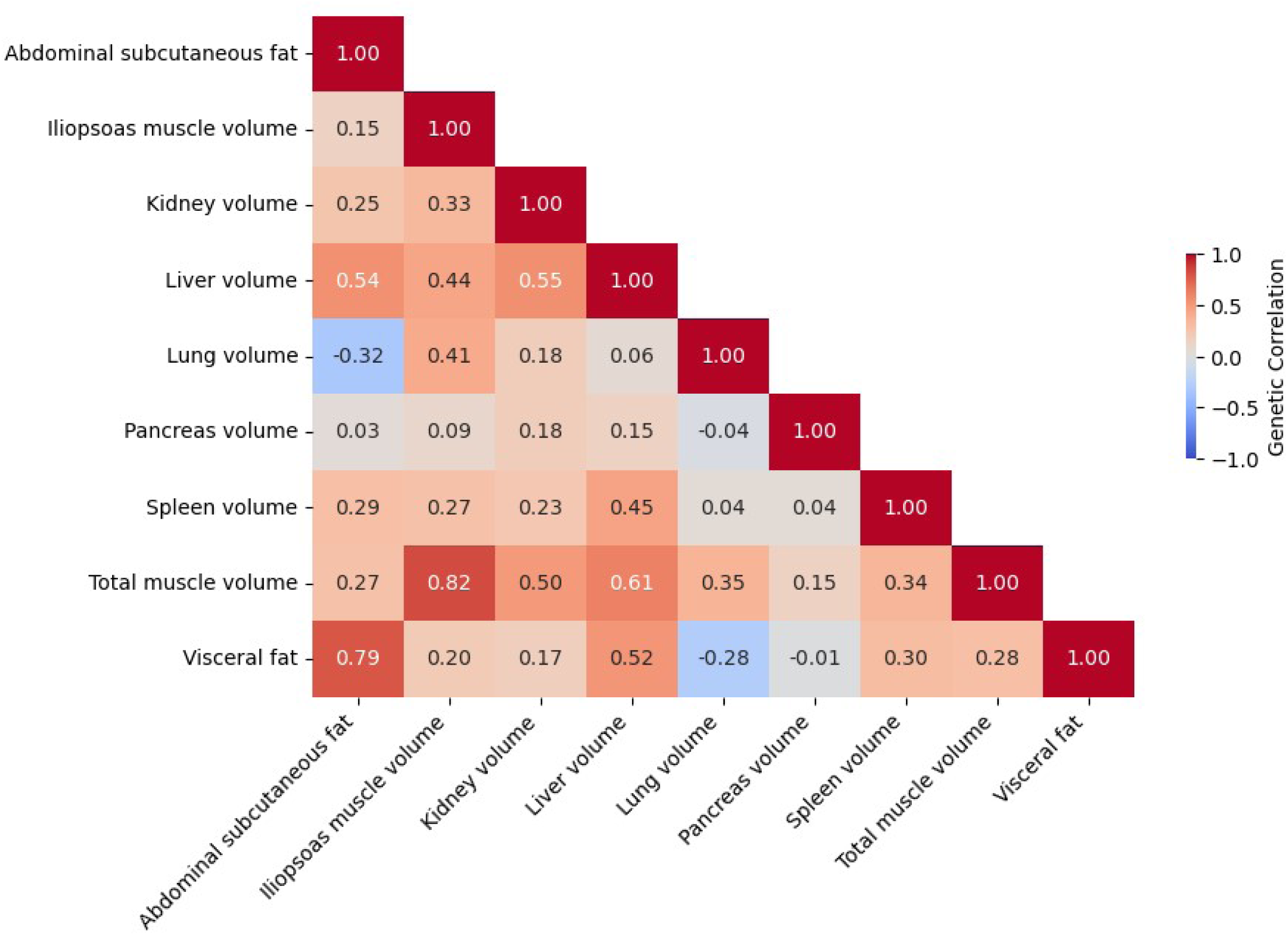
Genetic correlations among organ and tissue volumes. The heatmap shows pairwise genetic correlations (rg) between the nine IDPs. Red indicates positive correlation, blue indicates negative correlation, and colour intensity reflects the magnitude of the correlation.

We next explored genetic correlations between IDPs and a wide range of anthropometric, metabolic, and insulin-related biomarkers (**figures 3 and 4**). Abdominal subcutaneous fat, visceral fat, and liver volume had similar pattern of genetic correlation with other traits, including positive genetic correlations with obesity-related traits such as body fat mass, childhood and adult BMI and waist-hip ratio, measures of insulin resistance, CRP levels, triglycerides and negative correlations with sex hormone-binding globulin (SHBG) in both males and females, and with HDL cholesterol levels. Circulating leptin levels showed positive genetic correlation only with abdominal subcutaneous and visceral fat volumes, consistent with their role in adipose tissue biology.

**Figure 3.**
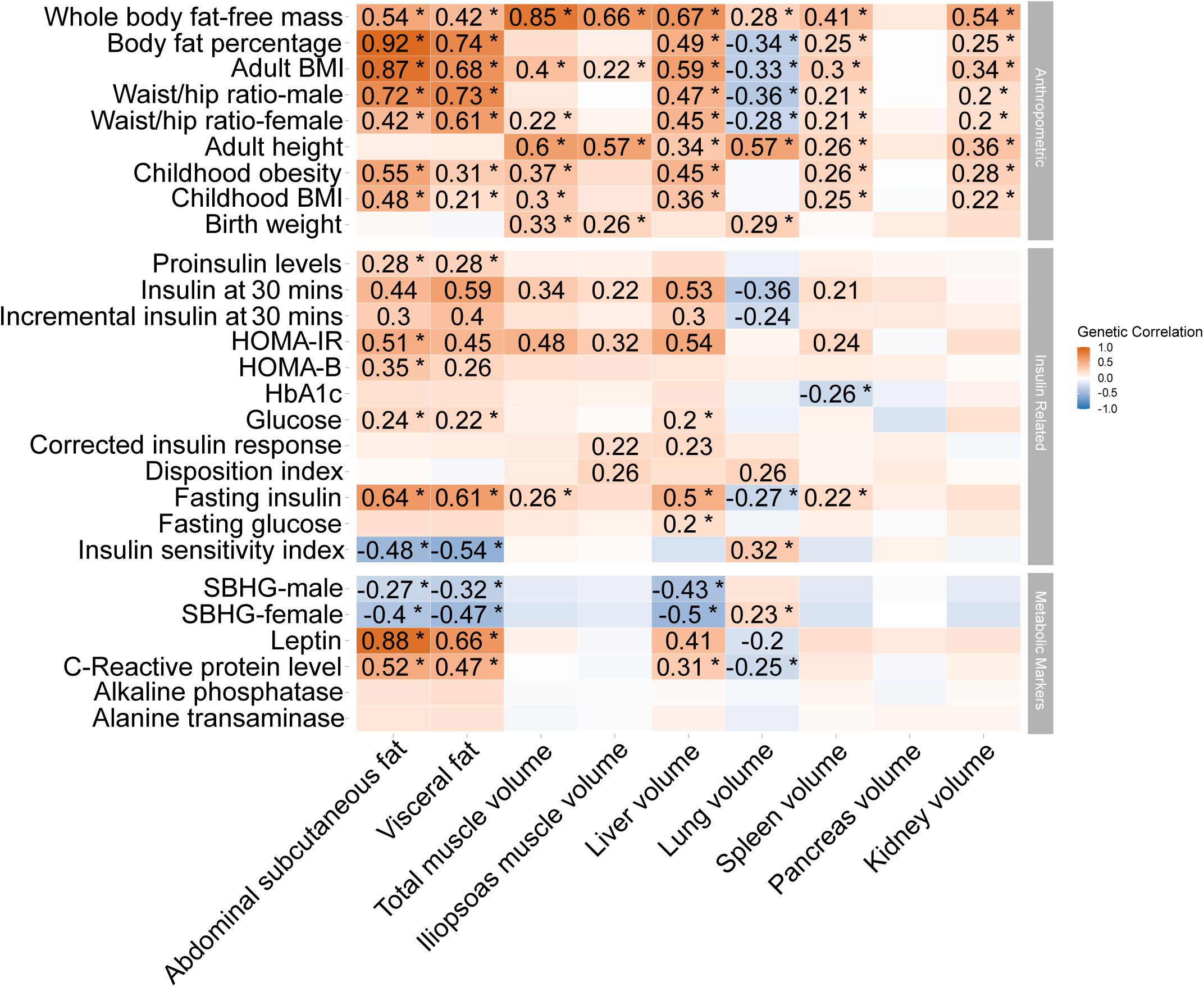
Genetic correlations between organ and tissue volumes and anthropometric, insulin-related, and metabolic biomarkers. Heatmap showing pairwise genetic correlations (rg). Red indicates positive correlation, blue indicates negative correlation. Asterisks denote associations passing Bonferroni correction for multiple testing (P < 0.0008; 0.05/60).

**Figure 4.**
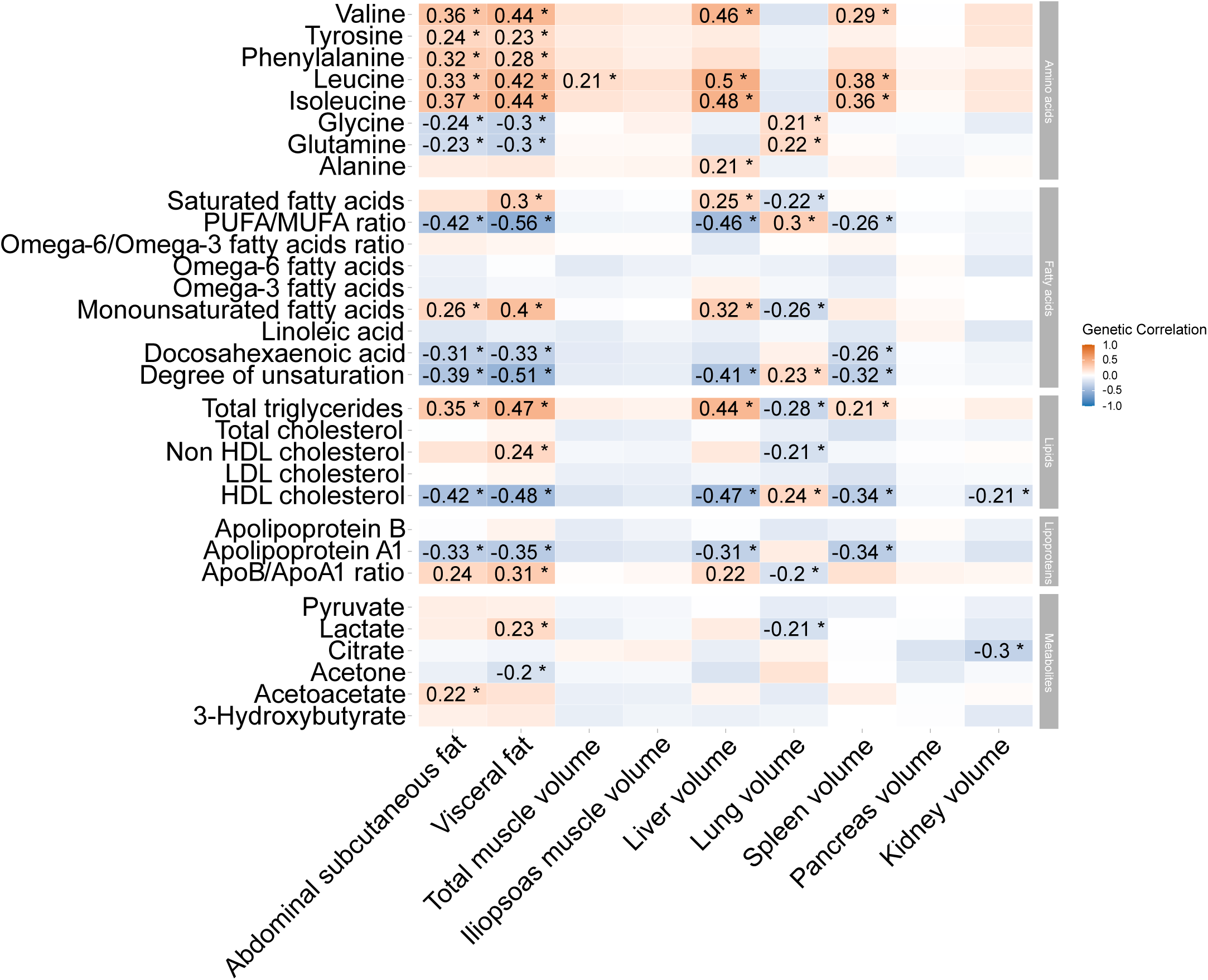
Genetic correlation between organ and tissue volumes and circulating metabolites. Heatmap showing pairwise genetic correlations (rg). Red indicates positive correlation, blue indicates negative correlation. Asterisks denote associations passing Bonferroni correction for multiple testing (P < 0.0008; 0.05/60).

Muscle-related IDPs, including iliopsoas and total muscle volume, were positively correlated with fat-free mass, BMI, and moderate birth weight. Lung volume showed a strong positive correlation with height. Spleen and kidney volumes were both positively correlated with fat-free mass, while pancreas volume did not show any strong or consistent genetic correlation with the traits examined.

### Mendelian randomization

We used Mendelian randomization (MR) to investigate potential causal relationships between genetically determined IDPs and disease outcomes (**Figure 5**). These analyses revealed both shared and organ-specific associations, highlighting the complex biological pathways linking organ size and fat distribution to disease risk.

**Figure 5.**
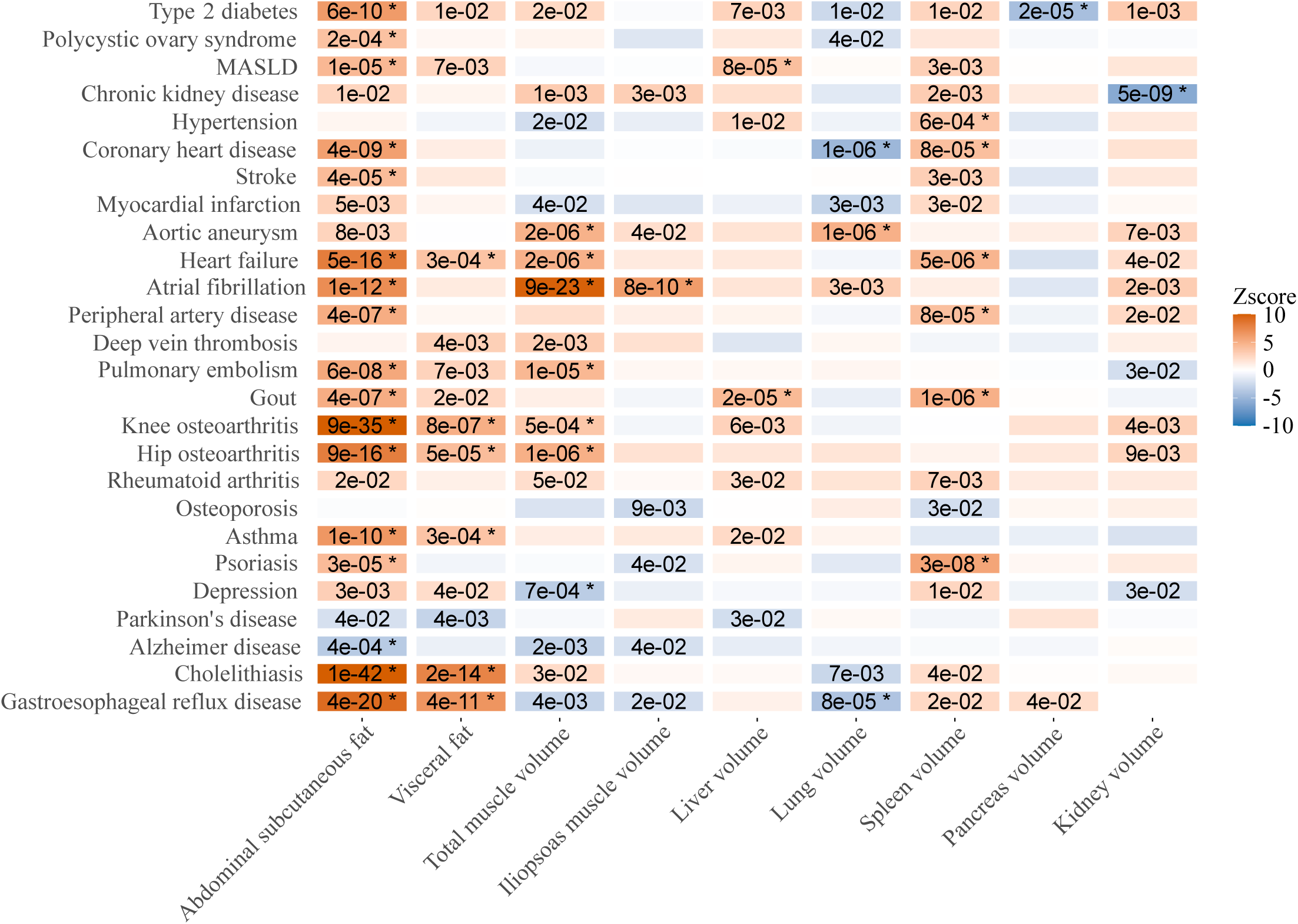
Mendelian randomization estimates for the causal effects of organ and tissue volumes on disease outcomes. Heatmap showing Z-scores from inverse-variance weighted (IVW) Mendelian randomization analyses across 35 disease outcomes using meta-analysed GWAS data (FinnGen and published studies). Colour intensity reflects effect direction and strength (Z-score), and asterisks indicate associations with Benjamini–Hochberg adjusted P < 0.05. Cell values represent FDR-corrected significance levels.

Genetically higher abdominal subcutaneous fat was associated with increased risks for several metabolic and inflammatory diseases, including type 2 diabetes, polycystic ovary syndrome, metabolic-associated steatotic liver disease (MASLD), cardiovascular disease, peripheral artery disease, pulmonary embolism, gout, osteoarthritis, asthma, psoriasis, cholelithiasis, and gastroesophageal reflux disease, and a lower risk of Alzheimer’s disease (all associations had multiple-test corrected P < 0.05).

Although visceral fat was strongly positively correlated with abdominal subcutaneous fat and shared a similar pattern of correlations with biomarkers, its disease associations differed. Genetically higher visceral fat was not associated with type 2 diabetes or cardiovascular disease but was associated with higher risk of heart failure, osteoarthritis, asthma, cholelithiasis, and gastroesophageal reflux disease.

Genetically higher total muscle volume was associated with higher risks of aortic aneurysm, hear failure, atrial fibrillation, thrombotic events, and osteoarthritis, and a lower risk of depression, highlighting the complex role of muscle mass in health and disease. Iliopsoas muscle volume was only associated with higher risk of atrial fibrillation.

Liver volume was associated with higher risks of MASLD and gout. Lung volume was associated with higher risks of aortic aneurysm but was associated with a lower risk of coronary heart disease and gastroesophageal reflux disease. Spleen volume was associated with higher risk of hypertension, coronary heart disease, heart failure, peripheral artery disease, gout, and psoriasis.

Kidney volume was associated with a lower risk of chronic kidney disease, suggesting a protective role of larger kidneys in renal health. Lastly, pancreas volume was only associated with lower risk of type 2 diabetes.

## Discussion

This study demonstrates the power of integrating machine learning-assisted phenotype imputation combined with the POP-GWAS framework to enable large-scale genetic discovery for costly, high-dimensional phenotypes such as MRI-derived organ volumes. While POP-GWAS has previously been applied to single traits such as bone mineral density, this work represents the first application of the method across a broad panel of MRI-derived organ and tissue volumes —including adiposity depots, muscle, and vital organs — providing novel insights into the genetic architecture of human anatomy at an unprecedented scale.

By imputing IDPs in over 370,000 non-imaged participants and applying POP-GWAS to correct for imputation bias, we achieved effective sample sizes exceeding 100,000 for several traits. This yielded 452 independent genetic loci across nine IDPs, with many mapping to biologically plausible genes. For instance, *KHK*—a gene central to fructose metabolism—was associated with subcutaneous fat volume. Knockout mice lacking KHK showing partial protection against the detrimental effects of high-fat, high-fructose diets [17]. *KHK* is also a promising therapeutic target for metabolic diseases such as NAFLD, type 2 diabetes, and cardiovascular disease. For example, PF-06835919, a reversible inhibitor of *KHK*, has demonstrated efficacy in reducing liver fat and improving cardiovascular risk factors in preclinical models [17]. Rare variants in the *SEMA3* genes are enriched in people with severe obesity [18]. In zebrafish, disrupting the Sema3 signaling pathway caused increased adiposity [18]. *SEMA3F* signaling is involved in the development of the melanocortin circuits that regulate energy homeostasis and appetite[18]. The *SULT1A1* gene plays a significant role in fat metabolism and adipose tissue browning, with deletion of the gene linked to lower body weight and the conversion of white adipose tissue to a more metabolically active brown-like state[19]. For muscle mass, we identified variants in *HEXIM1*, a gene with a crucial role in muscle regeneration and potential as a therapeutic target for degenerative muscular diseases [20].

Mendelian randomization analysis revealed that each organ volume has a distinct and unique effect on the risk of various diseases. Our findings align with existing literature linking genetically higher abdominal subcutaneous fat to various metabolic and inflammatory diseases. The association with type 2 diabetes reflects its role in promoting insulin resistance and systemic inflammation[21], while the connection to polycystic ovary syndrome (PCOS) is consistent with evidence linking excess adiposity to hormonal imbalances and metabolic dysfunctions characteristic of PCOS[22]. Similarly, the observed link to MASLD aligns with studies showing that subcutaneous fat contributes to hepatic fat deposition through lipolysis and inflammation[23]. The link to gout is supported by evidence connecting metabolic syndrome components, such as hyperuricemia, to gout risk [24], while the association with osteoarthritis reflects the impact of mechanical loading and inflammatory adipokines on joint degeneration [25]. The connection to asthma aligns with studies demonstrating obesity-related respiratory inflammation [26], and the link to psoriasis corroborates findings that systemic inflammation driven by obesity exacerbates psoriasis pathogenesis [27].

For visceral fat, the association with heart failure aligns with evidence showing that visceral fat contributes to systemic inflammation, vascular dysfunction, and cardiac remodeling, all of which exacerbate cardiovascular strain and heart failure risk [28] [29]. Similarly, the link to osteoarthritis reflects the dual impact of mechanical stress from increased adiposity and the pro-inflammatory cytokines secreted by visceral fat, which accelerate joint degeneration [25]. The connection to asthma is consistent with studies indicating that visceral fat releases inflammatory mediators that exacerbate airway inflammation and reduce pulmonary function [30]. The observed relationship with cholelithiasis (gallstones) aligns with the role of visceral fat in increasing cholesterol secretion into bile and impairing gallbladder motility, both of which contribute to gallstone formation [31]. Lastly, the association with reflux is likely mediated by the mechanical pressure exerted by visceral fat on the abdomen, which promotes gastroesophageal reflux, combined with the pro-inflammatory state that may impair esophageal motility and function [32].

Total muscle volume and iliopsoas muscle volume displayed partly overlapping but distinct patterns. For muscle volume, the association with aortic aneurysm and atrial fibrillation, may be partly explained by increased blood flow demands and vascular strain linked to greater muscle mass [33]. The connection to osteoarthritis likely reflects the mechanical loading exerted on joints by increased muscle mass [25]. Conversely, the protective association with depression aligns with evidence that higher muscle mass improves mental health through enhanced physical function, reduced inflammation, and hormonal regulation [34] [35].

Other IDPs exhibited similarly nuanced associations. Liver volume was associated with higher risks of MASLD and gout, aligning with the liver’s central role in lipid metabolism and uric acid production [36] [37]. These findings suggest that increased liver size may reflect a compensatory response to metabolic overload or inflammation, contributing to the pathophysiology of these conditions. Lung volume showed a mixed pattern of associations, with larger lungs linked to an increased risk of aortic aneurysm but a reduced risk of coronary heart disease and gastroesophageal reflux disease (GERD). The elevated risk of aortic aneurysm may be related to structural changes or vascular strain associated with larger lungs, while the protective effects against coronary heart disease and GERD could be due to improved respiratory efficiency or reduced abdominal pressure.

Spleen volume was associated with a broad range of conditions, including hypertension, coronary heart disease, heart failure, peripheral artery disease, gout, and psoriasis. These associations highlight the spleen’s critical role in immune regulation and inflammation, as splenic enlargement often reflects systemic immune activation or inflammatory states contributing to cardiovascular and metabolic diseases. Kidney volume was associated with a lower risk of chronic kidney disease, suggesting a protective role of larger kidneys in maintaining renal function and compensating for potential damage. Similarly, a larger pancreas volume was associated with a lower risk of type 2 diabetes as previously shown [37].

Collectively, these findings establish that organ size is not merely a passive correlate of health status but an active determinant of disease susceptibility. By leveraging imputed MRI-derived traits in a population-scale dataset, we uncovered causal mechanisms that were previously inaccessible due to the high cost and limited sample sizes of imaging studies. This study demonstrates that high-dimensional imaging phenotypes can be imputed with sufficient accuracy to enable robust downstream genetic analyses, particularly when paired with bias-corrected frameworks such as POP-GWAS. We observed strong consistency between effect estimates derived from directly measured and imputed traits, reinforcing the validity of our approach. while imputed IDPs may be noisier than directly measured values, our validation analyses showed high concordance between measured and imputed GWAS results, and the use of POP-GWAS ensured bias correction for imputation-related attenuation. This allowed us to unlock a substantially larger discovery sample and identify hundreds of novel loci with confidence. Moreover, functional follow-up analyses supported the biological relevance of the identified associations. The substantial increase in sample size also improved the power and resolution of Mendelian randomisation analyses, enabling the detection of more nuanced and disease-specific causal relationships across 35 complex traits.

However, some limitations should be acknowledged. Our analyses were limited to participants of White British ancestry, and future replication in more diverse populations is essential to ensure broader applicability. Although POP-GWAS corrects for attenuation bias introduced during imputation, it still depends on the quality and diversity of features used in the predictive model. Organ-specific variation in imputation accuracy may also have influenced power across traits. Moreover, while we identified several novel loci, functional interpretation remains speculative without experimental validation. Follow-up studies integrating transcriptomics, proteomics, and spatial gene expression will be critical to pinpoint causal mechanisms. Finally, longitudinal imaging and clinical data will be important for understanding how genetic variation influences organ size trajectories over time and their interactions with environmental exposures.

## Conclusions

This work provides a scalable and generalisable framework for expanding genetic discovery to phenotypes that are otherwise limited by measurement cost or logistical constraints. By integrating deep learning-based image analysis, biomarker-informed imputation, and bias-corrected GWAS methods, we demonstrated that imaging-derived traits can be robustly studied at population scale. Our findings reveal distinct organ-specific genetic architectures and highlight new causal pathways linking organ size to disease. This framework advances understanding of organ-specific biology and provides a blueprint for leveraging the remaining >60,000 MRI scans in the UK Biobank to accelerate genetic discovery and uncover mechanisms of disease. As biobanks increasingly integrate imaging and multi-omics data, the ability to scale genetic discovery using imputation frameworks such as ours may enable systematic identification of novel disease mechanisms, therapeutic targets, and stratification biomarkers to advance precision health.

## Methods

### Study design

We applied a machine learning-assisted GWAS framework to investigate the genetic determinants of MRI-derived organ and tissue volumes and their relationships with cvarious diseases (**Supplementary figure 1**). The UK Biobank cohort was stratified into two groups: an imaging cohort (n = 37,589) with available abdominal MRI scans, and a non-imaging cohort (370,568) without imaging data. Using the imaging cohort as a training set, we developed machine learning models to impute nine IDPs across the non-imaging cohort, leveraging 382 routinely collected biomarkers. This phenotypic imputation substantially increased the effective sample size, allowing for more powerful genetic discovery. We then conducted GWAS on the imputed IDPs in the non-imaging cohort, applying the POP-GWAS method to correct for biases introduced by imputation. Additional analyses included heritability estimation, genetic correlation with complex traits, functional enrichment, protein QTL integration, and Mendelian randomisation (MR) to explore biological mechanisms and disease relevance.

### Image acquisition and segmentation

In the imaging cohort, abdominal MRI scans were acquired using Siemens Aera 1.5T scanners (Syngo MR D13) as part of the UK Biobank imaging protocol [7]. Participants underwent both neck-to-knee Dixon MRI and single-slice multiecho MRI, covering key organ systems and fat depots. Dixon imaging data were assembled into 3D volumes with automated fat-water swap detection and correction.

Image processing was performed using an updated 3D U-Net architecture, implemented in TensorFlow and inspired by segmentation networks used in label-free microscopy. The model was trained on at least 100 manually annotated scans covering multiple organs and structures. Out-of-sample evaluation showed average Dice coefficients exceeding 0.8, confirming segmentation accuracy.

The network received five input channels — fat, water, in-phase, out-of-phase, and a body mask — and output binary segmentation masks via organ-specific supervision. The architecture used 72 channels at the outer layers, increasing up to 1,152 channels in deeper layers. It included skip connections, convolution-transpose upsampling, and multi-head outputs to account for overlapping anatomical structures (e.g., fat and muscle).

The model was trained on 80,000 96 × 96 × 96 voxel patches using the Adam optimiser with a learning rate decaying quadratically from 1e−5 to 1e−7. Loss was computed as the sum of Dice coefficient and binary cross-entropy across all organs. A batch size of six was used, with optimised GPU memory management for large 3D tensors [8] (see **Supplementary Methods** for details).

### Data augmentation and quality control

To enhance generalisation, data augmentation was performed on-the-fly on GPU. This included smooth 3D elastic warping based on Gaussian process-sampled optical flow vectors. Warps were applied consistently to both input images and training masks using polyharmonic spline interpolation, allowing realistic anatomical distortions.

Inference outputs were binarised using Otsu thresholding, and disconnected components were removed. Final organ volumes were calculated by summing mask voxels and multiplying by voxel resolution.

Segmentation-derived features such as visceral fat and abdominal subcutaneous fat were generated by subtracting organ and tissue masks from surrounding fat regions. Quality control involved both visual inspection of extreme cases and random spot checks of segmentation outputs. The training set was iteratively expanded to include failure cases until no systematic segmentation errors were observed (see **Supplementary Methods** for details).

**Supplementary Table 1** lists the IDPs used, including abdominal subcutaneous and visceral fat, iliopsoas muscle (left and right), pancreas, kidneys (left and right), spleen, liver, lungs, and total muscle volume.

### Phenotypic imputation

To extend genetic discovery beyond the subset with MRI data, we developed machine learning models to impute imaging-derived phenotypes (IDPs) in participants without scans. The UK Biobank imaging cohort included 42,963 participants with abdominal MRI scans, of which 37,705 individuals with high-quality organ volume measurements were designated as the training set for model development. An additional 5,258 participants were held out as an independent test set for evaluating model performance and imputation accuracy. The remaining 370,568 participants without imaging data formed the non-imaging cohort, in which IDPs were subsequently imputed using the trained models. We focused on nine IDPs: abdominal subcutaneous fat, visceral fat, iliopsoas muscle (left and right), total muscle volume, kidneys (left and right), liver, lungs, pancreas, and spleen.

Imputation models were built using up to 382 routinely collected variables—including anthropometric traits, lifestyle factors, clinical biochemistry, blood counts, metabolomics, and body impedance—available across the entire UK Biobank population. All variables were harmonised, standardised, and filtered to remove multicollinearity (pairwise Pearson correlation > 0.9) (see **Supplementary Methods** for details).

We evaluated four predictive modelling approaches for each IDP: linear regression, random forests, gradient boosting machines (GBM), and support vector machines (SVM) with a radial basis kernel. Ten-fold cross-validation was used to optimise hyperparameters and assess generalisation. Performance metrics included R², root mean squared error (RMSE), and mean absolute error (MAE), evaluated within the imaging cohort by comparing predicted and observed MRI-derived values.

SVMs consistently showed the best predictive accuracy for most IDPs, particularly fat and muscle depots. However, for spleen and kidney volumes, GBM models provided slightly better performance and were therefore selected. Model selection was based on highest cross-validated R². Final imputation was performed in the non-imaging cohort using the best-performing model for each IDP. Additional modelling details, performance metrics, and cross-validation results are provided in the **Supplementary Methods and Supplementary Tables 3-4**.

### Genetic association studies and POP-GWAS framework

To maximise discovery power while addressing bias introduced by phenotype imputation, we applied POP-GWAS [6], a framework specifically designed for GWAS of machine learning-imputed traits. In large-scale datasets such as UK Biobank, where only a subset of participants undergo imaging, imputation allows substantial sample size gains but can introduce systematic bias — typically attenuation of SNP effect sizes — due to imperfect prediction of the underlying traits.

POP-GWAS addresses this by using three separate GWAS analyses to estimate and correct for the bias introduced during imputation: (1) Imaging cohort (direct): GWAS of directly measured IDPs (N = 37,589), (2) Imaging cohort (imputed): GWAS of imputed IDPs in the same imaging cohort (i.e., comparing imputed vs. measured effects within the same individuals), and (3) Non-imaging cohort (imputed): GWAS of imputed IDPs in the remaining UK Biobank participants without MRI scans (N = 370,568).

The key insight of POP-GWAS is that the difference in effect sizes between the measured and imputed traits within the imaging cohort provides an estimate of imputation-induced bias. This estimate is then used to correct effect sizes in the non-imaging cohort, yielding unbiased and well-calibrated association statistics from the much larger sample. All corrections are performed using summary statistics only, making POP-GWAS computationally efficient and flexible.

The strength of bias is estimated using bivariate linkage disequilibrium score regression (LDSC), which quantifies the attenuation between measured and imputed traits in the imaging cohort. This value is then used to rescale the SNP effects from the GWAS of the non-imaging cohort. The imaging cohort is used solely to calibrate and adjust estimates from the non-imaging cohort, which serves as the discovery dataset and therefore, the results presented are from the non-imaging dataset and not meta-analyses with the imaging cohort.

We implemented GWAS in each dataset (imaging cohort with directly measured IDPs, imaging cohort with imputed IDPs, and non-imaging cohort with imputed IDPs) using REGENIE v3.1.1 [9], a linear mixed model approach that accounts for population structure and relatedness. The analyses were restricted to participants self-identified as ‘White British’ who clustered with this group in principal component analysis (PCA). Exclusion criteria included participants with sex chromosome anomalies, sex discrepancies, heterozygosity outliers, and genotype call rate outliers [10]. Covariates included age, squared age, sex, genotyping array, imaging center, and the first 10 genotype-related principal components. IDPs were inverse normal transformed before analysis. The final dataset comprised 9,788,243 SNPs after filtering imputed SNPs with a minor allele frequency (MAF) > 0.01 and an INFO score > 0.9.

The effective sample size for each imputed IDP was directly estimated by the POP-GWAS framework, which calculates the sample size equivalent to the corrected summary statistics after accounting for imputation-induced bias.

### Visualization and functional annotation

We used PheWeb [11] for visualization, including Manhattan and Q-Q plots and EasyStrata [12] for Miami plots. Functional mapping, gene set enrichment analysis, and tissue/cell-type annotations were performed using FUMA v1.5.2 with default MAGMA settings [13] v1.5.2.

Protein quantitative trait loci (p-QTL) analysis was conducted using data from the UK Biobank Pharma Proteomics Project[14], which involved 2,940 p-QTLs from approximately 52,000 participants. Proteins associated with significant GWAS variants were included if their p-QTL analysis p-value was < 1.7 × 10−11. This approach provided insights into protein-level associations and their potential biological significance in the context of the identified genetic variants.

### Genetic correlation and heritability analysis

We used LDSC [15] to compute heritability and genetic correlation. Genetic correlations were computed between organ volumes and various biomarkers, metabolic traits, lifestyle factors, and disease outcomes. To account for multiple testing, a Bonferroni-corrected significance threshold of p<0.05/60 (number of biomarkers) = 0.0008 was applied. A detailed list of the analyzed traits and their sources is provided in **Supplementary table 24**. LDSC was performed using precomputed linkage disequilibrium (LD) scores from European ancestry samples, ensuring robust estimates of heritability and correlations.

### Mendelian randomization

To explore the potential biological relevance of organ and tissue volumes, we conducted Mendelian randomisation (MR) analyses as a supporting line of evidence. These analyses were designed to illustrate how imaging-derived phenotypes (IDPs) imputed at scale can reveal insights into disease mechanisms when integrated with large-scale GWAS datasets.

We used the TwoSampleMR package (version 0.6.1) in R [16] to estimate causal effects of IDPs on disease outcomes. Genetic instruments for each IDP were selected from the POP-GWAS results using a genome-wide significance threshold (P < 5 × 10□□), and LD clumping was applied (r² < 0.01, 10,000 kb window) using European-ancestry reference data. All effect alleles were aligned to represent IDP-increasing alleles (organ volume-increasing alleles), ensuring consistent interpretation across traits.

Given that the primary aim of the study was to validate and demonstrate the utility of machine learning-assisted IDP imputation, we focused on a single MR method, inverse-variance weighted (IVW) analysis, which provides a powerful and interpretable estimate under the assumption of no directional pleiotropy. For each IDP–outcome pair, we report effect size (β) per 1 SD increase in IDP, 95% confidence interval (CI), and P-value.

We tested 35 disease outcomes. Details of trait definitions and sample sizes are provided in **Supplementary Tables 25-26**. Where available, results were meta-analysed across FinnGen (35 outcomes available from FinnGen Release 7) and public GWAS (32 outcomes) using the metafor R package (version 4.6.0), and statistical significance was assessed using a Bonferroni-corrected threshold to account for multiple testing.

We did not perform bidirectional MR or pleiotropy-robust methods, as the MR was not intended to establish definitive causal claims but rather to demonstrate the feasibility and interpretability of downstream analyses using imputed IDPs. Future studies using longitudinal imaging data or clinical endpoints may build upon this framework to assess causal directionality more comprehensively.

## Conflict of interest

MC and ES are employees of Calico Life Sciences LLC.

## Financial support statement

H.Y. is funded by Diabetes UK (grant 23/0006598) and Calico Life Sciences LLC.

## Author Contributions and Guarantor Statement

M.C., performed the GWAS. B.W. performed the imputation. A.N., and A.A. performed the pop-GWAS and analysed the data. H.Y. designed the study and wrote the manuscript. M.J., E.L.T. and J.D.B. provided all the MRI derived IDPs. All authors contributed to the writing, reviewing, editing and approving the manuscript. H.Y. is the guarantor of this work and, as such, has full access to all the data in the study and takes responsibility for the integrity of the data and the accuracy of the data analysis.

## Data availability

Our research was conducted using UK Biobank data. Under the standard UK Biobank data sharing agreement, we (and other researchers) cannot directly share raw data obtained or derived from the UK Biobank. However, under this agreement, all the data generated, and methodologies used in this paper are returned by us to the UK Biobank, where they will be fully available. Access can be obtained directly from the UK Biobank to all bona fide researchers upon submitting a health-related research proposal to the UK Biobank https://www.ukbiobank.ac.uk.

## Acknowledgments

This research was conducted using the UK Biobank Resource under Application Number 44584. We also want to acknowledge the participants and investigators of the FinnGen study.

