## Supplementary material for "Scaling genetic discovery for organ volumes using machine learning-assisted imputation and bias-corrected GWAS": popGWAS_supporting_information_23Sep2025.docx

### **Supplementary method**

**Deep Learning Image Processing Pipeline**

*Segmentation of organs, for volume assessment, from Dixon data*

We re-purposed an updated 3D iteration of the U-net architecture [1] based on label-free segmentation from 3D microscopy [2]. All algorithms were implemented in TensorFlow (TF). In order to produce sensible segmentations for quality control purposes on minimal data, we made the following choices. Training data is intrinsically scarce, and performance can always be improved with additional data. We pursued a multi-task approach [3] to improve data efficiency. The supervision loss consists of binary heads as opposed to multi-class classification because compartments can overlap spatially. We annotated multiple compartments and organs on the same individuals. Comparisons across datasets are also difficult because evaluation would be confounded by the specifics of how individuals are chosen, the conventions of annotation, and the specifics of data acquisition or processing.

Our implementation of U-net had 72 channels on the outside, and we capped the maximum number of channels in deeper layers of the network to 1152. We used concatenation on skip connections and convolution-transposes when upsampling. A heavily engineered system was used to stream large datasets efficiently and perform data augmentation on demand. To address computational bottlenecks, we encoded the 3D multichannel images as unrolled PNGs inside TFrecords. We relied on TF best practices to parallelise and streamline random batching during training. Data augmentation was performed on the fly on the GPU, and not pre-computed. We used a batch size of six, and some customised engineering was needed to accommodate very large tensors and total GPU memory use.

Input voxels were encoded into five channels: fat, water, in-phase, out-of-phase, and body mask. The body mask indicated whether a given voxel was inside the body. The neural network branched into a different logit head for supervision on each organ. Supervision included the sum of Dice coefficients [4] and binary cross-entropy across all organs.

Inspection of validation loss curves indicated that the use of batch normalisation and data augmentation provided sufficient regularisation. During training, the model utilised 80,000 96 × 96 × 96 patches as subsequently described, and the Adam optimiser learning rate was reduced from 1e-5 to 1e-7 following a quadratic decay. During inference, we used Otsu thresholding [5] to decode a binary decision for each voxel as to whether it was part of each given organ or not.

*Data augmentation*

Data augmentation included a 3D deformation to locally transform 3D data smoothly as a whole, rather than by slice. We iteratively batched a small number of individual voxels, assigned random Gaussian values and convolved noise with random-width Gaussian filters. The summed result was treated as a noise vector and added to the raw image dynamically. We also used a smooth elastic warping to augment the data. This augmentation assigned a different smooth 3D optical flow offset to each voxel in any spatial direction, which was effective since it could locally subsume a heterogeneous combination of commonly used spatial distortions. The same warping function was applied to training masks to ensure that supervision was consistent with input data.

Each final voxel obtained its value from a location offset by an optical flow vector sampled from a Gaussian process. To preserve visual details, voxels that were close together were sampled with strongly correlated optical flow offsets, while pairs further away were less correlated. To reduce the computational load in the optical flow sampling process, we cropped the image to a 174 × 174 × 174 window and placed a 4 × 4 × 4 lattice of equispaced points centred inside it. These 64 lattice points had fixed relative spatial positions. Based on pairwise distances, we created a (4 × 4 × 4)-by-(4 × 4 × 4) covariance matrix to describe how correlated distortions should be in the warping. We applied a Gaussian kernel with a width of 24 voxels. These 3 × 64 values were multiplied by a random scaling chosen uniformly in [0, 4], treated as optical flow values and applied to the image in the distortion along three spatial directions for each of the 64 lattice points. Next, we extrapolated optical flow values to each underlying voxel position with a polyharmonic spline and applied the warp by resampling the image at each voxel with its own floating point offsets in 3D. From the centre of the warped and resampled image, we cropped a 96 × 96 × 96 patch and used this as training data. When interpolating supervision segmentation masks, we converted the masks to floating-point probabilities and applied clipping heuristics after the warp and resampling to ensure that probabilities were valid. Finally, we obtained volume measurements by thresholding the model output, removing disconnected structures, and multiplying the number of mask voxels by the image resolution.

Quality control consisted of iterations of visual inspection of extreme volumes for each distinct organ/structure, as well as spot checks of hundreds of random subjects. The training data was regularly enriched to include problematic cases. We repeated this procedure and retrained the model until the results did not display outliers for extreme subjects nor any of the random spot checks. Further details on the model architecture are available in [6].

### **Data Preparation**

Individual fields (variables or measurements) in the UK Biobank are organized into categories. Here we provide details of the procedure used to select a set of data fields that will be used in the prediction models. All categories were initially considered for inclusion in predicting image-derived phenotypes (IDPs). Specific categories were excluded based on the following criteria:

- Disease-based categories
- Gender-specific categories
- UK Biobank “operations” categories
- Genetics categories

There were a few categories that were empty and excluded. A total of 102 categories were kept and all fields included in these categories were considered in the next stage.

Each data field in the UK Biobank was measured on at least one, and possibly multiple, time points known as instances. Data fields where the description included the text “device” or “method”, or “pilot” or “reason” were discarded. Data fields where the data type was “date” or “text”, or “time” were also discarded. Some data fields in the UK Biobank contain more than one column of information known as an array. For the remaining data fields, each column in the array was assessed. If the baseline instance (zero) existed and in contained measurements for greater than 50% of the total number of participants (approximately 500,000), then if the imaging instance (two) existed and there were greater than 50% of the number of imaging participants, then the imaging instance was kept else the baseline instance was kept. For example, sex was recorded at the baseline visit but not recorded in later visits, so the baseline instance is retained whereas weight was measured at both instances, so the value recorded at the imaging visit is retained. A total of 4261 unique variables were processed with only 449 surviving the selection criteria outlined above.

Variables with missing values were filled in using imputation by chained random forests in the missRanger package [1-3] . The correlation matrix was computed between all continuous variables, where variables were discarded using a cutoff of 0.9 in the caret package [4]. A total of 382 variables were kept after the correlation-based pruning procedure.

### **Predictive Models**

Four models were assessed in the prediction of IDPs: linear regression, gradient boosting, random forest and support vector machine (SVM). The data were split into training and test datasets, where the training data were all participants used in [5]. (N_{train} = 37,705) and the test data were participants in the UK Biobank imaging cohort who had been processed afterwards (N_{test} = 5,258). This is an approximate 88/12 split.

Variable selection, against each IDP, was performed on the training data using the Boruta algorithm [6] in the Pomona package [7]. The remaining variables (**Supplementary table 2**) were included in the predictive models. Ten-fold cross validation was performed to find the optimal parameter settings. Model performance was assessed using the R-squared statistic, root mean squared error (RMSE) and mean absolute error (MAE) on the out-of-sample test data. In almost all IDPs the SVM model was superior except for spleen and kidneys, where the gradient boosting model performed better in the R-squared statistic and RMSE. However, the difference between the two models in these performance metrics was less than 1% and, therefore, the SVM model was used to predict all IDPs.

**Supplementary figures**

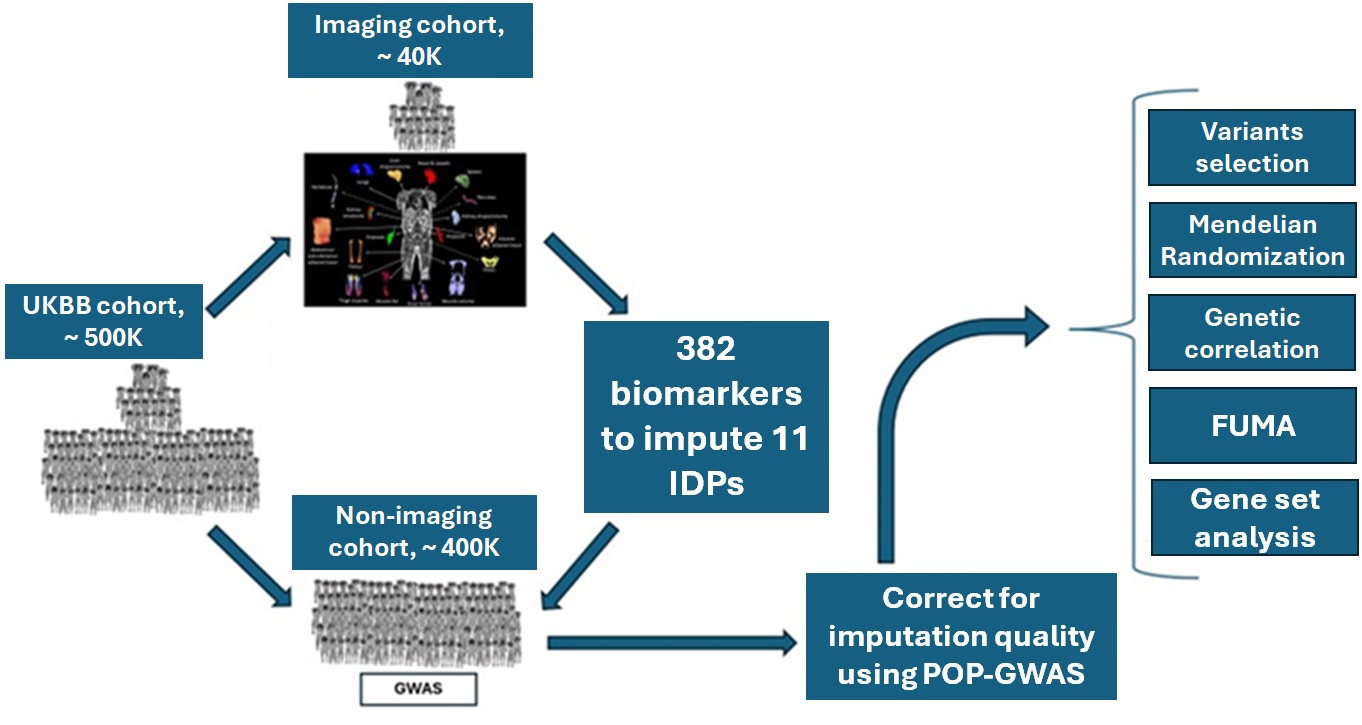

**Supplementary figure 1. Study Design.** We applied a machine learning-assisted GWAS framework to investigate the genetic architecture of organ volumes and their links to disease. Organ and tissue volumes were measured in 37,589 UK Biobank participants with abdominal MRI scans (imaging cohort). Using up to 382 routinely collected biomarkers, we imputed nine imaging-derived phenotypes (IDPs) in the remaining 370,568 participants without MRI data (non-imaging cohort), substantially increasing the effective sample size and statistical power. GWAS was performed on the imputed traits, and the POP-GWAS method was applied to correct for bias introduced by phenotype imputation, ensuring robust and well-calibrated association estimates. Downstream analyses included heritability estimation, genetic correlation with complex traits, functional annotation, and Mendelian randomisation to prioritise causal pathways linking organ size to disease risk.

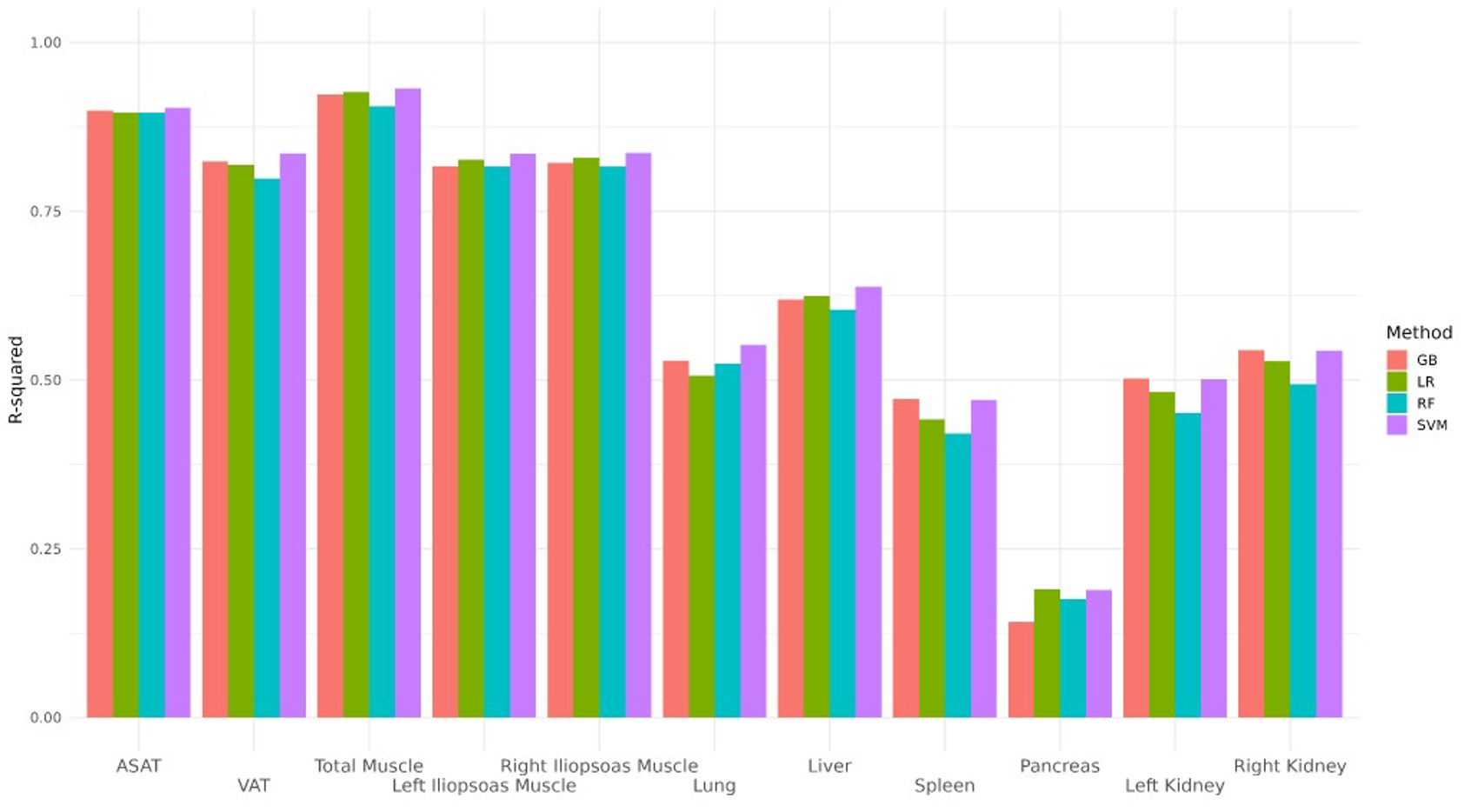

**Supplementary figure 2. Predictive performance of machine learning models for imaging-derived phenotypes (IDPs).**
R-squared values are shown for four predictive modelling approaches—linear regression (LR), random forest (RF), gradient boosting (GB), and support vector machine (SVM)—evaluated on out-of-sample test data (N = 5,258). Performance is reported across nine IDPs, including abdominal subcutaneous fat, visceral fat, liver, pancreas, lungs, spleen, total muscle volume, and left and right iliopsoas muscle volumes. Kidney volumes (left and right) and iliopsoas muscles were modelled individually. SVM yielded the highest predictive accuracy for most traits, with GB performing slightly better for spleen and kidney volumes.

(a).

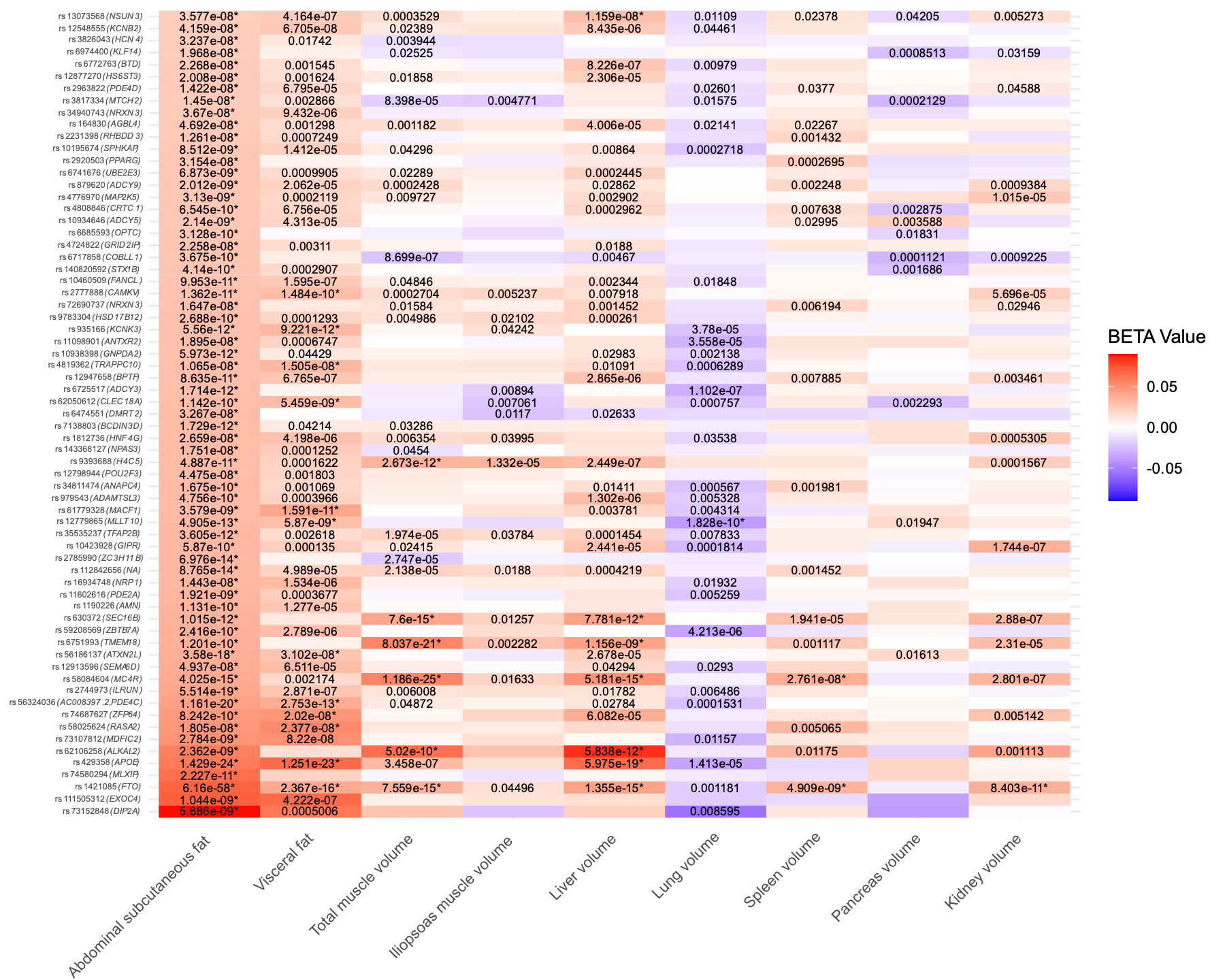

(b).

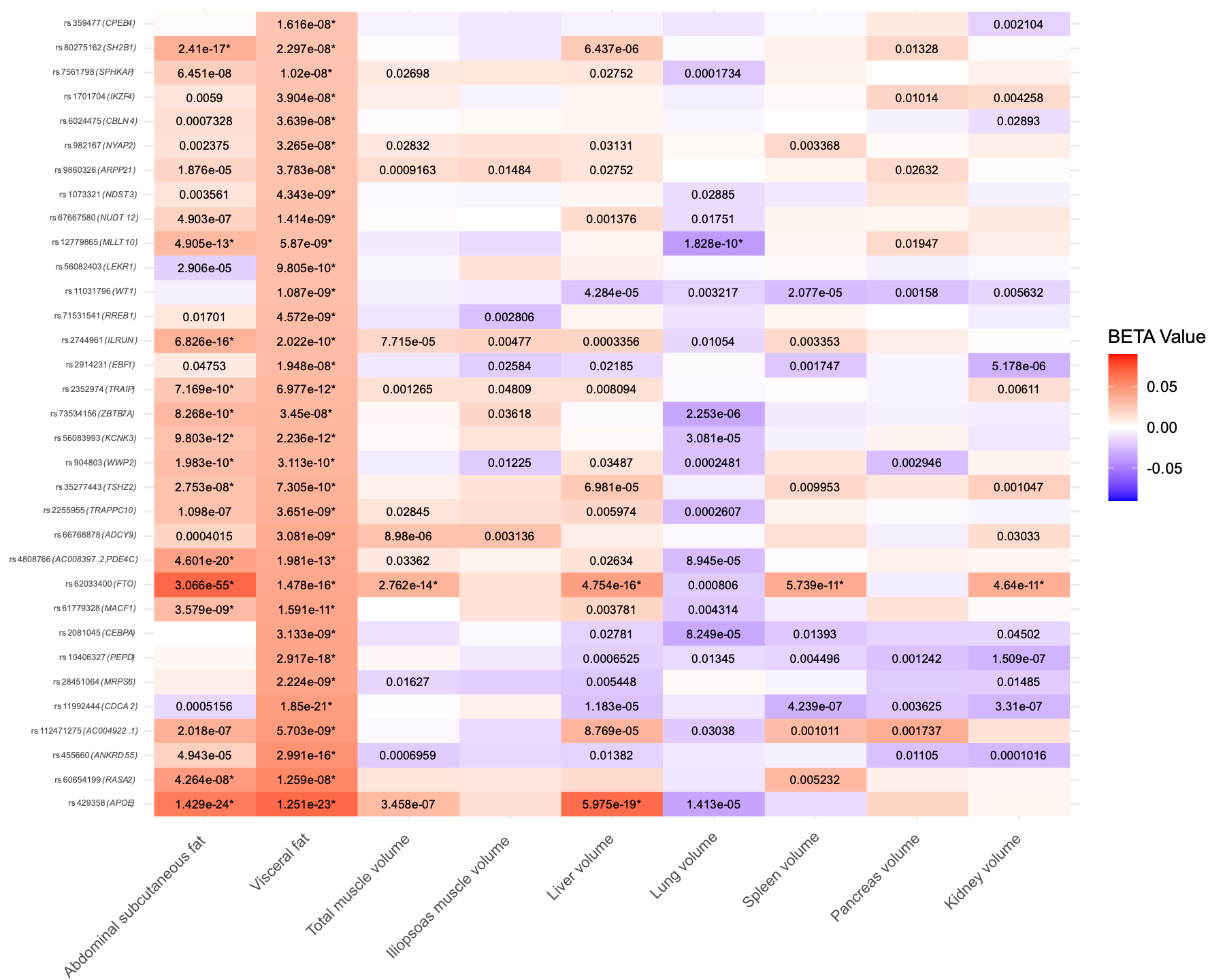

(c).

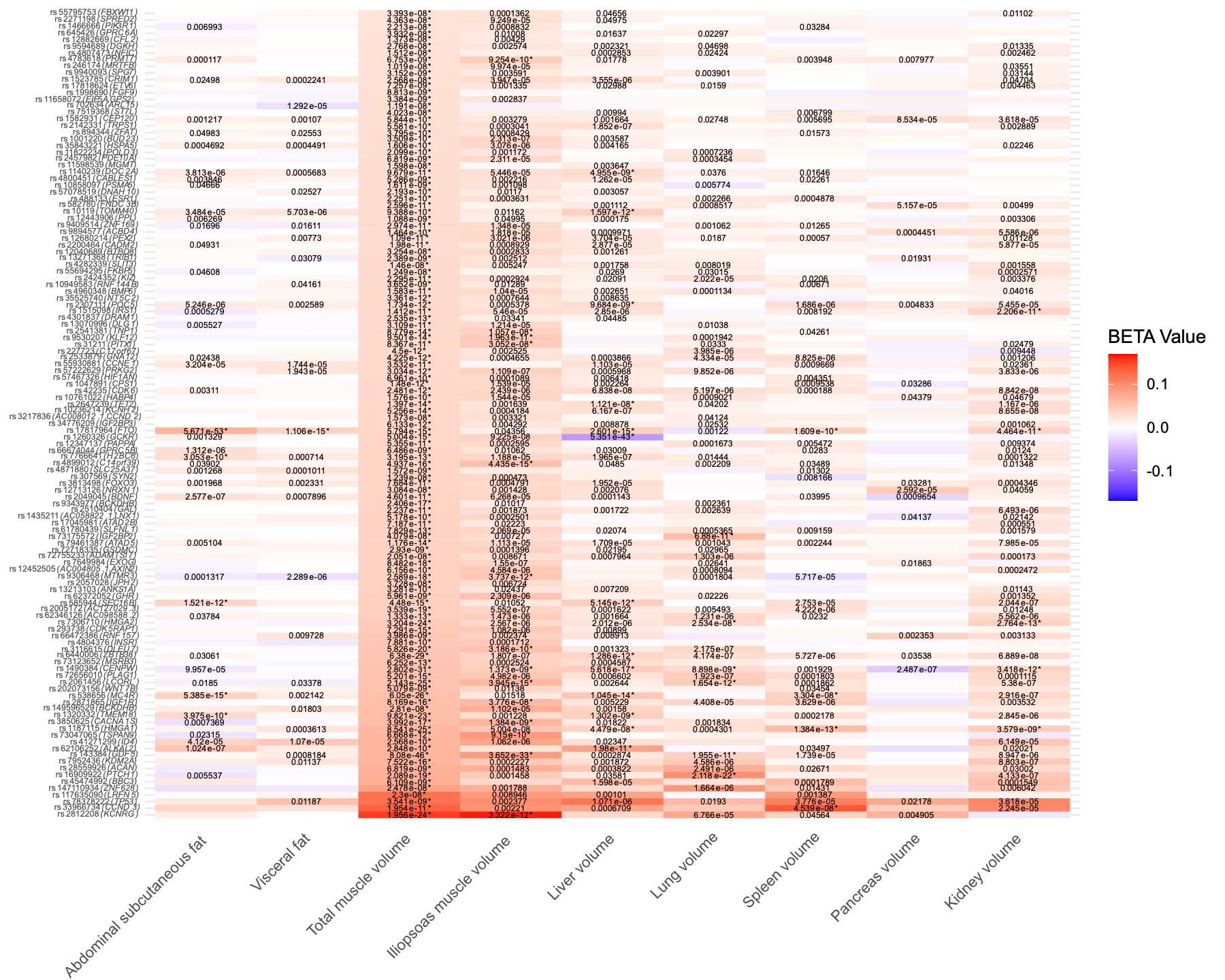

(d).

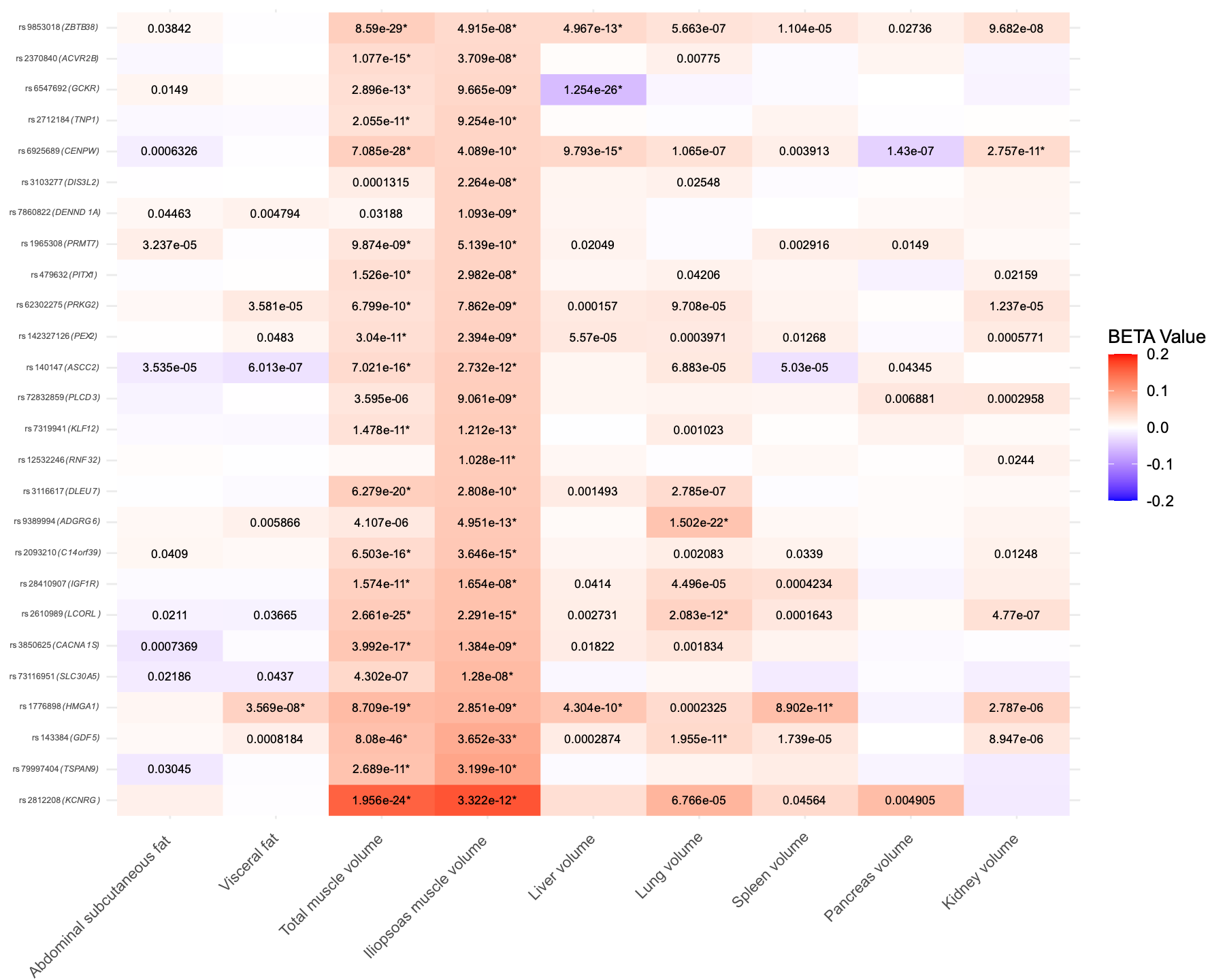

(e).

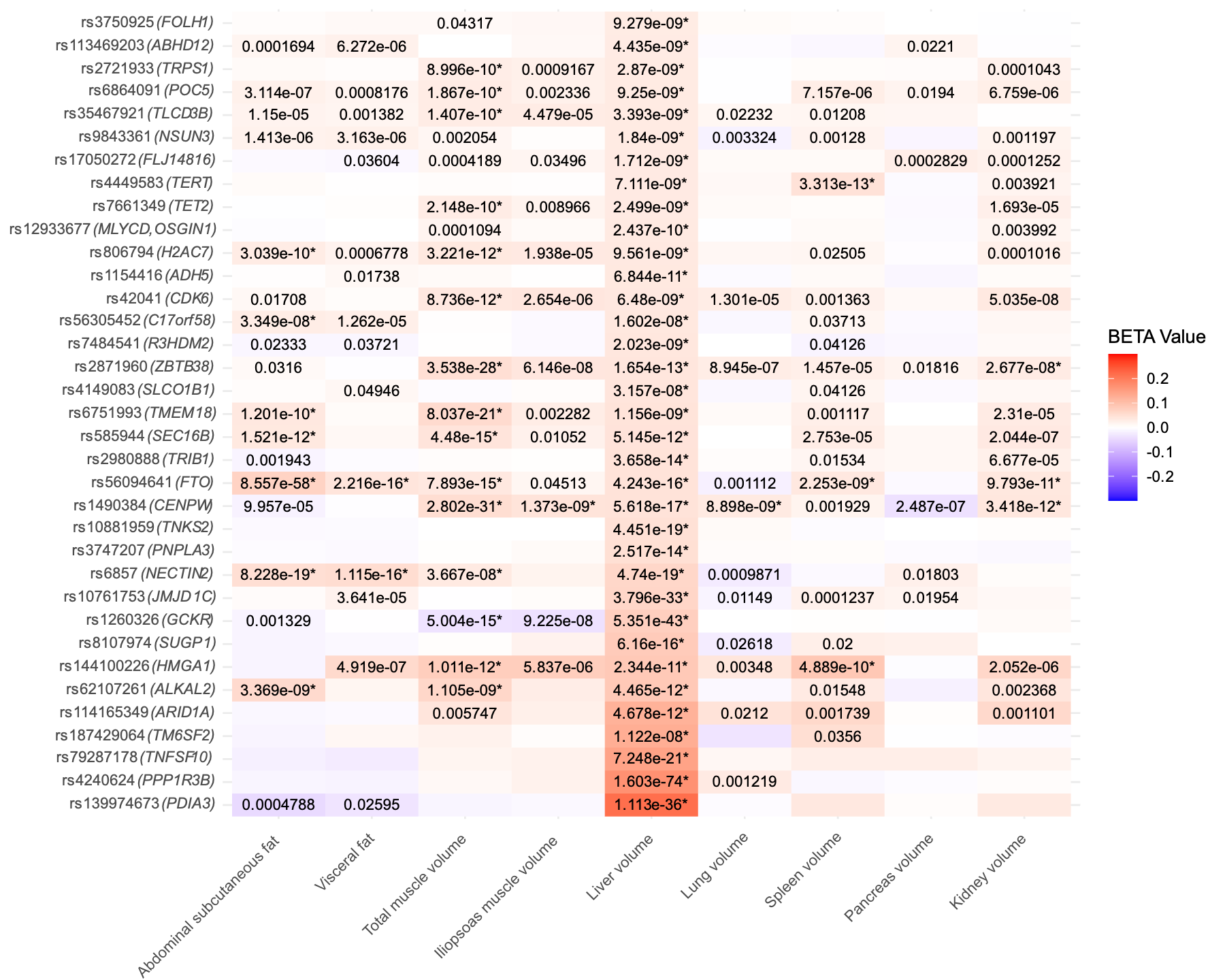

(f).

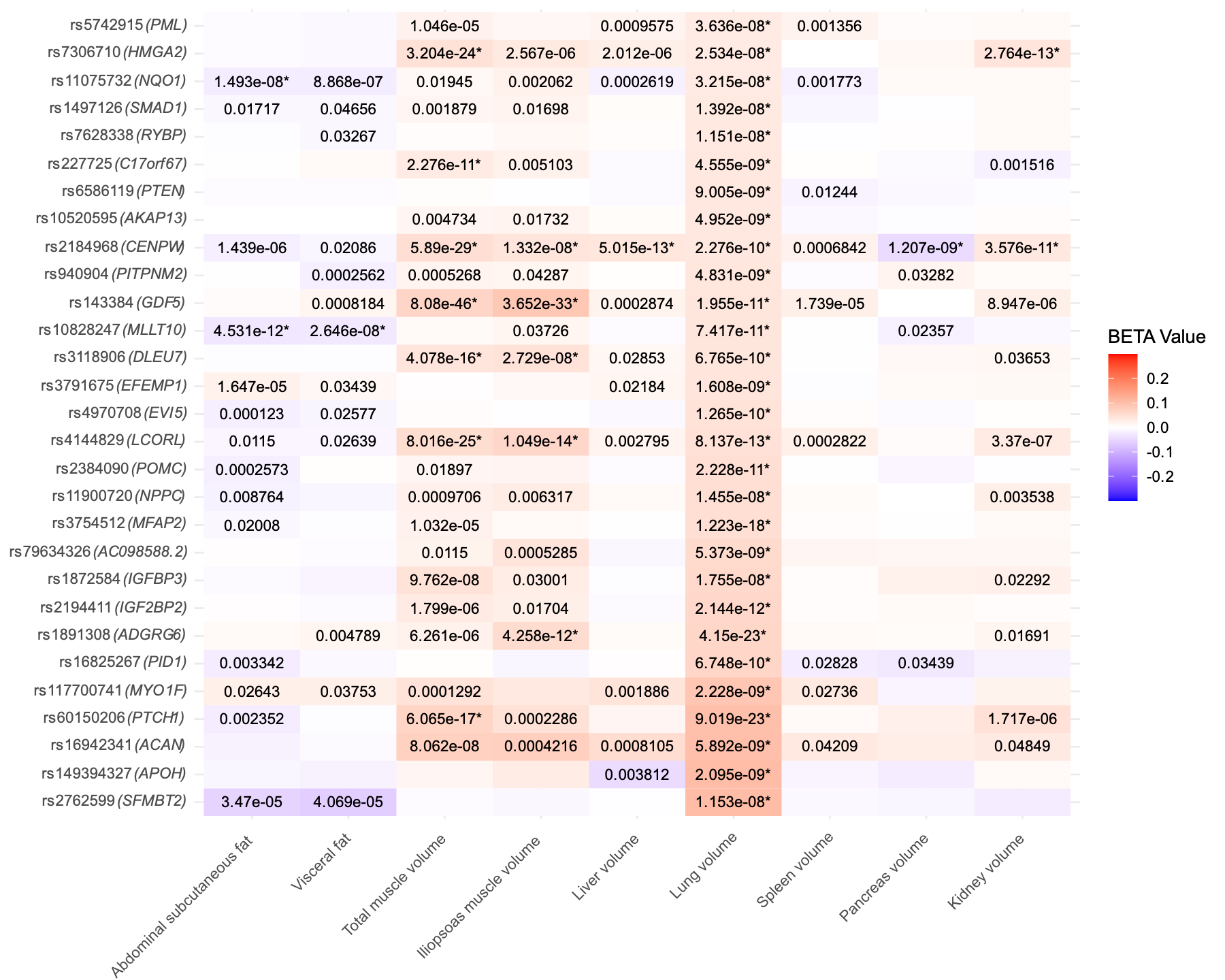

(g).

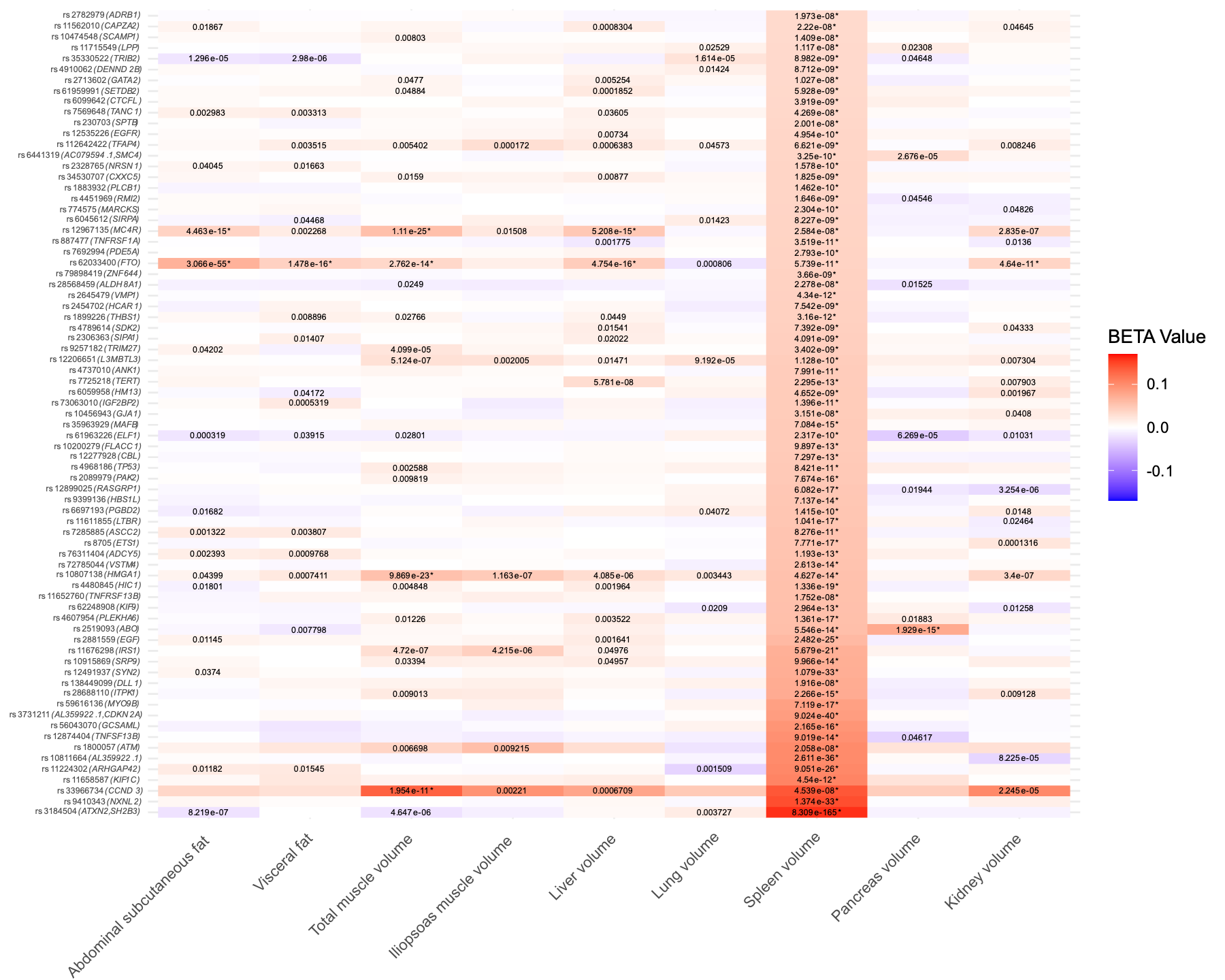

(h).

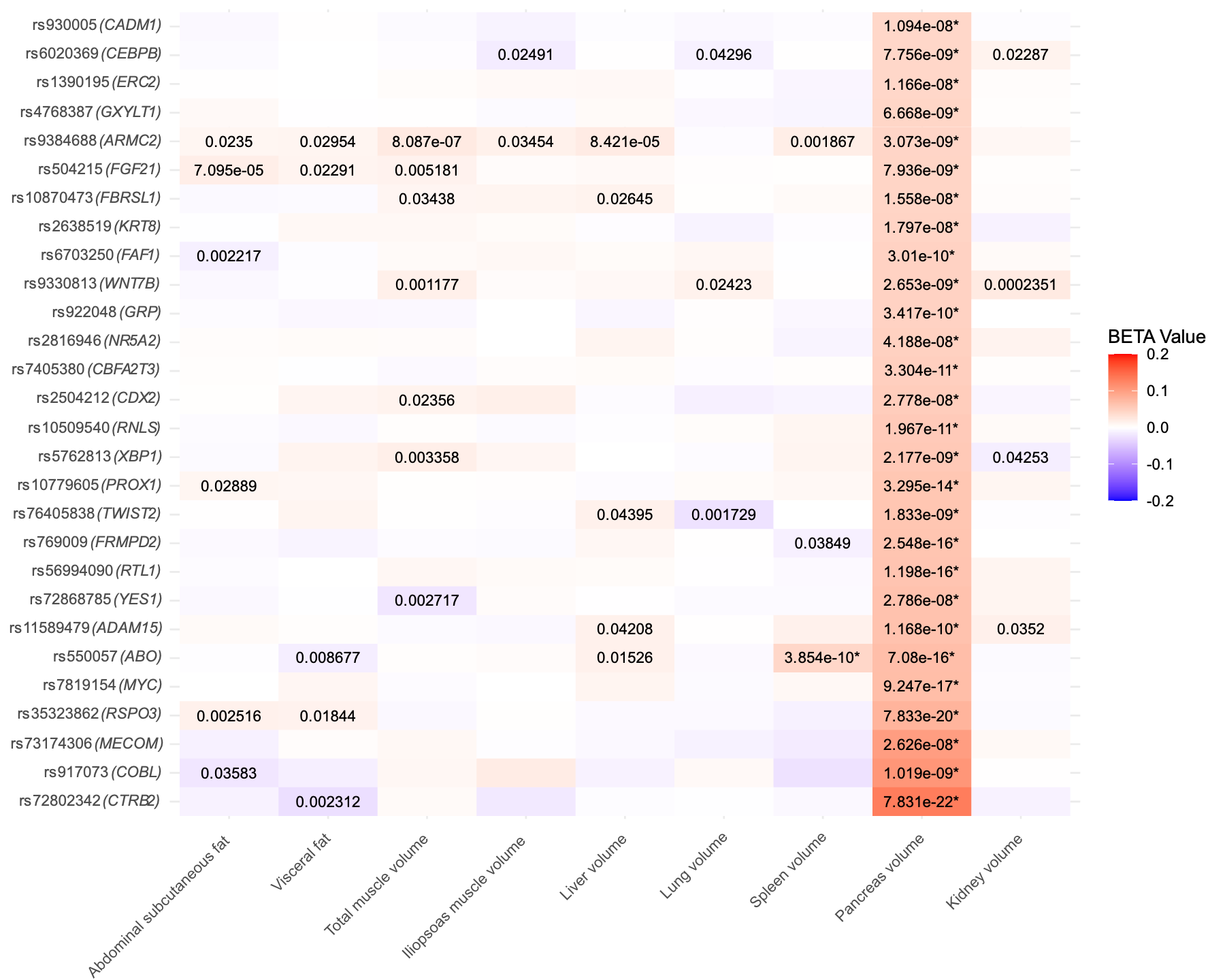

(i).

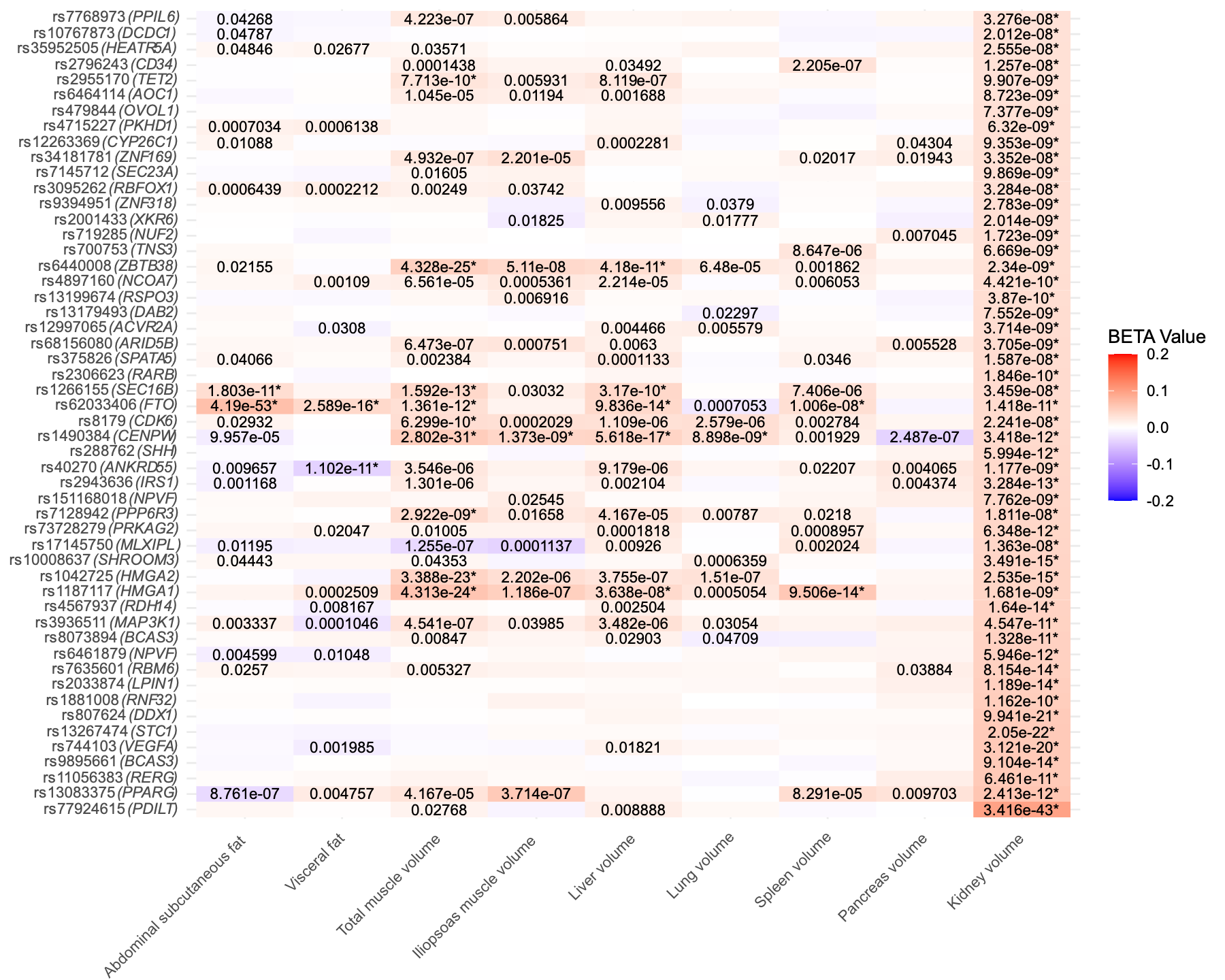

**Supplementary Figure 3. Cross-trait effects of genome-wide significant variants across nine imaged-derived phenotypes (IDPs).**
Each panel (a–i) corresponds to one IDP and displays the effects of its genome-wide significant variants across all nine IDPs. The x-axis shows independent genome-wide significant variants, annotated by their nearest gene. The y-axis lists the nine IDPs. Each cell represents the effect of increasing allele dosage (β) of a given variant on the corresponding IDP. Colours indicate the direction of effect (red = positive β, blue = negative β), and cell values report the raw p-values. Asterisks (*) denote associations surpassing the genome-wide significance threshold (p < 5×10⁻⁸). Panels: (a) Abdominal subcutaneous fat, (b) Visceral fat, (c) Total muscle volume, (d) Iliopsoas muscle volume, (e) Liver volume, (f) Lung volume, (g) Spleen volume, (h) Pancreas volume, (i) Kidney volume.

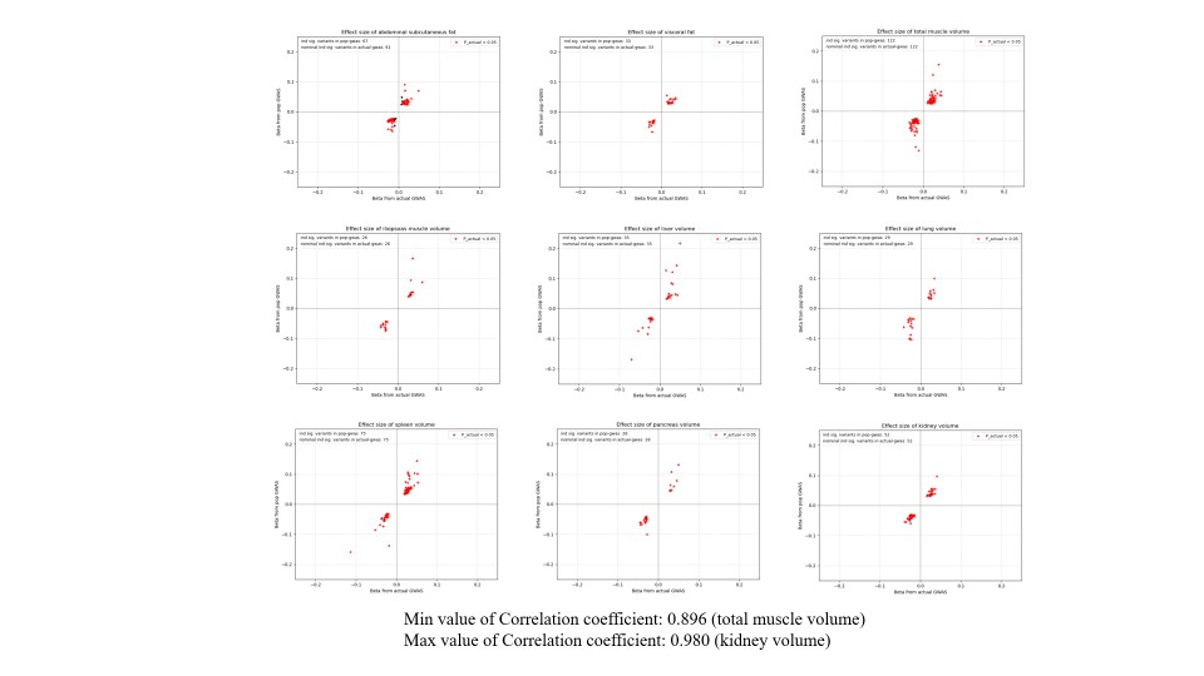

**Supplementary Figure 4. Concordance of effect sizes between GWAS of MRI-derived IDPs and POP-GWAS of imputed IDPs.**
Scatter plots compare SNP effect sizes from GWAS of MRI-derived IDPs in the imaging cohort (x-axis: β from imaging cohort GWAS) and POP-GWAS of imputed IDPs in the non-imaging cohort (y-axis: β from POP-GWAS). Each point represents a genome-wide significant variant.

(a).

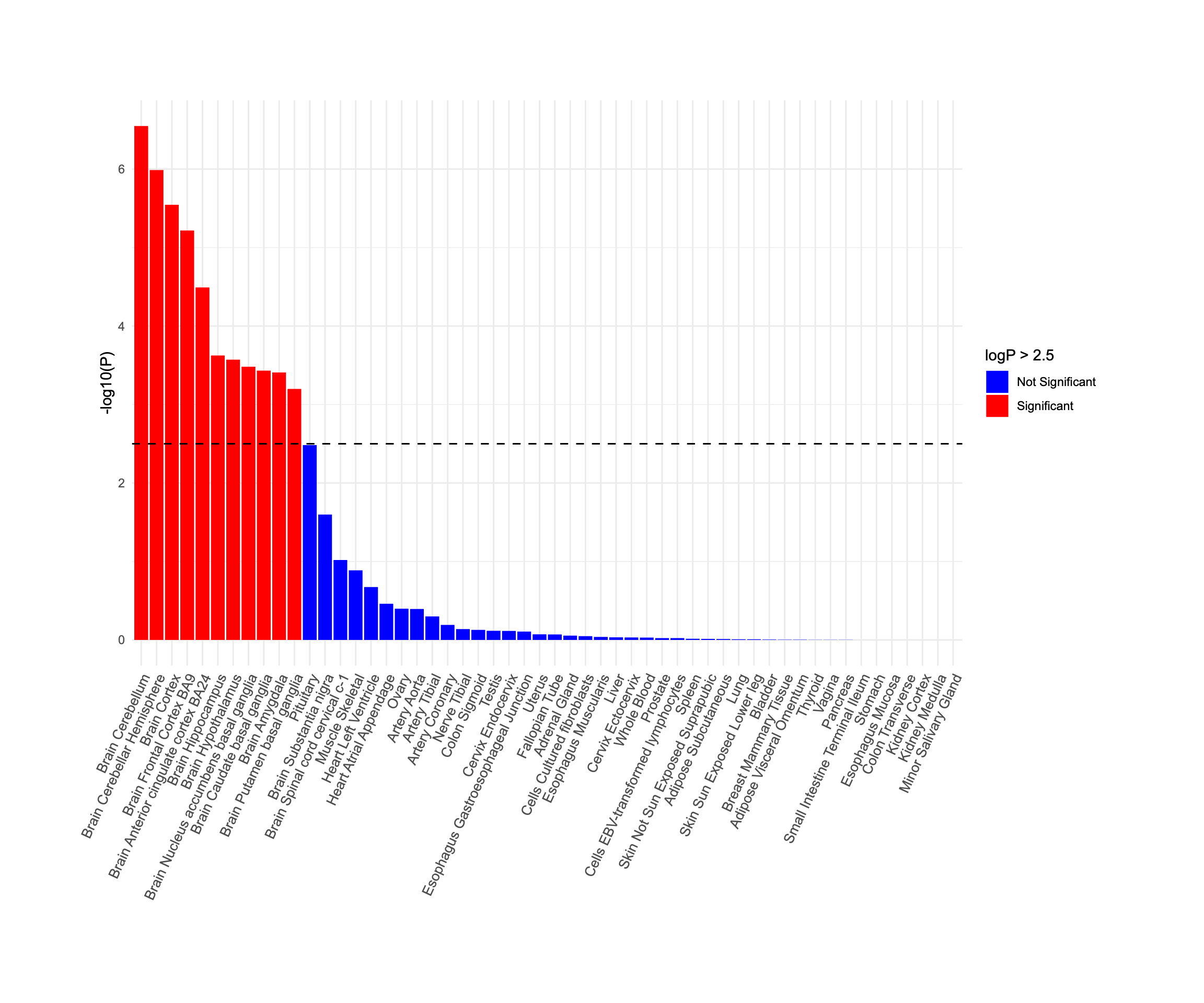

(b).

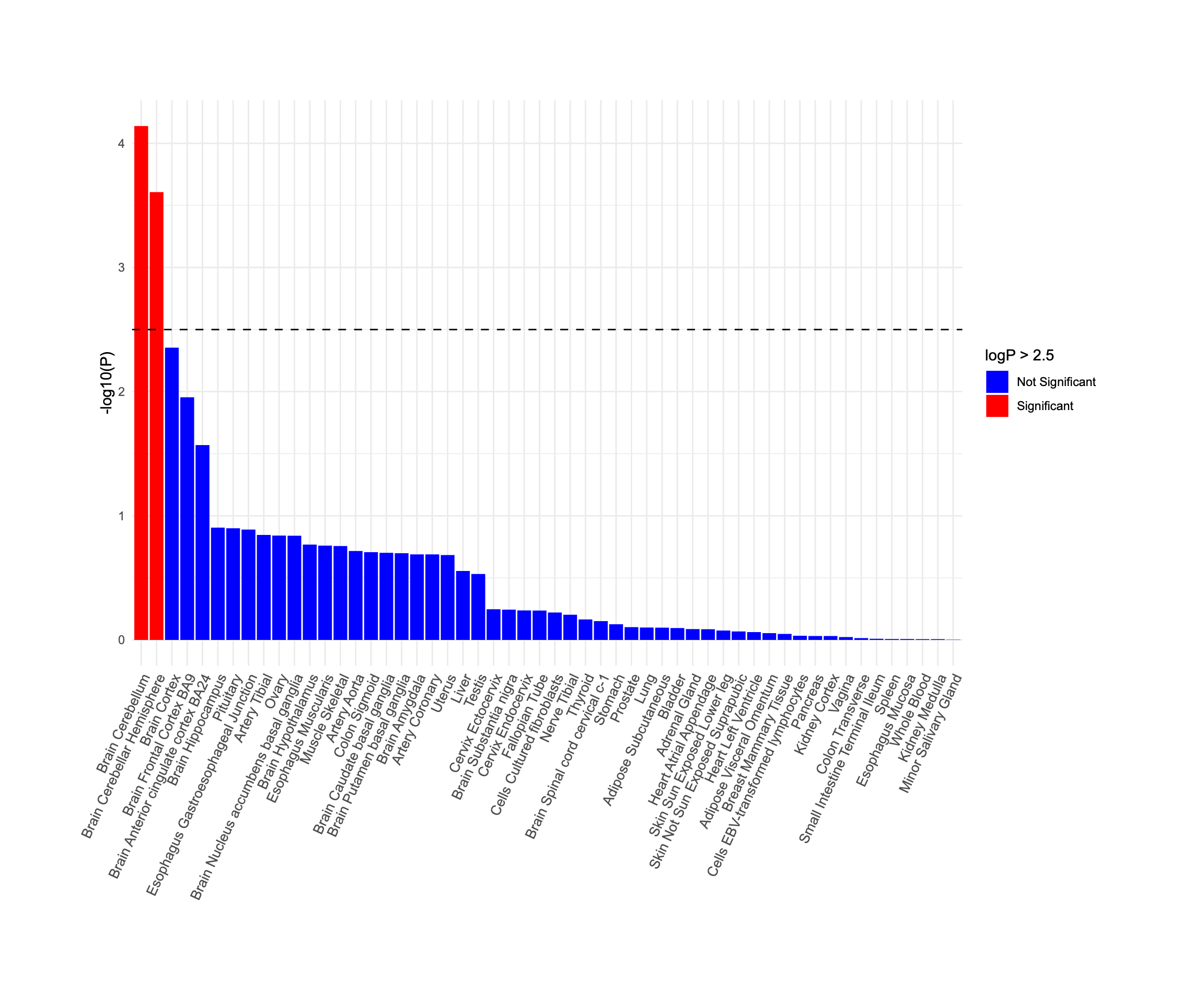

(c).

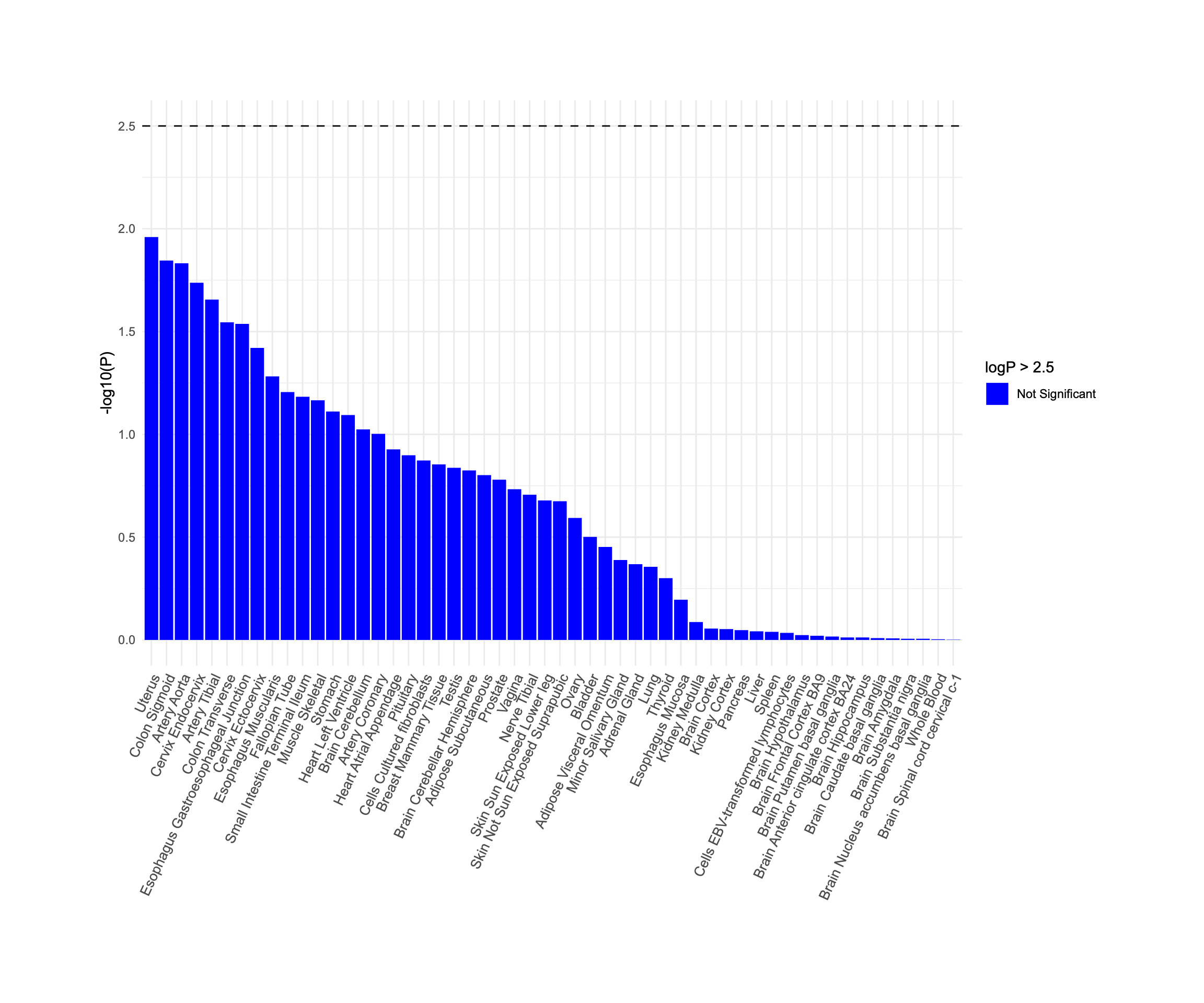

(d).

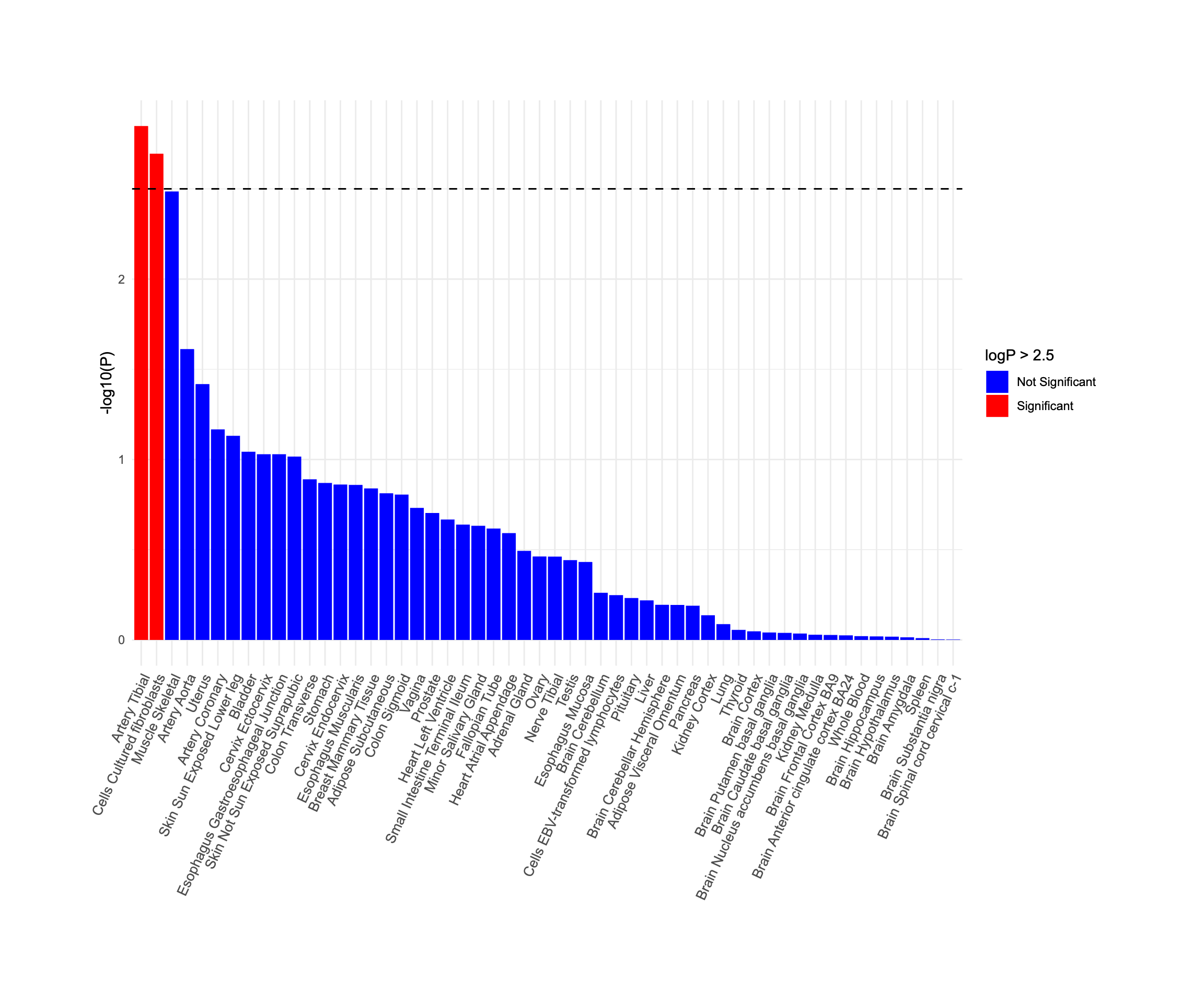

(e).

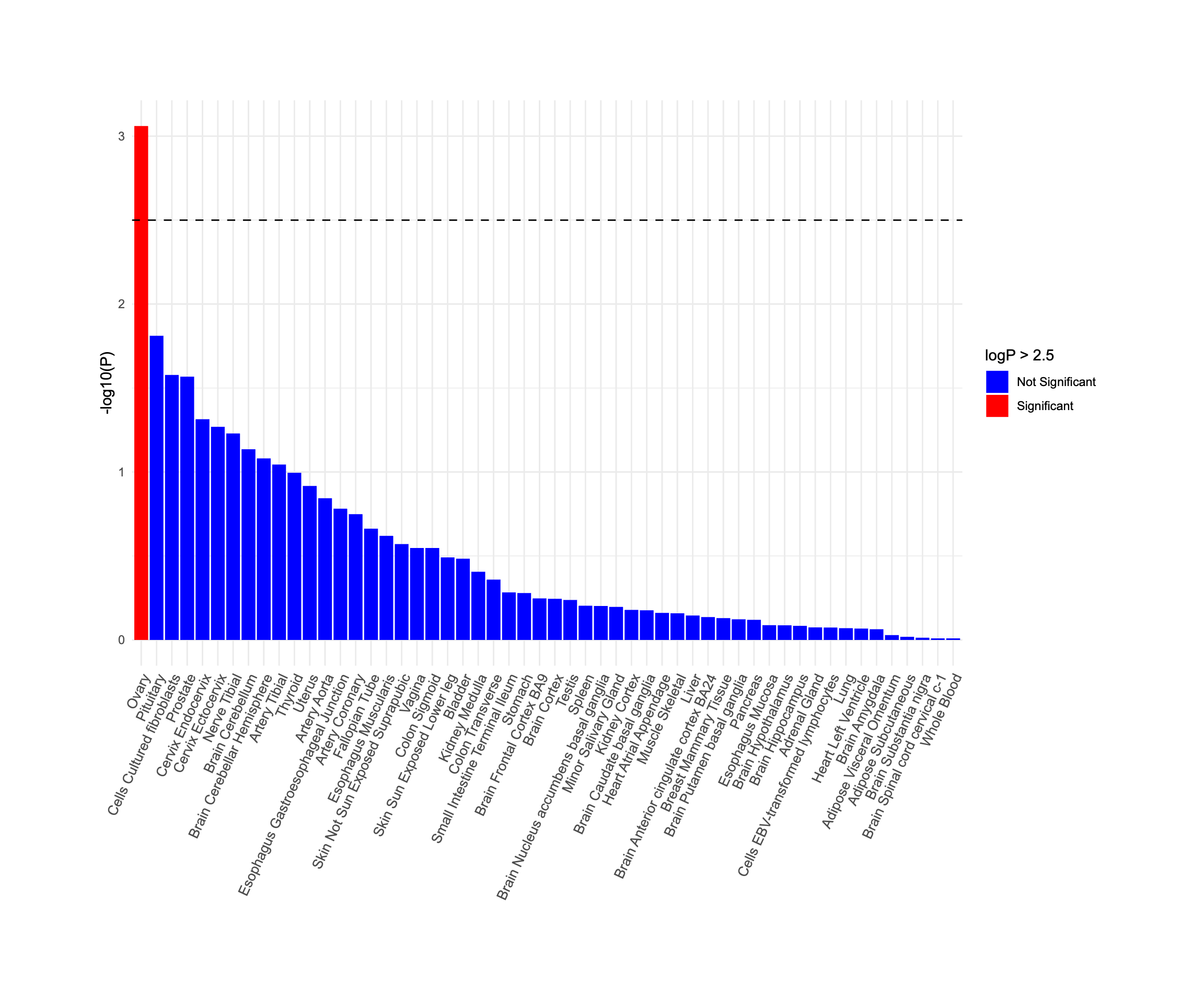

(f).

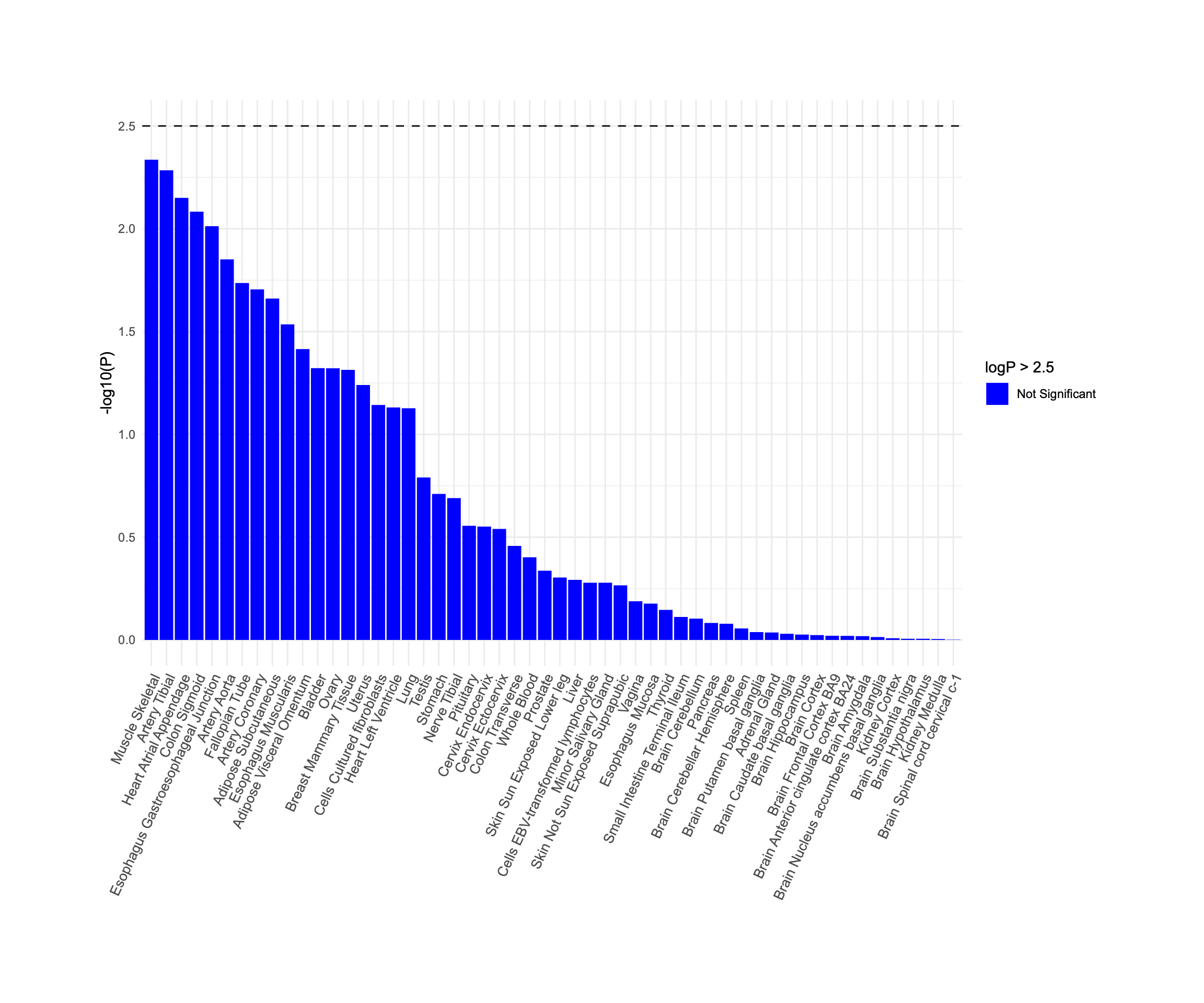

(g).

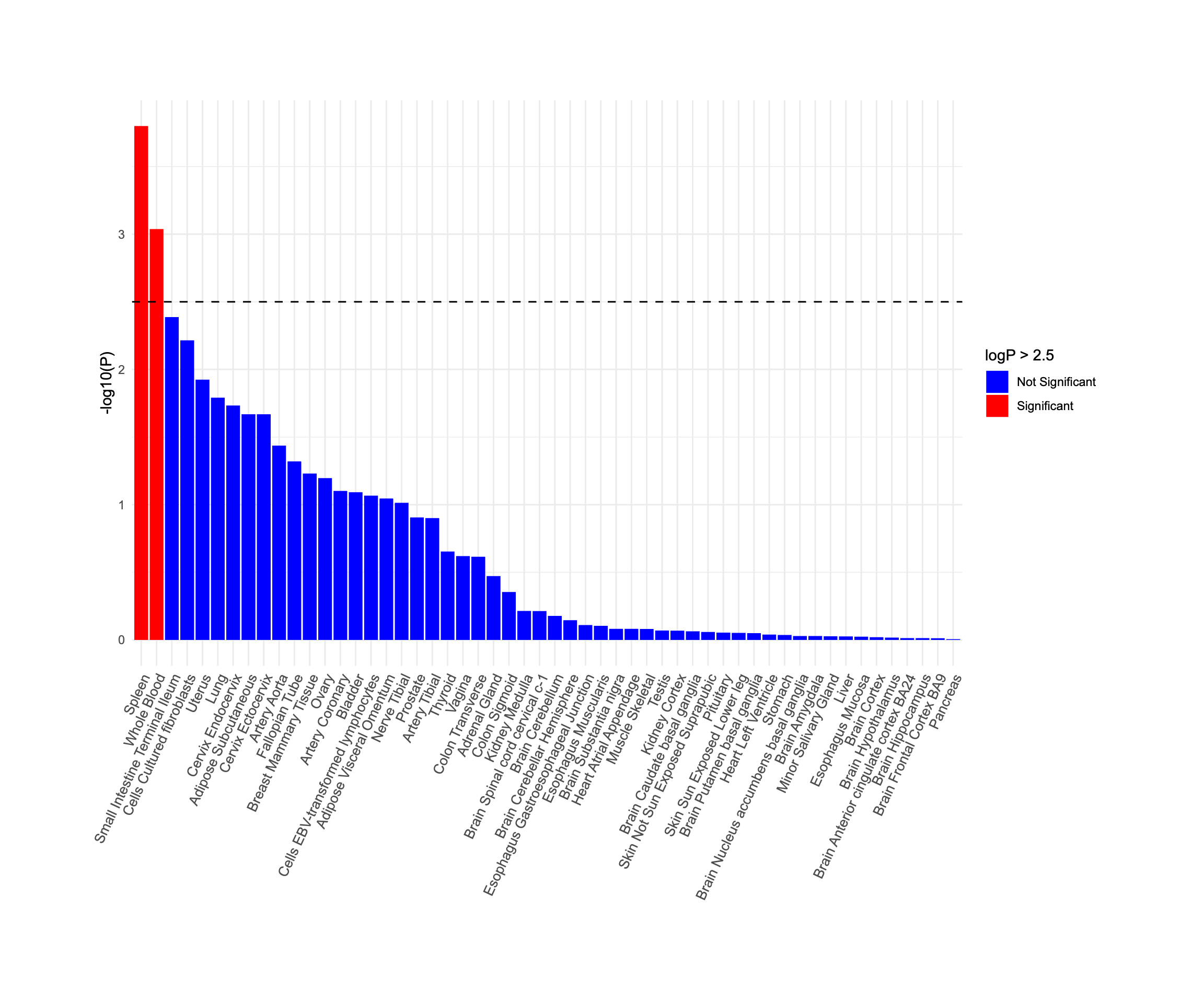

(h).

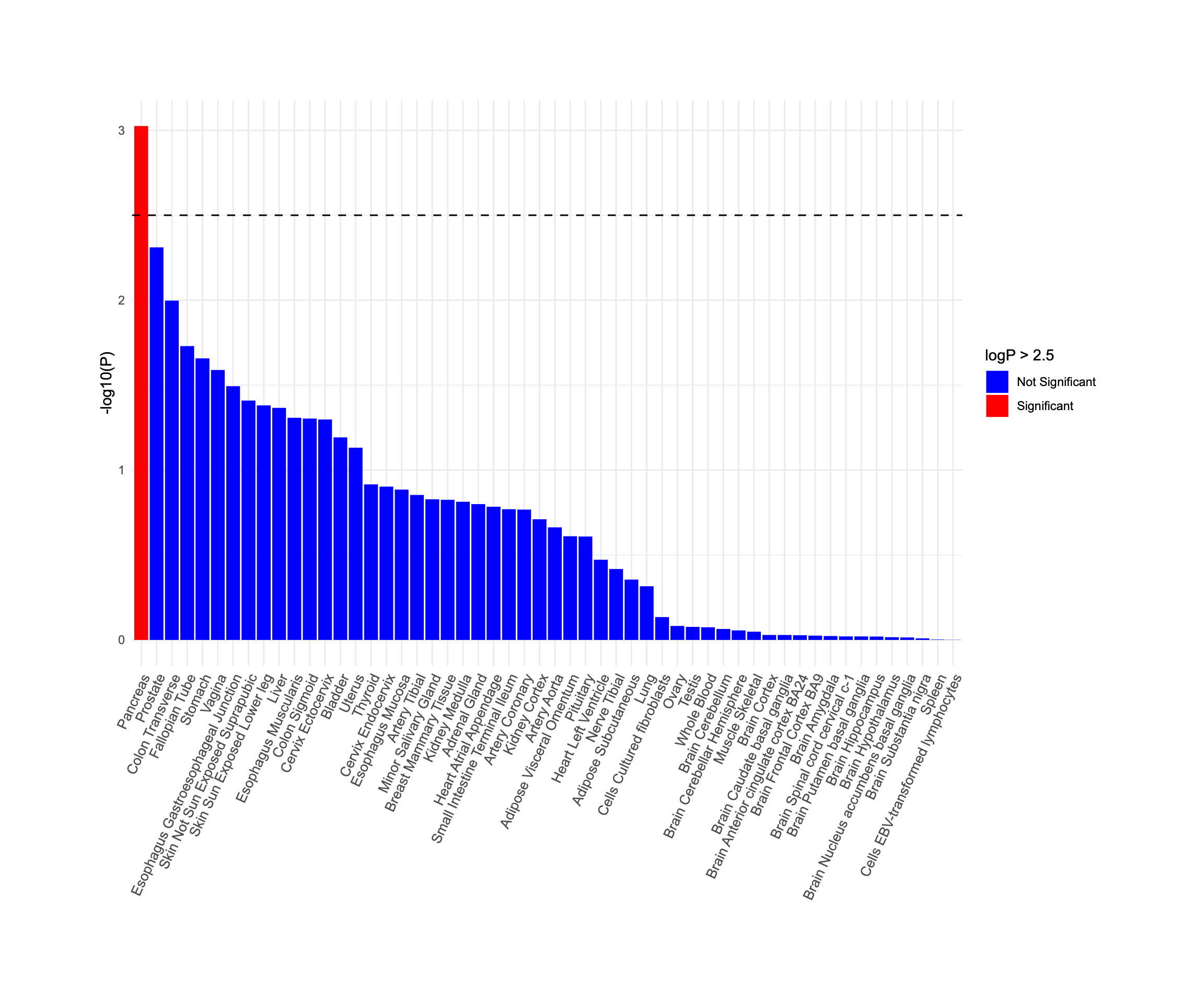

(i)

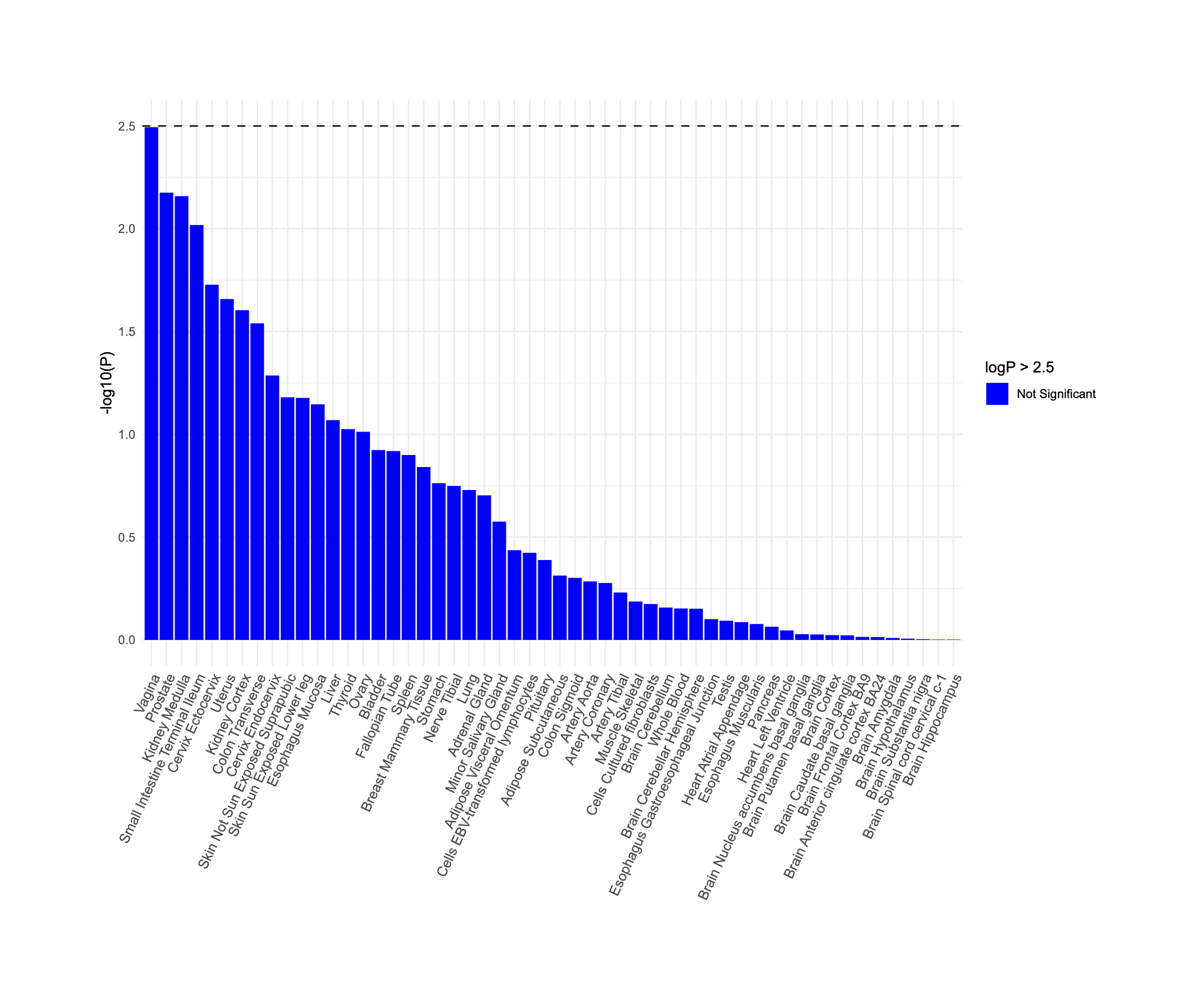

**Supplementary Figure 5. Tissue expression enrichment for image-derived phenotypes (IDPs).**
Bar plots show tissue-specific expression profiles based on MAGMA gene-property analysis using GTEx v8 transcriptomic data across 53 tissue types. The x-axis indicates tissue types, and the y-axis shows the –log₁₀(p-values) of enrichment. A significance threshold of –log₁₀(p) > 2.5 (equivalent to p < 0.0032) was used to identify significantly enriched tissues. Bars are coloured red for significant associations and blue for non-significant. Panels represent the following IDPs: (a) Abdominal subcutaneous fat; (b) Visceral fat; (c) Total muscle volume; (d) Iliopsoas muscle volume; (e) Liver volume; (f) Lung volume; (g) Spleen volume; (h) Pancreas volume; (i) Kidney volume.

**Supplementary tables**

| **Supplementary table 1:** Demographics and baseline health metrics of study population | | | |
| --- | --- | --- | --- |
|  | **Imaging cohort** | **Train Set cohort** | **Test Set cohort** |
| No. of participants (%female) | 42.963 (51.6) | 37,705 (51.8) | 5,258 (50.4) |
| Age (**years**) (Mean±SD) | 64.18 ± 7.72 | 63.68 ± 7.55 | 67.71 ± 8.02 |
| BMI (**kg/m²)** Mean±SD) | 26.5 ± 4.36 | 26.5 ± 4.39 | 26.3 ± 4.16 |
| Abdominal Subcutaneous fat (L) (Mean±SD) | 8.47 ± 4.12 | 8.51 ± 4.16 | 8.17 ± 3.87 |
| Visceral fat (L) (Mean±SD) | 3.95 ± 2.31 | 3.95 ± 2.31 | 3.96 ± 2.30 |
| Liver volume (L) (Mean±SD) | 1.41 ± 0. | 1.42 ± 0.52 | 1.38 ± 0.49 |
| Lung volume (L) (Mean±SD) | 2.60 ± 0.73 | 2.60 ± 0.73 | 2.59 ± 0.73 |
| Kidney volume left (mL) (Mean±SD) | 134.5 ± 31.6 | 135.1 ± 31.5 | 130.4 ± 31.7 |
| Kidney volume right (mL) (Mean±SD) | 135.1 ± 30.3 | 135.5 ± 30.3 | 132.5 ± 29.7 |
| Pancreas volume (mL) (Mean±SD) | 58.8 ± 17.7 | 59.1 ± 17.7 | 56.9 ± 17.8 |
| Spleen volume (mL) (Mean±SD) | 165.7 ± 70.7 | 166.6 ± 71.1 | 158.7 ± 67.7 |
| Total muscle volume (L) (Mean±SD) | 17.81 ± 4.62 | 17.86 ± 4.64 | 17.38 ± 4.47 |
| Iliopsoas muscle volume left (mL) (Mean±SD) | 314.2 ± 85.9 | 315.3 ± 86.2 | 306.7 ± 83.1 |
| Iliopsoas muscle volume right (mL) (Mean±SD) | 327.0 ± 86.4 | 328.1 ± 86.7 | 319.1 ± 83.9 |

**Supplementary table 2:** Overview of imaging-derived phenotypes (IDPs) and the number of predictive features used in model training. The table reports the number of input variables retained for each organ or tissue after applying the Boruta feature selection algorithm, based on an initial set of 382 candidate biomarkers spanning anthropometry, clinical biochemistry, and lifestyle traits. For the kidney and iliopsoas muscle volume, the left and right sides were merged by calculating their average.

| **IDPs** | **Number of features** | **UK Biobank**  **Field number** |
| --- | --- | --- |
| Abdominal subcutaneous fat | 101 | 21002, 22033, 22039, 26414, 30600, 30610, 30620, 30630, 30640, 30650, 30660, 30700, 30710, 30720, 30730, 30770, 30830, 30840, 30850, 30870, 30880, 30890, 23106, 23108, 23112, 23113, 23124, 23127, 48, 49, 50, 51, 20015, 21001, 102, 4079, 4079, 4080, 4080, 3143, 47, 3062, 3062, 3062, 3063, 3063, 3064, 3064, 3064, 20150, 20256, 20257, 20258, 1588, 1598, 1438, 1458, 1070, 864, 884, 904, 914, 2139, 845, 767, 137, 30000, 30010, 30030, 30040, 30050, 30080, 30090, 30120, 30140, 30190, 30240, 30270, 30280, 30290, 30510, 30530, 34, 22035, 1697, 2178, 2306, 6150, 6154, 6159, 1558, 1349, 1548, 2237, 981, 924, 6164, 2080, 6142, 31 |
| Visceral fat | 90 | 21002, 22039, 30610, 30620, 30630, 30640, 30650, 30660, 30690, 30700, 30710, 30720, 30730, 30740, 30750, 30770, 30810, 30830, 30850, 30870, 30880, 23106, 23108, 23112, 23113, 23124, 23127, 48, 49, 50, 51, 20015, 21001, 102, 4079, 4079, 4080, 4080, 3143, 47, 3062, 3062, 3062, 3063, 3063, 3064, 3064, 3064, 20150, 20256, 20257, 1588, 1309, 1070, 904, 914, 845, 137, 30000, 30010, 30030, 30130, 30140, 30240, 30280, 30290, 30510, 30520, 30530, 34, 1687, 1835, 2316, 2178, 2188, 2306, 2443, 6150, 2492, 6153, 1349, 1448, 2237, 924, 6164, 1210, 6138, 6142, 31 |
| Total Muscle volume | 97 | 21002, 22033, 22034, 22039, 30610, 30620, 30630, 30650, 30660, 30700, 30710, 30720, 30730, 30770, 30810, 30830, 30840, 30850, 30870, 30880, 23106, 23108, 23112, 23113, 23124, 23127, 48, 49, 50, 51, 20015, 21001, 102, 4079, 4079, 4080, 4080, 78, 3143, 47, 3062, 3062, 3062, 3063, 3063, 3064, 3064, 3064, 20150, 20256, 20257, 400, 404, 404, 404, 20023, 1588, 1438, 1070, 904, 914, 2139, 2149, 767, 699, 709, 20022, 137, 20011, 30010, 30030, 30090, 30240, 30290, 30510, 30530, 34, 22032, 22035, 3077, 1687, 1697, 1797, 1835, 2345, 6153, 1349, 1100, 924, 6164, 6160, 6142, 680, 728, 738, 31 |
| Left Iliopsoas Muscle volume | 90 | 21002, 22039, 30600, 30620, 30630, 30650, 30660, 30700, 30710, 30720, 30730, 30770, 30810, 30830, 30840, 30850, 30870, 30880, 23106, 23108, 23112, 23113, 23124, 23127, 48, 49, 50, 51, 20015, 21001, 102, 4079, 4079, 4080, 4080, 78, 3143, 47, 3062, 3062, 3062, 3063, 3063, 3064, 3064, 3064, 20150, 20256, 20257, 400, 400, 404, 404, 404, 20023, 1588, 1438, 904, 914, 2139, 2149, 767, 699, 709, 20022, 137, 20011, 30010, 30030, 30080, 30090, 30190, 30510, 30530, 34, 3077, 1697, 1797, 1835, 2345, 6153, 1349, 1100, 6164, 6160, 6142, 680, 738, 31 |
| Right Iliopsoas Muscle volume | 91 | 21002, 22039, 30600, 30620, 30630, 30650, 30660, 30700, 30710, 30720, 30730, 30770, 30810, 30830, 30840, 30850, 30870, 30880, 23106, 23108, 23112, 23113, 23124, 23127, 48, 49, 50, 51, 20015, 21001, 102, 4079, 4079, 4080, 4080, 78, 3143, 47, 3062, 3062, 3062, 3063, 3063, 3064, 3064, 3064, 20150, 20256, 20257, 400, 400, 404, 404, 404, 404, 20023, 3526, 1588, 1438, 904, 914, 2139, 2149, 767, 699, 709, 20022, 137, 20011, 30010, 30030, 30080, 30090, 30510, 30530, 34, 3077, 1697, 1797, 1835, 2345, 2492, 6153, 1349, 6164, 6160, 6142, 680, 738, 31 |
| Liver volume | 76 | 21002, 30600, 30620, 30630, 30650, 30700, 30710, 30730, 30740, 30750, 30830, 30850, 30860, 30870, 30880, 23106, 23108, 23112, 23113, 23124, 23127, 48, 49, 50, 51, 20015, 21001, 102, 4079, 4079, 3143, 47, 3062, 3062, 3062, 3063, 3063, 3064, 3064, 3064, 20150, 20257, 1568, 1588, 1458, 1498, 2139, 2149, 767, 699, 20022, 137, 20011, 30000, 30010, 30030, 30080, 30090, 30120, 30240, 30280, 30290, 30510, 30530, 34, 3077, 1697, 1835, 2178, 2306, 2443, 1558, 6142, 680, 31 |
| Lung volume | 80 | 21002, 30620, 30630, 30660, 30700, 30710, 30720, 30730, 30830, 30840, 30850, 30870, 30880, 23106, 23108, 23112, 23113, 23124, 23127, 48, 49, 50, 51, 20015, 21001, 102, 4079, 4079, 3143, 47, 3062, 3062, 3062, 3063, 3063, 3064, 3064, 3064, 20150, 20256, 20257, 20258, 1588, 1438, 1458, 904, 767, 699, 709, 20022, 20011, 30010, 30030, 30040, 30050, 30080, 30090, 30240, 30270, 30280, 30290, 34, 3077, 1647, 1697, 1835, 2345, 2306, 1558, 924, 1717, 1239, 1249, 20116, 20160, 6142, 21000, 680, 31 |
| Spleen volume | 69 | 21002, 30600, 30620, 30630, 30660, 30690, 30700, 30710, 30720, 30730, 30750, 30830, 30840, 30850, 30870, 30880, 23106, 23108, 23112, 23113, 23124, 23127, 48, 49, 50, 51, 20015, 21001, 3143, 47, 3062, 3062, 3062, 3063, 3063, 3064, 3064, 3064, 20150, 1588, 767, 30000, 30010, 30030, 30040, 30050, 30060, 30070, 30080, 30090, 30110, 30120, 30130, 30180, 30190, 30240, 30260, 30270, 30290, 30510, 30530, 34, 3077, 2306, 1717, 6142, 21000, 31 |
| Pancreas volume | 79 | 21002, 24005, 30600, 30610, 30620, 30630, 30650, 30670, 30700, 30720, 30730, 30740, 30750, 30770, 30830, 30850, 30880, 23106, 23108, 23112, 23113, 23124, 23127, 48, 49, 50, 51, 20015, 21001, 96, 4079, 4079, 3143, 47, 3062, 3062, 3062, 3063, 3063, 3064, 3064, 3064, 20150, 20256, 20257, 400, 404, 404, 404, 20023, 3526, 1588, 2149, 767, 699, 709, 20022, 137, 20011, 30010, 30030, 30510, 30530, 34, 3077, 1797, 1835, 2345, 2188, 2443, 2492, 6154, 1110, 6142, 680, 728, 738, 31 |
| Left Kidney volume | 74 | 21002, 30600, 30620, 30630, 30660, 30670, 30700, 30710, 30720, 30730, 30750, 30770, 30830, 30850, 30870, 30880, 23106, 23108, 23112, 23113, 23124, 23127, 48, 49, 50, 51, 20015, 21001, 4079, 4079, 3143, 47, 3062, 3062, 3062, 3063, 3063, 3064, 3064, 3064, 20150, 20256, 20257, 1588, 1498, 2139, 2149, 767, 699, 20022, 20011, 30010, 30030, 30240, 30290, 30510, 30530, 34, 3077, 1697, 1835, 2345, 2443, 6153, 1349, 1359, 1369, 1379, 20116, 6142, 680, 738, 31 |
| Right Kidney volume | 73 | 21002, 30600, 30620, 30630, 30650, 30660, 30670, 30700, 30710, 30720, 30730, 30740, 30750, 30830, 30850, 30870, 30880, 23106, 23108, 23112, 23113, 23124, 23127, 48, 49, 50, 51, 20015, 21001, 4079, 4079, 4080, 4080, 3143, 47, 3062, 3062, 3062, 3063, 3063, 3064, 3064, 3064, 20150, 20256, 20257, 1568, 1588, 1498, 2139, 2149, 767, 20022, 30010, 30030, 30240, 30290, 30510, 34, 3077, 1697, 2306, 2443, 6150, 6153, 1558, 1349, 1369, 1249, 20116, 6142, 31 |

**Supplementary table 3:** Imputation accuracy metrics and sample characteristics following IDP imputation. The table summarises R² (the squared Pearson correlation between predicted and MRI-derived IDPs in the independent test set (N = 5,258)), RMSE, and MAE values for each imaging-derived phenotype (IDP) based on out-of-sample performance in the test set (N = 5,258).

| **Organ / Tissue** | **Model** | **R-squared** | **RMSE** | **MAE** |
| --- | --- | --- | --- | --- |
| Abdominal subcutaneous fat | SVM | **0.9035** | **1204.47** | **919.36** |
|  | LR | 0.8963 | 1249.83 | 961.05 |
|  | RF | 0.8965 | 1266.41 | 977.67 |
|  | GB | 0.899 | 1233.59 | 947.02 |
| Visceral fat | SVM | **0.8358** | **932.94** | **701.32** |
|  | LR | 0.8187 | 983.16 | 750.26 |
|  | RF | 0.7986 | 1046.22 | 790.58 |
|  | GB | 0.8242 | 967.72 | 733.44 |
| Total Muscle volume | SVM | **0.9321** | **1175.15** | **893.03** |
|  | LR | 0.9267 | 1224.54 | 935.5 |
|  | RF | 0.9058 | 1394.18 | 1069.57 |
|  | GB | 0.9229 | 1250.13 | 955.02 |
| Left Iliopsoas Muscle volume | SVM | **0.8353** | **33.78** | **25.76** |
|  | LR | 0.8265 | 34.72 | 26.64 |
|  | RF | 0.8161 | 35.91 | 27.53 |
|  | GB | 0.8168 | 35.66 | 27.3 |
| Right Iliopsoas Muscle volume | SVM | **0.8361** | **34.03** | **26.19** |
|  | LR | 0.8295 | 34.72 | 26.7 |
|  | RF | 0.8161 | 36.31 | 27.95 |
|  | GB | 0.8217 | 35.54 | 27.36 |
| Liver volume | SVM | **0.6387** | **175.19** | **126.75** |
|  | LR | 0.6241 | 179.55 | 131.81 |
|  | RF | 0.6043 | 184.72 | 136.55 |
|  | GB | 0.6193 | 180.94 | 132.9 |
| Lung volume | SVM | **0.5521** | **488.48** | **343.33** |
|  | LR | 0.5066 | 510.01 | 361.07 |
|  | RF | 0.5241 | 505.24 | 366.25 |
|  | GB | 0.529 | 498.42 | 357.51 |
| Spleen volume | SVM | 0.4705 | **47.3** | **34.76** |
|  | LR | 0.4421 | 48.65 | 35.97 |
|  | RF | 0.4205 | 49.78 | 37.06 |
|  | GB | **0.4722** | 47.31 | 35.11 |
| Pancreas volume | SVM | **0.1893** | **14.3** | **11.31** |
|  | LR | 0.1906 | 14.31 | 11.35 |
|  | RF | 0.1757 | 14.46 | 11.45 |
|  | GB | 0.1419 | 14.82 | 11.74 |
| Left Kidney volume | SVM | 0.5012 | 21.14 | **15.53** |
|  | LR | 0.4821 | 21.55 | 15.86 |
|  | RF | 0.4511 | 22.39 | 16.72 |
|  | GB | **0.5025** | **21.1** | 15.65 |
| Right Kidney volume | SVM | 0.5436 | 19.33 | **14.31** |
|  | LR | 0.5281 | 19.69 | 14.55 |
|  | RF | 0.4943 | 20.51 | 15.46 |
|  | GB | **0.5445** | **19.32** | 14.39 |

**Supplementary Table 4.** Summary of imputed IDPs: imputation performance, effective sample sizes, and GWAS metrics.
This table provides an overview of the nine imaging-derived phenotypes (IDPs) analysed in this study. For each IDP, we report the imputation R² (from LDSC), effective sample size (calculated by POP-GWAS) represents the equivalent sample size of an unbiased GWAS with similar power, SNP heritability estimate (h²) with its standard error (h²_se), LD score regression intercept (Int) with standard error (Int_se), and the genomic inflation factor (λGC). r = Imputation quality from LDSC; N = Effective sample size from POP-GWAS; h² = SNP-based heritability; h²_se = Standard error of heritability estimate; Int = LDSC intercept; Int_se = Standard error of the intercept; λGC = Genomic control lambda.

| **Phenotype** | **r** | **Effective Sample Size (N)** | **h2** | **h2_se** | **Int** | **Int_se** | **λGC** |
| --- | --- | --- | --- | --- | --- | --- | --- |
| Abdominal subcutaneous fat | 0.857 | 112710 | 0.2259 | 0.0104 | 1.215 | 0.0104 | 1.5883 |
| Visceral fat | 0.791 | 87024 | 0.2149 | 0.0102 | 1.133 | 0.0092 | 1.4281 |
| Total muscle volume | 0.825 | 98463 | 0.29 | 0.015 | 1.167 | 0.0112 | 1.5401 |
| Iliopsoas muscle volume | 0.652 | 61192 | 0.3027 | 0.0212 | 1.074 | 0.0092 | 1.3203 |
| Liver volume | 0.72 | 71028 | 0.2231 | 0.014 | 1.104 | 0.0092 | 1.3306 |
| Lung volume | 0.647 | 60610 | 0.2207 | 0.0152 | 1.084 | 0.0093 | 1.2899 |
| Spleen volume | 0.631 | 58912 | 0.239 | 0.0234 | 1.131 | 0.0089 | 1.2899 |
| Pancreas volume | 0.207 | 37607 | 0.1777 | 0.0181 | 1.022 | 0.0078 | 1.1207 |
| Kidney volume | 0.683 | 65191 | 0.2605 | 0.0148 | 1.089 | 0.0102 | 1.3238 |

**Supplementary Table 5.** Summary of genomic loci, significant variants, instrument strength, and concordance between measured and imputed IDPs.
For each imaging-derived phenotype (IDP), this table reports: the number of genomic risk loci identified, the number of independent significant SNPs at LD threshold r² = 0.05, the number of clumped SNPs at r² = 0.01, F-statistics from the GWAS of imputed IDPs (POP-GWAS), and the correlation coefficient (Pearson’s r) between effect sizes from the measured and imputed GWAS. These metrics provide an overview of genetic signal strength, instrument validity, and consistency between measured and imputed trait analyses.

| **Phenotype** | **No. of Genomic risk loci** | **No. of Ind. Sig. SNPs** | **No. of Ind. Sig. SNPs after clumping** | **F-statistics** | **Correlation coefficient** |
| --- | --- | --- | --- | --- | --- |
| Abdominal subcutaneous fat | 65 | 76 | 67 | 44.18457 | 0.908 |
| Visceral fat | 32 | 36 | 33 | 42.90715 | 0.959 |
| Total muscle volume | 116 | 139 | 122 | 49.31725 | 0.896 |
| Iliopsoas muscle volume | 25 | 30 | 26 | 62.25260 | 0.928 |
| Liver volume | 34 | 44 | 35 | 64.63031 | 0.903 |
| Lung volume | 29 | 34 | 29 | 42.34447 | 0.918 |
| Spleen volume | 72 | 104 | 75 | 62.14944 | 0.938 |
| Pancreas volume | 28 | 29 | 28 | 44.44285 | 0.958 |
| Kidney volume | 47 | 56 | 52 | 45.59069 | 0.98 |
| Total | 448 | 548 | 467 (452 – unique) |  |  |

**Supplementary Table 6.** Independent genome-wide significant SNPs associated with abdominal subcutaneous fat volume. Reported columns include chromosome (Chr), SNP ID, base pair position, effect allele, other allele, effect size (beta), standard error (SE), and P-value. All results are based on POP-GWAS analysis in the non-imaging cohort.

| **SNP** | **chr** | **position** | **Effect allele** | **Other allele** | **beta** | **se** | **p-value** |
| --- | --- | --- | --- | --- | --- | --- | --- |
| rs61779328 | 1 | 39953467 | G | A | 0.03196 | 0.00541 | 3.58E-09 |
| rs164830 | 1 | 49758862 | A | G | -0.0246 | 0.00451 | 4.69E-08 |
| rs630372 | 1 | 177885762 | A | G | 0.03547 | 0.00498 | 1.02E-12 |
| rs6685593 | 1 | 203516075 | A | T | 0.02651 | 0.00421 | 3.13E-10 |
| rs2785990 | 1 | 219687432 | T | C | -0.0332 | 0.00443 | 6.98E-14 |
| rs62106258 | 2 | 417167 | C | T | -0.058 | 0.00971 | 2.36E-09 |
| rs6751993 | 2 | 635864 | G | A | 0.03593 | 0.00558 | 1.20E-10 |
| rs6725517 | 2 | 25129473 | G | A | 0.03003 | 0.00426 | 1.71E-12 |
| rs935166 | 2 | 26949366 | A | G | -0.029 | 0.00421 | 5.56E-12 |
| rs10460509 | 2 | 58772377 | A | T | -0.0275 | 0.00425 | 9.95E-11 |
| rs42825 | 2 | 59122090 | C | T | -0.0319 | 0.00525 | 1.19E-09 |
| rs6717858 | 2 | 165539661 | C | T | 0.02689 | 0.00429 | 3.68E-10 |
| rs6741676 | 2 | 181618654 | G | A | -0.0258 | 0.00444 | 6.87E-09 |
| rs10195674 | 2 | 229016094 | A | G | 0.02562 | 0.00445 | 8.51E-09 |
| rs2920503 | 3 | 12324230 | T | C | -0.0257 | 0.00464 | 3.15E-08 |
| rs6772763 | 3 | 15702641 | C | T | -0.0238 | 0.00425 | 2.27E-08 |
| rs2777888 | 3 | 49898000 | G | A | 0.02849 | 0.00421 | 1.36E-11 |
| rs73107812 | 3 | 70508856 | A | G | -0.0497 | 0.00837 | 2.78E-09 |
| rs13073568 | 3 | 94031839 | T | G | -0.0232 | 0.00422 | 3.58E-08 |
| rs10934646 | 3 | 123084541 | G | A | -0.0262 | 0.00437 | 2.14E-09 |
| rs58025624 | 3 | 141320585 | G | A | 0.04749 | 0.00843 | 1.81E-08 |
| rs34811474 | 4 | 25408838 | A | G | -0.0317 | 0.00496 | 1.68E-10 |
| rs10938398 | 4 | 45186139 | A | G | 0.02923 | 0.00425 | 5.97E-12 |
| rs11098901 | 4 | 80733423 | G | A | -0.0291 | 0.00518 | 1.90E-08 |
| rs2963822 | 5 | 59105755 | T | C | -0.0239 | 0.00422 | 1.42E-08 |
| rs10055091 | 5 | 75116421 | T | C | -0.057 | 0.01024 | 2.56E-08 |
| rs9393688 | 6 | 26207174 | T | A | -0.0314 | 0.00478 | 4.89E-11 |
| rs62398682 | 6 | 34433678 | T | G | -0.027 | 0.00459 | 3.89E-09 |
| rs2744973 | 6 | 34580221 | T | C | 0.04085 | 0.00459 | 5.51E-19 |
| rs19058433 | 6 | 35304354 | A | G | 0.10296 | 0.01564 | 4.58E-11 |
| rs72890671 | 6 | 50788566 | A | G | -0.0517 | 0.0089 | 6.27E-09 |
| rs35535237 | 6 | 50935295 | C | G | 0.03255 | 0.00468 | 3.61E-12 |
| rs4724822 | 7 | 6601568 | C | T | -0.0269 | 0.00481 | 2.26E-08 |
| rs6974400 | 7 | 130424075 | A | G | 0.02365 | 0.00421 | 1.97E-08 |
| rs11150531 | 7 | 133040106 | G | A | 0.07035 | 0.01153 | 1.04E-09 |
| rs12548555 | 8 | 73445459 | A | G | -0.0234 | 0.00426 | 4.16E-08 |
| rs1812736 | 8 | 76299138 | A | G | 0.03109 | 0.00559 | 2.66E-08 |
| rs6474551 | 9 | 1053060 | C | G | -0.0307 | 0.00556 | 3.27E-08 |
| rs12779865 | 10 | 21829162 | C | T | 0.03241 | 0.00448 | 4.91E-13 |
| rs16934748 | 10 | 33970319 | C | T | 0.03338 | 0.00589 | 1.44E-08 |
| rs11358547 | 10 | 33985434 | T | C | -0.0385 | 0.00705 | 4.67E-08 |
| rs11284265 | 11 | 27691085 | C | T | -0.0332 | 0.00446 | 8.77E-14 |
| rs9783304 | 11 | 43660255 | T | G | -0.0289 | 0.00457 | 2.69E-10 |
| rs3817334 | 11 | 47650993 | T | C | 0.02429 | 0.00429 | 1.45E-08 |
| rs11602616 | 11 | 72364077 | C | T | 0.0345 | 0.00575 | 1.92E-09 |
| rs12798944 | 11 | 120149741 | C | T | -0.0315 | 0.00576 | 4.48E-08 |
| rs7138803 | 12 | 50247468 | A | G | 0.03083 | 0.00437 | 1.73E-12 |
| rs74580294 | 12 | 122622795 | G | A | -0.0647 | 0.00968 | 2.23E-11 |
| rs12877270 | 13 | 97047020 | A | G | 0.02382 | 0.00424 | 2.01E-08 |
| rs14336812 | 14 | 33408139 | T | C | 0.03128 | 0.00555 | 1.75E-08 |
| rs72690737 | 14 | 79930644 | C | T | 0.02869 | 0.00508 | 1.65E-08 |
| rs34940743 | 14 | 80102233 | G | A | 0.0244 | 0.00443 | 3.67E-08 |
| rs1190226 | 14 | 103390383 | C | T | 0.03467 | 0.00538 | 1.13E-10 |
| rs12913596 | 15 | 47663870 | A | G | -0.0376 | 0.0069 | 4.94E-08 |
| rs4776970 | 15 | 68080886 | T | A | -0.026 | 0.00438 | 3.13E-09 |
| rs3826043 | 15 | 73618238 | T | C | -0.0235 | 0.00425 | 3.24E-08 |
| rs979543 | 15 | 84352564 | A | C | 0.03194 | 0.00513 | 4.76E-10 |
| rs879620 | 16 | 4015729 | T | C | 0.02596 | 0.00433 | 2.01E-09 |
| rs56186137 | 16 | 28825953 | G | A | 0.0374 | 0.0043 | 3.58E-18 |
| rs14082059 | 16 | 31021880 | T | G | 0.02718 | 0.00435 | 4.14E-10 |
| rs1421085 | 16 | 53800954 | C | T | 0.069 | 0.0043 | 6.16E-58 |
| rs62050612 | 16 | 69999032 | G | T | 0.03066 | 0.00476 | 1.14E-10 |
| rs11647479 | 16 | 70710549 | C | A | 0.02448 | 0.00421 | 6.27E-09 |
| rs12947658 | 17 | 65903006 | G | A | -0.0299 | 0.00461 | 8.64E-11 |
| rs58084604 | 18 | 57849429 | T | C | 0.03908 | 0.00498 | 4.03E-15 |
| rs76061089 | 18 | 58047158 | G | A | -0.1085 | 0.01482 | 2.52E-13 |
| rs59208569 | 19 | 4044424 | C | G | 0.0358 | 0.00565 | 2.42E-10 |
| rs56324036 | 19 | 18326931 | G | A | 0.04346 | 0.00466 | 1.16E-20 |
| rs4808846 | 19 | 18814330 | G | A | -0.026 | 0.00422 | 6.55E-10 |
| rs429358 | 19 | 45411941 | C | T | -0.06 | 0.00587 | 1.43E-24 |
| rs7412 | 19 | 45412079 | T | C | 0.04593 | 0.00775 | 3.04E-09 |
| rs10423928 | 19 | 46182304 | A | T | -0.033 | 0.00532 | 5.87E-10 |
| rs74687627 | 20 | 51100982 | T | C | -0.0468 | 0.00762 | 8.24E-10 |
| rs4819362 | 21 | 45420937 | G | A | -0.0297 | 0.00519 | 1.07E-08 |
| rs73152848 | 21 | 47934849 | C | A | 0.08939 | 0.01534 | 5.69E-09 |
| rs2231398 | 22 | 29656389 | T | G | 0.02495 | 0.00438 | 1.26E-08 |

**Supplementary table 7:** Independent genome-wide significant SNPs associated with Visceral fat volume. Reported columns include chromosome (Chr), SNP ID, base pair position, effect allele, other allele, effect size (beta), standard error (SE), and P-value. All results are based on POP-GWAS analysis in the non-imaging cohort.

| **SNP** | **chr** | **position** | **Effect allele** | **Other allele** | **beta** | **se** | **p-value** |
| --- | --- | --- | --- | --- | --- | --- | --- |
| rs61779328 | 1 | 39953467 | G | A | 0.04153 | 0.00616 | 1.59E-11 |
| rs56083993 | 2 | 26941482 | T | C | -0.0337 | 0.0048 | 2.24E-12 |
| rs982167 | 2 | 226176152 | A | T | 0.02803 | 0.00507 | 3.27E-08 |
| rs7561798 | 2 | 228973660 | G | A | 0.02747 | 0.0048 | 1.02E-08 |
| rs9860326 | 3 | 35683104 | G | C | 0.02806 | 0.0051 | 3.78E-08 |
| rs2352974 | 3 | 49890613 | T | C | 0.03289 | 0.0048 | 6.98E-12 |
| rs60654199 | 3 | 141267295 | A | C | 0.05464 | 0.0096 | 1.26E-08 |
| rs56082403 | 3 | 156797225 | C | T | 0.02988 | 0.00489 | 9.81E-10 |
| rs1073321 | 4 | 118512852 | C | T | 0.02874 | 0.0049 | 4.34E-09 |
| rs455660 | 5 | 55816888 | C | T | -0.0501 | 0.00613 | 2.99E-16 |
| rs67667580 | 5 | 103978833 | G | A | 0.02924 | 0.00483 | 1.41E-09 |
| rs2914231 | 5 | 158011435 | C | G | -0.0318 | 0.00566 | 1.95E-08 |
| rs359477 | 5 | 173305432 | C | T | 0.02719 | 0.00481 | 1.62E-08 |
| rs71531541 | 6 | 7066481 | T | C | 0.03159 | 0.00539 | 4.57E-09 |
| rs2797964 | 6 | 34185755 | T | C | -0.0475 | 0.00849 | 2.24E-08 |
| rs2744961 | 6 | 34655000 | T | C | 0.03181 | 0.005 | 2.02E-10 |
| rs11247127 | 7 | 98896068 | C | T | -0.0464 | 0.00797 | 5.70E-09 |
| rs11992444 | 8 | 25464690 | T | G | 0.04561 | 0.00479 | 1.85E-21 |
| rs12779865 | 10 | 21829162 | C | T | 0.0297 | 0.0051 | 5.87E-09 |
| rs11031796 | 11 | 32479807 | A | G | -0.0301 | 0.00494 | 1.09E-09 |
| rs1701704 | 12 | 56412487 | G | T | -0.0277 | 0.00504 | 3.90E-08 |
| rs66768878 | 16 | 4022638 | C | T | -0.0388 | 0.00654 | 3.08E-09 |
| rs80275162 | 16 | 28863517 | A | T | 0.02744 | 0.00491 | 2.30E-08 |
| rs62033400 | 16 | 53811788 | G | A | 0.04056 | 0.00491 | 1.48E-16 |
| rs904803 | 16 | 69970529 | G | A | 0.03411 | 0.00542 | 3.11E-10 |
| rs73534156 | 19 | 4058652 | C | T | -0.0337 | 0.00611 | 3.45E-08 |
| rs4808766 | 19 | 18335715 | C | G | 0.03978 | 0.00541 | 1.98E-13 |
| rs2081045 | 19 | 33789010 | A | G | -0.0416 | 0.00702 | 3.13E-09 |
| rs41355649 | 19 | 33790556 | A | G | -0.0786 | 0.00946 | 9.88E-17 |
| rs10406327 | 19 | 33890838 | G | C | 0.04182 | 0.0048 | 2.92E-18 |
| rs429358 | 19 | 45411941 | C | T | -0.0669 | 0.00668 | 1.25E-23 |
| rs7247937 | 19 | 45827863 | C | G | 0.0306 | 0.00532 | 9.01E-09 |
| rs35277443 | 20 | 51233000 | A | C | -0.0358 | 0.00581 | 7.31E-10 |
| rs6024475 | 20 | 54414271 | G | A | 0.028 | 0.00508 | 3.64E-08 |
| rs28451064 | 21 | 35593827 | A | G | 0.04202 | 0.00703 | 2.22E-09 |
| rs2255955 | 21 | 45493560 | A | T | -0.0361 | 0.00612 | 3.65E-09 |

**Supplementary table 8:** Independent genome-wide significant SNPs associated with total muscle volume. Reported columns include chromosome (Chr), SNP ID, base pair position, effect allele, other allele, effect size (beta), standard error (SE), and P-value. All results are based on POP-GWAS analysis in the non-imaging cohort.

| **SNP** | **chr** | **position** | **Effect allele** | **Other allele** | **beta** | **se** | **p-value** |
| --- | --- | --- | --- | --- | --- | --- | --- |
| rs6693113 | 1 | 36757801 | T | G | 0.02514 | 0.00451 | 2.44E-08 |
| rs61780439 | 1 | 41490177 | A | G | 0.03906 | 0.00545 | 7.83E-13 |
| rs12040689 | 1 | 92644370 | A | G | 0.03156 | 0.00571 | 3.25E-08 |
| rs10858097 | 1 | 109967324 | A | C | -0.0301 | 0.00499 | 1.61E-09 |
| rs7519368 | 1 | 113150644 | A | T | 0.02791 | 0.00508 | 4.02E-08 |
| rs585944 | 1 | 177882786 | T | C | 0.04176 | 0.00533 | 4.48E-15 |
| rs28823884 | 1 | 184103187 | T | A | 0.03215 | 0.00459 | 2.41E-12 |
| rs3850625 | 1 | 201016296 | A | G | -0.0588 | 0.00699 | 3.99E-17 |
| rs62106252 | 2 | 408713 | A | G | -0.0652 | 0.01033 | 2.85E-10 |
| rs1320332 | 2 | 621954 | C | T | 0.05858 | 0.00597 | 9.82E-23 |
| rs17045981 | 2 | 24109316 | G | A | -0.039 | 0.00598 | 7.19E-11 |
| rs1260326 | 2 | 27730940 | C | T | 0.03614 | 0.00462 | 5.00E-15 |
| rs17758988 | 2 | 28311121 | G | A | -0.035 | 0.00492 | 1.14E-12 |
| rs17759740 | 2 | 28393115 | A | T | 0.0424 | 0.00703 | 1.59E-09 |
| rs1523785 | 2 | 36737784 | A | G | 0.02693 | 0.00484 | 2.57E-08 |
| rs12713126 | 2 | 51187589 | C | G | -0.038 | 0.00685 | 3.08E-08 |
| rs2271198 | 2 | 65559027 | G | A | 0.02495 | 0.00456 | 4.36E-08 |
| rs1047891 | 2 | 211540507 | A | C | 0.03433 | 0.00485 | 1.48E-12 |
| rs2541381 | 2 | 217683836 | T | G | 0.03364 | 0.00451 | 8.78E-14 |
| rs4675045 | 2 | 226952104 | G | C | 0.03034 | 0.00549 | 3.27E-08 |
| rs1515098 | 2 | 227073854 | T | C | 0.03271 | 0.00484 | 1.41E-11 |
| rs307569 | 3 | 12124003 | C | A | -0.0375 | 0.00658 | 1.24E-08 |
| rs7649984 | 3 | 38570221 | T | C | -0.0398 | 0.00464 | 8.48E-18 |
| rs2200464 | 3 | 85157148 | C | T | 0.03147 | 0.00469 | 1.98E-11 |
| rs6440006 | 3 | 141142691 | A | G | 0.05059 | 0.00453 | 6.38E-29 |
| rs582780 | 3 | 172121443 | G | A | 0.03039 | 0.00456 | 2.60E-11 |
| rs73175572 | 3 | 185490184 | G | A | 0.03927 | 0.00716 | 4.08E-08 |
| rs13070996 | 3 | 196906074 | T | G | 0.03333 | 0.00502 | 3.11E-11 |
| rs6854757 | 4 | 17932555 | G | A | -0.053 | 0.00598 | 7.94E-19 |
| rs2061456 | 4 | 17998426 | A | C | 0.05335 | 0.00512 | 2.14E-25 |
| rs1435211 | 4 | 54379445 | A | G | 0.03886 | 0.00625 | 5.18E-10 |
| rs57222629 | 4 | 82195346 | C | G | 0.03397 | 0.00487 | 3.03E-12 |
| rs2647239 | 4 | 106217349 | G | A | -0.0351 | 0.00456 | 1.40E-14 |
| rs62346126 | 4 | 145560166 | A | C | 0.04275 | 0.00577 | 1.33E-13 |
| rs62372052 | 5 | 42724294 | G | A | 0.04159 | 0.00715 | 5.96E-09 |
| rs702634 | 5 | 53271420 | A | G | -0.0278 | 0.00487 | 1.19E-08 |
| rs1466666 | 5 | 67632194 | G | C | 0.02562 | 0.00458 | 2.21E-08 |
| rs2307111 | 5 | 75003678 | C | T | -0.0325 | 0.00461 | 1.73E-12 |
| rs1582931 | 5 | 122657199 | A | G | -0.028 | 0.00451 | 5.84E-10 |
| rs31211 | 5 | 134363145 | A | G | -0.0338 | 0.0052 | 8.37E-11 |
| rs4282339 | 5 | 168256240 | A | G | -0.0317 | 0.0056 | 1.46E-08 |
| rs55795753 | 5 | 171442208 | T | G | 0.02495 | 0.00452 | 3.39E-08 |
| rs4960348 | 6 | 7723175 | T | C | 0.03237 | 0.0048 | 1.58E-11 |
| rs10949583 | 6 | 19129293 | T | C | 0.03226 | 0.00547 | 3.65E-09 |
| rs41271299 | 6 | 19839415 | T | C | 0.06413 | 0.01014 | 2.57E-10 |
| rs7766641 | 6 | 26184102 | A | G | -0.0374 | 0.00513 | 3.20E-13 |
| rs1187115 | 6 | 34172055 | A | C | -0.0617 | 0.006 | 8.54E-25 |
| rs2814973 | 6 | 34569720 | T | C | 0.0473 | 0.00651 | 3.73E-13 |
| rs13213103 | 6 | 35069112 | C | T | -0.0412 | 0.00655 | 3.28E-10 |
| rs55694295 | 6 | 35675579 | T | C | 0.03172 | 0.00557 | 1.25E-08 |
| rs33966734 | 6 | 41903798 | A | C | -0.1313 | 0.01957 | 1.95E-11 |
| rs9343977 | 6 | 80967001 | T | C | -0.0382 | 0.00451 | 2.41E-17 |
| rs14959652 | 6 | 81533301 | G | T | -0.057 | 0.01026 | 2.81E-08 |
| rs3813498 | 6 | 108944165 | T | C | 0.03751 | 0.00576 | 7.68E-11 |
| rs645426 | 6 | 117109229 | C | T | 0.02563 | 0.00467 | 3.93E-08 |
| rs1490384 | 6 | 126851160 | T | C | 0.05243 | 0.00451 | 2.80E-31 |
| rs7740107 | 6 | 130374461 | A | T | -0.0347 | 0.00509 | 9.30E-12 |
| rs488133 | 6 | 152125444 | C | T | 0.03024 | 0.00477 | 2.25E-10 |
| rs2457982 | 6 | 166313684 | A | G | 0.02924 | 0.00504 | 6.82E-09 |
| rs2533879 | 7 | 2859847 | A | G | -0.0339 | 0.00489 | 4.23E-12 |
| rs34776209 | 7 | 23513093 | T | C | -0.0359 | 0.00522 | 6.13E-12 |
| rs74445550 | 7 | 46391213 | T | A | -0.0456 | 0.00797 | 1.02E-08 |
| rs71556736 | 7 | 73034929 | T | C | 0.04183 | 0.00672 | 4.95E-10 |
| rs1001220 | 7 | 73110603 | T | C | 0.02886 | 0.0046 | 3.51E-10 |
| rs42235 | 7 | 92248076 | T | C | 0.03455 | 0.00493 | 2.48E-12 |
| rs10236214 | 7 | 150668070 | T | C | 0.03536 | 0.0047 | 5.26E-14 |
| rs4871880 | 8 | 23393572 | A | G | -0.0374 | 0.0062 | 1.57E-09 |
| rs72656010 | 8 | 57122215 | C | T | -0.0525 | 0.00671 | 5.20E-15 |
| rs62515432 | 8 | 57123523 | C | T | 0.03414 | 0.00553 | 6.53E-10 |
| rs12680214 | 8 | 78146537 | T | C | 0.03118 | 0.00459 | 1.09E-11 |
| rs2142331 | 8 | 116636719 | T | C | -0.0285 | 0.0046 | 5.58E-10 |
| rs13271368 | 8 | 126506140 | T | C | -0.0316 | 0.0053 | 2.39E-09 |
| rs72718335 | 8 | 130789472 | T | C | -0.0397 | 0.00669 | 2.93E-09 |
| rs894344 | 8 | 135612745 | G | A | -0.0287 | 0.00459 | 3.80E-10 |
| rs9409514 | 9 | 97065439 | T | C | -0.0311 | 0.00467 | 2.97E-11 |
| rs473902 | 9 | 98256235 | G | T | -0.067 | 0.00789 | 2.18E-17 |
| rs16909922 | 9 | 98265901 | G | A | 0.06955 | 0.00772 | 2.09E-19 |
| rs10761022 | 9 | 99224746 | T | A | 0.03486 | 0.00545 | 1.58E-10 |
| rs12347137 | 9 | 119122721 | C | A | -0.0365 | 0.00556 | 5.36E-11 |
| rs35843221 | 9 | 128013848 | G | T | -0.029 | 0.00453 | 1.61E-10 |
| rs57467326 | 10 | 102317557 | G | T | -0.0341 | 0.00554 | 6.96E-10 |
| rs35525740 | 10 | 104941112 | T | A | 0.0324 | 0.00465 | 3.36E-12 |
| rs11598539 | 10 | 130842093 | C | T | 0.02931 | 0.00519 | 1.60E-08 |
| rs2049045 | 11 | 27694241 | C | G | -0.038 | 0.00577 | 4.60E-11 |
| rs7952436 | 11 | 67024534 | T | C | -0.0658 | 0.00817 | 7.52E-16 |
| rs2510404 | 11 | 68425034 | T | A | 0.03856 | 0.00576 | 2.24E-11 |
| rs11822234 | 11 | 74385146 | T | C | -0.0292 | 0.00459 | 2.10E-10 |
| rs73047065 | 12 | 3364536 | T | G | 0.06367 | 0.00928 | 6.67E-12 |
| rs3217836 | 12 | 4394017 | T | C | -0.0356 | 0.00629 | 1.57E-08 |
| rs17818624 | 12 | 11896846 | T | C | 0.027 | 0.00467 | 7.26E-09 |
| rs73123652 | 12 | 65874956 | C | T | -0.0512 | 0.00712 | 6.25E-13 |
| rs11746034 | 12 | 66329978 | A | C | -0.0838 | 0.01514 | 3.08E-08 |
| rs7306710 | 12 | 66376091 | C | T | -0.0458 | 0.00451 | 3.20E-24 |
| rs4301837 | 12 | 102336310 | C | T | -0.033 | 0.00451 | 2.54E-13 |
| rs57078519 | 12 | 124388784 | A | G | -0.0302 | 0.00476 | 2.19E-10 |
| rs1998690 | 13 | 22346432 | G | T | -0.0275 | 0.00478 | 8.81E-09 |
| rs9594689 | 13 | 42737268 | G | T | -0.0259 | 0.00466 | 2.77E-08 |
| rs2812208 | 13 | 50707087 | C | G | 0.15424 | 0.01512 | 1.96E-24 |
| rs3116615 | 13 | 51158752 | C | T | -0.0505 | 0.00552 | 5.83E-20 |
| rs9530207 | 13 | 74162373 | A | G | 0.03377 | 0.00453 | 9.50E-14 |
| rs12882669 | 14 | 35178297 | G | A | 0.0257 | 0.00453 | 1.37E-08 |
| rs11763509 | 14 | 42518957 | A | G | -0.1196 | 0.0214 | 2.30E-08 |
| rs4899012 | 14 | 61003889 | C | G | -0.0374 | 0.00461 | 4.94E-16 |
| rs28559926 | 15 | 89400043 | C | G | -0.0677 | 0.01169 | 6.82E-09 |
| rs2871865 | 15 | 99194896 | G | C | -0.057 | 0.00707 | 8.17E-16 |
| rs72755233 | 15 | 100692953 | A | G | -0.0397 | 0.00708 | 2.05E-08 |
| rs12443906 | 16 | 4948936 | T | C | -0.0307 | 0.00504 | 1.09E-09 |
| rs246174 | 16 | 14379931 | T | C | 0.02672 | 0.00466 | 1.02E-08 |
| rs66674044 | 16 | 19904344 | T | A | -0.0371 | 0.00639 | 6.49E-09 |
| rs1140239 | 16 | 30021402 | T | C | 0.02972 | 0.00459 | 9.68E-11 |
| rs17817964 | 16 | 53828066 | T | C | 0.03608 | 0.00462 | 5.79E-15 |
| rs4783618 | 16 | 68356419 | G | A | -0.0267 | 0.00461 | 6.75E-09 |
| rs9940093 | 16 | 89558705 | A | G | -0.0268 | 0.00453 | 3.15E-09 |
| rs11658072 | 17 | 7214824 | T | C | 0.02768 | 0.00468 | 3.38E-09 |
| rs78378222 | 17 | 7571752 | G | T | 0.1206 | 0.02043 | 3.54E-09 |
| rs79461387 | 17 | 29168077 | T | G | -0.0395 | 0.00512 | 1.18E-14 |
| rs9894577 | 17 | 43223292 | A | G | -0.0311 | 0.00486 | 1.46E-10 |
| rs227723 | 17 | 54778904 | T | C | 0.03381 | 0.00488 | 4.50E-12 |
| rs2005172 | 17 | 61996255 | C | A | 0.04205 | 0.0047 | 3.54E-19 |
| rs79148459 | 17 | 62053388 | C | T | 0.08097 | 0.01456 | 2.69E-08 |
| rs12452505 | 17 | 63556402 | G | C | -0.0402 | 0.00649 | 6.16E-10 |
| rs66472386 | 17 | 74237294 | T | C | 0.04886 | 0.0083 | 3.99E-09 |
| rs4800451 | 18 | 20716805 | T | C | 0.03009 | 0.00515 | 5.29E-09 |
| rs538656 | 18 | 57850422 | T | G | 0.05602 | 0.00532 | 6.05E-26 |
| rs4807473 | 19 | 3448869 | G | A | -0.0266 | 0.00469 | 1.51E-08 |
| rs4804376 | 19 | 7194766 | T | C | 0.0498 | 0.0081 | 7.88E-10 |
| rs55930881 | 19 | 30343667 | C | G | 0.03394 | 0.00513 | 3.53E-11 |
| rs10119 | 19 | 45406673 | A | G | -0.0305 | 0.00498 | 9.39E-10 |
| rs45474992 | 19 | 47724564 | T | C | -0.0702 | 0.01208 | 6.11E-09 |
| rs14711093 | 19 | 55993436 | T | G | -0.0804 | 0.01443 | 2.48E-08 |
| rs2424352 | 20 | 21215001 | A | C | -0.032 | 0.00478 | 2.30E-11 |
| rs293738 | 20 | 31925918 | G | C | 0.04676 | 0.00601 | 7.29E-15 |
| rs8114616 | 20 | 32969092 | T | G | -0.0496 | 0.00451 | 3.86E-28 |
| rs2247828 | 20 | 33839939 | G | A | 0.03827 | 0.00654 | 5.00E-09 |
| rs143384 | 20 | 34025756 | G | A | 0.06521 | 0.00459 | 8.08E-46 |
| rs1204552 | 20 | 34638903 | A | T | -0.0628 | 0.00832 | 4.64E-14 |
| rs2057028 | 20 | 42750661 | C | A | -0.0407 | 0.0074 | 3.73E-08 |
| rs9306468 | 22 | 30374281 | C | T | -0.0404 | 0.00462 | 2.59E-18 |
| rs20207315 | 22 | 46439738 | C | A | 0.05349 | 0.00915 | 5.08E-09 |

**Supplementary table 9:** Independent genome-wide significant SNPs associated with Iliopsoas muscle volume. Reported columns include chromosome (Chr), SNP ID, base pair position, effect allele, other allele, effect size (beta), standard error (SE), and P-value. All results are based on POP-GWAS analysis in the non-imaging cohort.

| **SNP** | **chr** | **position** | **Effect allele** | **Other allele** | **beta** | **se** | **p-value** |
| --- | --- | --- | --- | --- | --- | --- | --- |
| rs3850625 | 1 | 201016296 | A | G | -0.0657 | 0.00887 | 1.29E-13 |
| rs6547692 | 2 | 27734972 | A | G | 0.03896 | 0.00576 | 1.29E-11 |
| rs2712184 | 2 | 217682779 | A | C | 0.04396 | 0.00578 | 2.84E-14 |
| rs3103277 | 2 | 233095677 | A | T | -0.0417 | 0.00644 | 9.23E-11 |
| rs2370840 | 3 | 38529825 | T | C | 0.04169 | 0.00576 | 4.43E-13 |
| rs9853018 | 3 | 141101961 | T | C | 0.04223 | 0.00575 | 2.09E-13 |
| rs6854757 | 4 | 17932555 | G | A | -0.0504 | 0.00759 | 3.22E-11 |
| rs2610989 | 4 | 18022834 | C | T | -0.0583 | 0.00649 | 2.97E-19 |
| rs62302275 | 4 | 82139174 | G | A | 0.04437 | 0.00637 | 3.38E-12 |
| rs73116951 | 5 | 68043900 | T | C | -0.0663 | 0.0101 | 5.32E-11 |
| rs479632 | 5 | 134364518 | G | C | -0.049 | 0.0066 | 1.22E-13 |
| rs1776898 | 6 | 34194092 | T | C | -0.0715 | 0.01011 | 1.57E-12 |
| rs6925689 | 6 | 126865884 | C | T | 0.04146 | 0.00572 | 4.10E-13 |
| rs9389994 | 6 | 142789158 | T | C | -0.0511 | 0.0063 | 4.66E-16 |
| rs12532246 | 7 | 156276306 | G | A | 0.0528 | 0.00637 | 1.19E-16 |
| rs142327126 | 8 | 78115715 | T | A | 0.04532 | 0.00635 | 9.52E-13 |
| rs7860822 | 9 | 126341464 | G | A | 0.04586 | 0.00593 | 1.08E-14 |
| rs79997404 | 12 | 3390803 | A | G | 0.09155 | 0.01197 | 2.02E-14 |
| rs2812208 | 13 | 50707087 | C | G | 0.15371 | 0.01918 | 1.11E-15 |
| rs3116617 | 13 | 51167209 | A | T | -0.0564 | 0.00701 | 8.33E-16 |
| rs7319941 | 13 | 74188590 | A | G | 0.05226 | 0.00572 | 6.25E-20 |
| rs2093210 | 14 | 60957279 | T | C | -0.0516 | 0.00585 | 1.28E-18 |
| rs28410907 | 15 | 99206835 | A | G | -0.0493 | 0.009 | 4.29E-08 |
| rs1965308 | 16 | 68355181 | T | A | -0.0468 | 0.00583 | 9.90E-16 |
| rs72832859 | 17 | 43188891 | G | C | 0.0413 | 0.00722 | 1.07E-08 |
| rs8114616 | 20 | 32969092 | T | G | -0.0535 | 0.00572 | 8.12E-21 |
| rs143384 | 20 | 34025756 | G | A | 0.08668 | 0.00582 | 3.81E-50 |
| rs41290920 | 20 | 34088151 | C | T | -0.1117 | 0.0181 | 6.67E-10 |
| rs4911523 | 20 | 34545495 | G | A | 0.07464 | 0.0101 | 1.45E-13 |
| rs140147 | 22 | 30184599 | A | G | -0.0473 | 0.00586 | 7.15E-16 |

**Supplementary table 10:** Independent genome-wide significant SNPs associated with liver volume. Reported columns include chromosome (Chr), SNP ID, base pair position, effect allele, other allele, effect size (beta), standard error (SE), and P-value. All results are based on POP-GWAS analysis in the non-imaging cohort.

| SNP | chr | position | Effect allele | Other allele | beta | se | p-vakye |
| --- | --- | --- | --- | --- | --- | --- | --- |
| rs114165349 | 1 | 27021913 | C | G | 0.12048 | 0.01742 | 4.68E-12 |
| rs585944 | 1 | 177882786 | T | C | 0.04327 | 0.00627 | 5.15E-12 |
| rs62107261 | 2 | 422144 | C | T | -0.0847 | 0.01223 | 4.47E-12 |
| rs6751993 | 2 | 635864 | G | A | 0.04278 | 0.00703 | 1.16E-09 |
| rs61737373 | 2 | 27423913 | A | G | -0.0734 | 0.01137 | 1.11E-10 |
| rs1260326 | 2 | 27730940 | C | T | -0.0747 | 0.00544 | 5.35E-43 |
| rs17050272 | 2 | 121306440 | A | G | -0.0325 | 0.0054 | 1.71E-09 |
| rs9843361 | 3 | 94091416 | G | A | 0.03215 | 0.00535 | 1.84E-09 |
| rs2871960 | 3 | 141121814 | C | A | 0.03935 | 0.00534 | 1.65E-13 |
| rs4894553 | 3 | 172199795 | C | T | -0.0368 | 0.00587 | 3.83E-10 |
| rs79287178 | 3 | 172294500 | A | G | 0.14292 | 0.01525 | 7.25E-21 |
| rs1154416 | 4 | 99996373 | T | G | 0.03521 | 0.0054 | 6.84E-11 |
| rs7661349 | 4 | 106066982 | C | T | 0.03281 | 0.0055 | 2.50E-09 |
| rs4449583 | 5 | 1284135 | T | C | 0.03278 | 0.00566 | 7.11E-09 |
| rs6864091 | 5 | 75010002 | C | T | -0.0318 | 0.00553 | 9.25E-09 |
| rs806794 | 6 | 26200677 | G | A | -0.0341 | 0.00595 | 9.56E-09 |
| rs144100226 | 6 | 34180297 | T | C | 0.08461 | 0.01266 | 2.34E-11 |
| rs1490384 | 6 | 126851160 | T | C | 0.04443 | 0.00531 | 5.62E-17 |
| rs42041 | 7 | 92246744 | G | C | 0.03576 | 0.00616 | 6.48E-09 |
| rs441177 | 8 | 9170929 | T | G | 0.04042 | 0.00689 | 4.50E-09 |
| rs4240624 | 8 | 9184231 | A | G | -0.1691 | 0.00926 | 1.60E-74 |
| rs732839 | 8 | 9194172 | A | G | -0.053 | 0.0063 | 4.34E-17 |
| rs59197158 | 8 | 9350576 | T | G | -0.0331 | 0.00567 | 5.11E-09 |
| rs2721933 | 8 | 116627175 | C | T | -0.0317 | 0.00534 | 2.87E-09 |
| rs2980888 | 8 | 126507308 | C | T | -0.0438 | 0.00578 | 3.66E-14 |
| rs10761753 | 10 | 65161903 | A | G | -0.0637 | 0.00531 | 3.80E-33 |
| rs11814936 | 10 | 65323857 | A | G | 0.07843 | 0.01288 | 1.15E-09 |
| rs1044563 | 10 | 93390149 | T | C | 0.06254 | 0.01098 | 1.22E-08 |
| rs10881959 | 10 | 93507964 | T | G | 0.04782 | 0.00536 | 4.45E-19 |
| rs3750925 | 11 | 49598207 | C | T | -0.0306 | 0.00532 | 9.28E-09 |
| rs4149083 | 12 | 21380630 | T | A | 0.04027 | 0.00728 | 3.16E-08 |
| rs7484541 | 12 | 57714803 | T | A | -0.0378 | 0.00631 | 2.02E-09 |
| rs184334219 | 15 | 42721845 | A | G | 0.12548 | 0.0193 | 7.96E-11 |
| rs139974673 | 15 | 44027885 | C | T | 0.21615 | 0.01709 | 1.11E-36 |
| rs35467921 | 16 | 30048553 | T | C | 0.03195 | 0.00541 | 3.39E-09 |
| rs56094641 | 16 | 53806453 | G | A | 0.04403 | 0.00541 | 4.24E-16 |
| rs12933677 | 16 | 83983775 | C | T | -0.0337 | 0.00532 | 2.44E-10 |
| rs56305452 | 17 | 65990500 | A | C | -0.0367 | 0.00649 | 1.60E-08 |
| rs35693910 | 18 | 57913965 | A | G | 0.04786 | 0.00598 | 1.16E-15 |
| rs187429064 | 19 | 19380513 | G | A | 0.12621 | 0.0221 | 1.12E-08 |
| rs8107974 | 19 | 19388500 | T | A | 0.08094 | 0.01001 | 6.16E-16 |
| rs6857 | 19 | 45392254 | T | C | -0.0632 | 0.00708 | 4.74E-19 |
| rs113469203 | 20 | 25343258 | A | G | -0.0314 | 0.00535 | 4.44E-09 |
| rs3747207 | 22 | 44324855 | A | G | 0.04941 | 0.00648 | 2.52E-14 |

**Supplementary table 11:** Independent genome-wide significant SNPs associated with lung volume. Reported columns include chromosome (Chr), SNP ID, base pair position, effect allele, other allele, effect size (beta), standard error (SE), and P-value. All results are based on POP-GWAS analysis in the non-imaging cohort.

| **SNP** | **chr** | **position** | **Effect allele** | **Other allele** | **beta** | **se** | **p-value** |
| --- | --- | --- | --- | --- | --- | --- | --- |
| rs3754512 | 1 | 17309718 | T | C | 0.05068 | 0.00575 | 1.22E-18 |
| rs4970708 | 1 | 92986514 | T | G | 0.04554 | 0.00708 | 1.27E-10 |
| rs2384090 | 2 | 25406190 | C | T | 0.04983 | 0.00745 | 2.23E-11 |
| rs3791675 | 2 | 56111309 | T | C | -0.041 | 0.0068 | 1.61E-09 |
| rs16825267 | 2 | 229569919 | G | C | -0.0647 | 0.01048 | 6.75E-10 |
| rs11900720 | 2 | 232790491 | T | C | -0.0502 | 0.00885 | 1.46E-08 |
| rs7628338 | 3 | 72432888 | A | G | 0.03407 | 0.00597 | 1.15E-08 |
| rs9681202 | 3 | 158044880 | G | A | -0.035 | 0.00601 | 5.92E-09 |
| rs2194411 | 3 | 185548663 | A | G | 0.0605 | 0.00861 | 2.14E-12 |
| rs4144829 | 4 | 17903654 | T | C | -0.0466 | 0.00651 | 8.14E-13 |
| rs6854757 | 4 | 17932555 | G | A | -0.0438 | 0.00762 | 9.11E-09 |
| rs79634326 | 4 | 145461119 | C | T | 0.05715 | 0.00979 | 5.37E-09 |
| rs1844428 | 4 | 145574196 | G | A | -0.0474 | 0.00843 | 1.90E-08 |
| rs1497126 | 4 | 146463966 | C | T | -0.0331 | 0.00582 | 1.39E-08 |
| rs2184968 | 6 | 126760994 | C | T | 0.0366 | 0.00577 | 2.28E-10 |
| rs55987383 | 6 | 142338903 | G | A | -0.0365 | 0.00649 | 1.82E-08 |
| rs1891308 | 6 | 142648235 | A | G | -0.0629 | 0.00635 | 4.15E-23 |
| rs1872584 | 7 | 46324724 | T | G | -0.0598 | 0.0106 | 1.76E-08 |
| rs357550 | 9 | 98138533 | T | C | 0.03561 | 0.00614 | 6.53E-09 |
| rs60150206 | 9 | 98262223 | A | G | 0.09895 | 0.01007 | 9.02E-23 |
| rs2762599 | 10 | 7250998 | C | G | -0.1039 | 0.01821 | 1.15E-08 |
| rs10828247 | 10 | 21822856 | G | A | -0.0395 | 0.00606 | 7.42E-11 |
| rs6586119 | 10 | 89863723 | G | T | -0.0359 | 0.00624 | 9.01E-09 |
| rs7306710 | 12 | 66376091 | C | T | -0.032 | 0.00575 | 2.53E-08 |
| rs940904 | 12 | 123491572 | A | G | -0.0382 | 0.00652 | 4.83E-09 |
| rs3118906 | 13 | 51106788 | A | G | -0.0396 | 0.00641 | 6.77E-10 |
| rs5742915 | 15 | 74336633 | C | T | 0.03171 | 0.00576 | 3.64E-08 |
| rs10520595 | 15 | 86128717 | T | C | -0.0359 | 0.00615 | 4.95E-09 |
| rs16942341 | 15 | 89388905 | T | C | -0.0999 | 0.01717 | 5.89E-09 |
| rs11075732 | 16 | 69766038 | G | A | 0.03217 | 0.00582 | 3.22E-08 |
| rs227725 | 17 | 54778620 | C | T | 0.03554 | 0.00606 | 4.56E-09 |
| rs149394327 | 17 | 64228995 | C | G | -0.103 | 0.0172 | 2.10E-09 |
| rs117700741 | 19 | 8637832 | T | C | -0.0889 | 0.01487 | 2.23E-09 |
| rs143384 | 20 | 34025756 | G | A | 0.03925 | 0.00585 | 1.96E-11 |

**Supplementary table 12:** Independent genome-wide significant SNPs associated with spleen volume. Reported columns include chromosome (Chr), SNP ID, base pair position, effect allele, other allele, effect size (beta), standard error (SE), and P-value. All results are based on POP-GWAS analysis in the non-imaging cohort.

| **SNP** | **chr** | **position** | **Effect allele** | **Other allele** | **beta** | **se** | **p-value** |
| --- | --- | --- | --- | --- | --- | --- | --- |
| rs79898419 | 1 | 91587561 | G | A | -0.04 | 0.00678 | 3.66E-09 |
| rs2482963 | 1 | 158596438 | T | G | -0.0589 | 0.00659 | 3.77E-19 |
| rs4607954 | 1 | 204266022 | G | A | 0.05602 | 0.00656 | 1.36E-17 |
| rs4653647 | 1 | 225860310 | T | C | -0.0503 | 0.00834 | 1.61E-09 |
| rs10915869 | 1 | 225937402 | A | G | 0.07081 | 0.00952 | 9.97E-14 |
| rs56043070 | 1 | 247719769 | A | G | 0.09279 | 0.0113 | 2.17E-16 |
| rs6697193 | 1 | 249182887 | C | T | 0.05041 | 0.00786 | 1.42E-10 |
| rs35330522 | 2 | 12877060 | A | G | -0.0335 | 0.00583 | 8.98E-09 |
| rs7569648 | 2 | 159957389 | A | T | 0.03472 | 0.00634 | 4.27E-08 |
| rs10200279 | 2 | 202170655 | C | T | -0.0465 | 0.00652 | 9.90E-13 |
| rs11676298 | 2 | 227291731 | G | C | -0.0695 | 0.0074 | 5.68E-21 |
| rs144154898 | 2 | 227302076 | A | G | -0.164 | 0.02745 | 2.31E-09 |
| rs160113 | 3 | 12145239 | C | T | 0.04052 | 0.00701 | 7.55E-09 |
| rs12491937 | 3 | 12268244 | G | A | 0.07146 | 0.00591 | 1.08E-33 |
| rs62248908 | 3 | 47297060 | G | A | 0.05554 | 0.00761 | 2.96E-13 |
| rs76311404 | 3 | 123099129 | A | G | 0.05242 | 0.00707 | 1.19E-13 |
| rs2713602 | 3 | 128203035 | G | A | -0.0338 | 0.00591 | 1.03E-08 |
| rs6441319 | 3 | 160139696 | A | G | 0.03677 | 0.00585 | 3.25E-10 |
| rs73063010 | 3 | 185538006 | G | A | 0.04558 | 0.00674 | 1.40E-11 |
| rs11715549 | 3 | 188088118 | G | C | 0.03343 | 0.00585 | 1.12E-08 |
| rs2089979 | 3 | 196501413 | G | A | 0.04762 | 0.00591 | 7.67E-16 |
| rs2881559 | 4 | 110840946 | G | A | 0.06168 | 0.00593 | 2.48E-25 |
| rs7692994 | 4 | 120427489 | A | C | 0.03899 | 0.00618 | 2.79E-10 |
| rs7725218 | 5 | 1282414 | A | G | 0.04523 | 0.00617 | 2.30E-13 |
| rs10474548 | 5 | 77745537 | G | A | -0.0331 | 0.00583 | 1.41E-08 |
| rs34530707 | 5 | 139059016 | A | G | 0.03732 | 0.00621 | 1.83E-09 |
| rs2328765 | 6 | 23559114 | G | C | 0.03728 | 0.00583 | 1.58E-10 |
| rs9257182 | 6 | 28754817 | A | C | 0.04263 | 0.00721 | 3.40E-09 |
| rs10807138 | 6 | 34184938 | A | G | -0.0545 | 0.00723 | 4.63E-14 |
| rs33966734 | 6 | 41903798 | A | C | -0.1384 | 0.0253 | 4.54E-08 |
| rs774575 | 6 | 113509950 | A | G | 0.03782 | 0.00597 | 2.30E-10 |
| rs10456943 | 6 | 121877965 | C | A | 0.04571 | 0.00826 | 3.15E-08 |
| rs12206651 | 6 | 130422142 | G | A | 0.04417 | 0.00685 | 1.13E-10 |
| rs28568459 | 6 | 135040792 | G | A | -0.0402 | 0.00719 | 2.28E-08 |
| rs9399136 | 6 | 135402339 | C | T | 0.04956 | 0.00662 | 7.14E-14 |
| rs138449099 | 6 | 170519417 | C | A | 0.07339 | 0.01306 | 1.92E-08 |
| rs12535226 | 7 | 55156419 | A | T | -0.0363 | 0.00583 | 4.95E-10 |
| rs11562010 | 7 | 116521644 | A | T | -0.0329 | 0.00587 | 2.22E-08 |
| rs4737010 | 8 | 41630447 | A | G | -0.0451 | 0.00694 | 7.99E-11 |
| rs9657608 | 9 | 21956230 | T | C | -0.0644 | 0.00978 | 4.47E-11 |
| rs3731211 | 9 | 21986847 | A | T | -0.0859 | 0.00651 | 9.02E-40 |
| rs12555274 | 9 | 22136440 | C | G | 0.03678 | 0.00674 | 4.84E-08 |
| rs10811664 | 9 | 22142907 | A | G | 0.10068 | 0.008 | 2.61E-36 |
| rs9410343 | 9 | 91400584 | A | G | 0.14433 | 0.01195 | 1.37E-33 |
| rs565296553 | 9 | 136064133 | G | A | 0.04482 | 0.00744 | 1.73E-09 |
| rs2519093 | 9 | 136141870 | T | C | -0.0567 | 0.00754 | 5.55E-14 |
| rs72785044 | 10 | 50309849 | G | A | 0.0531 | 0.00697 | 2.61E-14 |
| rs2782979 | 10 | 115781367 | G | T | -0.0328 | 0.00584 | 1.97E-08 |
| rs4910062 | 11 | 8862188 | C | A | 0.03367 | 0.00585 | 8.71E-09 |
| rs2306363 | 11 | 65405600 | T | G | -0.0424 | 0.00721 | 4.09E-09 |
| rs11224302 | 11 | 100456604 | T | C | 0.10328 | 0.00984 | 9.05E-26 |
| rs7127313 | 11 | 100508897 | T | C | -0.0358 | 0.00613 | 5.15E-09 |
| rs1800057 | 11 | 108143456 | G | C | 0.10058 | 0.01794 | 2.06E-08 |
| rs12277928 | 11 | 119096126 | T | G | 0.04706 | 0.00656 | 7.30E-13 |
| rs8705 | 11 | 128328913 | A | G | 0.05201 | 0.00624 | 7.77E-17 |
| rs887477 | 12 | 6445982 | C | A | 0.03863 | 0.00583 | 3.52E-11 |
| rs11611855 | 12 | 6502260 | T | C | 0.0506 | 0.00591 | 1.04E-17 |
| rs876373 | 12 | 111279341 | C | T | -0.0383 | 0.00596 | 1.38E-10 |
| rs756825 | 12 | 111598202 | T | C | -0.0723 | 0.00883 | 2.56E-16 |
| rs150481160 | 12 | 111761895 | T | C | -0.1071 | 0.01616 | 3.40E-11 |
| rs546459162 | 12 | 111865264 | G | C | -0.1477 | 0.0269 | 4.02E-08 |
| rs3184504 | 12 | 111884608 | C | T | -0.1595 | 0.00583 | 8.309E-165 |
| rs138525463 | 12 | 111897485 | T | C | -0.0831 | 0.014 | 2.87E-09 |
| rs117864178 | 12 | 111944439 | T | C | -0.1359 | 0.01489 | 7.14E-20 |
| rs112867136 | 12 | 112032145 | T | C | 0.14214 | 0.02106 | 1.47E-11 |
| rs79423041 | 12 | 112413025 | C | T | -0.103 | 0.01386 | 1.08E-13 |
| rs77979552 | 12 | 112508217 | A | T | 0.10905 | 0.01686 | 1.01E-10 |
| rs111597699 | 12 | 112545031 | C | T | -0.1178 | 0.01527 | 1.25E-14 |
| rs73207624 | 12 | 112674718 | T | C | 0.08935 | 0.01503 | 2.79E-09 |
| rs61941336 | 12 | 112778443 | C | T | -0.0747 | 0.01356 | 3.53E-08 |
| rs11066320 | 12 | 112906415 | G | A | -0.1235 | 0.0059 | 3.01E-97 |
| rs74672796 | 12 | 113016244 | A | C | -0.0602 | 0.01 | 1.73E-09 |
| rs4767948 | 12 | 113116000 | A | G | -0.0575 | 0.00659 | 2.68E-18 |
| rs7970581 | 12 | 113265248 | G | T | -0.0415 | 0.00672 | 6.67E-10 |
| rs2454702 | 12 | 123209159 | T | C | -0.0412 | 0.00713 | 7.54E-09 |
| rs61963226 | 13 | 41535409 | A | T | 0.04634 | 0.00731 | 2.32E-10 |
| rs112193159 | 13 | 49750380 | C | G | -0.0495 | 0.00892 | 2.83E-08 |
| rs61959991 | 13 | 50018842 | G | T | 0.03413 | 0.00587 | 5.93E-09 |
| rs12874404 | 13 | 108993494 | G | A | 0.09617 | 0.0129 | 9.02E-14 |
| rs230703 | 14 | 65267469 | C | T | -0.0358 | 0.00638 | 2.00E-08 |
| rs28688110 | 14 | 93511785 | T | A | -0.0735 | 0.00928 | 2.27E-15 |
| rs12899025 | 15 | 39297872 | A | G | -0.0493 | 0.0059 | 6.08E-17 |
| rs1899226 | 15 | 39610231 | A | T | -0.0412 | 0.00591 | 3.16E-12 |
| rs112642422 | 16 | 4306785 | T | C | 0.03668 | 0.00632 | 6.62E-09 |
| rs4451969 | 16 | 11383519 | C | T | 0.03742 | 0.00621 | 1.65E-09 |
| rs62033400 | 16 | 53811788 | G | A | 0.0391 | 0.00597 | 5.74E-11 |
| rs4480845 | 17 | 1958609 | C | T | 0.05497 | 0.00607 | 1.34E-19 |
| rs111277136 | 17 | 2129904 | A | C | 0.05915 | 0.01033 | 1.02E-08 |
| rs11658587 | 17 | 4901982 | T | C | 0.10596 | 0.01531 | 4.54E-12 |
| rs4968186 | 17 | 7570930 | G | A | 0.04757 | 0.00733 | 8.42E-11 |
| rs34219250 | 17 | 7630365 | T | C | 0.04157 | 0.00734 | 1.49E-08 |
| rs11652760 | 17 | 16786819 | G | T | 0.05507 | 0.00977 | 1.75E-08 |
| rs2060779 | 17 | 57506092 | A | G | 0.0405 | 0.00701 | 7.61E-09 |
| rs2645479 | 17 | 57920532 | G | A | 0.04058 | 0.00586 | 4.34E-12 |
| rs4789614 | 17 | 71438310 | T | C | -0.0416 | 0.00719 | 7.39E-09 |
| rs12967135 | 18 | 57849023 | A | G | 0.03832 | 0.00688 | 2.58E-08 |
| rs59616136 | 19 | 17252041 | A | G | 0.0835 | 0.01001 | 7.12E-17 |
| rs17533903 | 19 | 17256523 | A | G | -0.0511 | 0.0072 | 1.28E-12 |
| rs6045612 | 20 | 1931001 | T | C | 0.03821 | 0.00663 | 8.23E-09 |
| rs1883932 | 20 | 8609588 | T | A | 0.03735 | 0.00583 | 1.46E-10 |
| rs6059958 | 20 | 30143278 | T | C | -0.0454 | 0.00775 | 4.65E-09 |
| rs35963929 | 20 | 39270992 | A | G | -0.0463 | 0.00594 | 7.08E-15 |
| rs6099642 | 20 | 56051451 | T | G | -0.0346 | 0.00587 | 3.92E-09 |
| rs7285885 | 22 | 30182558 | A | G | -0.0519 | 0.00799 | 8.28E-11 |

**Supplementary Table 13:** Independent genome-wide significant SNPs associated with pancreas volume. Reported columns include chromosome (Chr), SNP ID, base pair position, effect allele, other allele, effect size (beta), standard error (SE), and P-value. All results are based on POP-GWAS analysis in the non-imaging cohort.

| **SNP** | **chr** | **pos** | **effect_**allele | **other_allele** | **beta** | **se** | **pval** |
| --- | --- | --- | --- | --- | --- | --- | --- |
| rs6703250 | 1 | 51418472 | T | C | -0.0466 | 0.0074 | 3.01E-10 |
| rs11589479 | 1 | 155033308 | A | G | -0.0632 | 0.00981 | 1.17E-10 |
| rs2816946 | 1 | 199993451 | T | C | -0.0483 | 0.00882 | 4.19E-08 |
| rs10779605 | 1 | 213828800 | A | G | 0.05896 | 0.00777 | 3.30E-14 |
| rs76405838 | 2 | 239791547 | C | T | -0.0599 | 0.00996 | 1.83E-09 |
| rs1390195 | 3 | 55780816 | T | A | -0.0422 | 0.0074 | 1.17E-08 |
| rs73174306 | 3 | 169194244 | T | A | -0.1005 | 0.01805 | 2.63E-08 |
| rs9384688 | 6 | 109132221 | C | T | -0.0437 | 0.00737 | 3.07E-09 |
| rs112166936 | 6 | 126733140 | C | A | -0.044 | 0.00733 | 1.94E-09 |
| rs35323862 | 6 | 127428232 | T | C | 0.07769 | 0.00852 | 7.83E-20 |
| rs917073 | 7 | 51023664 | A | G | 0.10737 | 0.01758 | 1.02E-09 |
| rs7819154 | 8 | 129558351 | A | C | -0.0679 | 0.00816 | 9.25E-17 |
| rs550057 | 9 | 136146597 | T | C | -0.0676 | 0.00838 | 7.08E-16 |
| rs769009 | 10 | 49390449 | T | G | -0.061 | 0.00745 | 2.55E-16 |
| rs10509540 | 10 | 90023033 | C | T | -0.0549 | 0.00818 | 1.97E-11 |
| rs930005 | 11 | 115917338 | T | C | -0.0417 | 0.0073 | 1.09E-08 |
| rs4768387 | 12 | 42331217 | C | G | -0.0431 | 0.00743 | 6.67E-09 |
| rs2638519 | 12 | 53267299 | G | A | 0.04522 | 0.00803 | 1.80E-08 |
| rs10870473 | 12 | 133153281 | T | A | -0.0444 | 0.00784 | 1.56E-08 |
| rs2504212 | 13 | 28537537 | G | T | -0.0518 | 0.00932 | 2.78E-08 |
| rs56994090 | 14 | 101306447 | C | T | -0.0613 | 0.00739 | 1.20E-16 |
| rs72802342 | 16 | 75234872 | A | C | 0.1312 | 0.01366 | 7.83E-22 |
| rs7405380 | 16 | 88975910 | C | G | -0.0499 | 0.00752 | 3.30E-11 |
| rs72868785 | 18 | 855667 | A | T | 0.06319 | 0.01138 | 2.79E-08 |
| rs922048 | 18 | 56876386 | T | A | 0.04668 | 0.00743 | 3.42E-10 |
| rs504215 | 19 | 49272484 | T | G | 0.0442 | 0.00766 | 7.94E-09 |
| rs6020369 | 20 | 48835972 | C | T | -0.0422 | 0.00731 | 7.76E-09 |
| rs5762813 | 22 | 29203314 | T | C | -0.0562 | 0.0094 | 2.18E-09 |
| rs9330813 | 22 | 46364161 | A | G | -0.0467 | 0.00784 | 2.65E-09 |

**Supplementary Table 14:** Independent genome-wide significant SNPs associated with kidney volume. Reported columns include chromosome (Chr), SNP ID, base pair position, effect allele, other allele, effect size (beta), standard error (SE), and P-value. All results are based on POP-GWAS analysis in the non-imaging cohort.

| **SNP** | **chr** | **pos** | **effect_allele** | **other_allele** | **beta** | **se** | **pval** |
| --- | --- | --- | --- | --- | --- | --- | --- |
| rs606240 | 1 | 10539203 | G | T | -0.1493 | 0.0268 | 2.55E-08 |
| rs719285 | 1 | 163619414 | T | C | 0.03336 | 0.00554 | 1.72E-09 |
| rs1266155 | 1 | 177881424 | A | G | 0.03744 | 0.00679 | 3.46E-08 |
| rs2796243 | 1 | 208029182 | T | C | -0.0316 | 0.00555 | 1.26E-08 |
| rs2033874 | 2 | 12102888 | C | A | 0.05 | 0.00648 | 1.19E-14 |
| rs807624 | 2 | 15782471 | T | G | 0.05397 | 0.00578 | 9.94E-21 |
| rs4567937 | 2 | 18676265 | A | G | -0.0457 | 0.00595 | 1.64E-14 |
| rs12997065 | 2 | 148534818 | G | C | -0.0358 | 0.00606 | 3.71E-09 |
| rs2943636 | 2 | 227087284 | A | G | 0.04034 | 0.00554 | 3.28E-13 |
| rs13083375 | 3 | 12365308 | T | G | -0.0596 | 0.0085 | 2.41E-12 |
| rs2306623 | 3 | 25424929 | C | T | 0.03741 | 0.00587 | 1.85E-10 |
| rs7635601 | 3 | 50044006 | A | T | 0.04791 | 0.00642 | 8.15E-14 |
| rs6440008 | 3 | 141154542 | C | T | 0.03397 | 0.00569 | 2.34E-09 |
| rs6440054 | 3 | 141678347 | T | A | -0.0427 | 0.00774 | 3.62E-08 |
| rs10008637 | 4 | 77414144 | C | T | -0.0437 | 0.00555 | 3.49E-15 |
| rs2955170 | 4 | 106214339 | A | T | -0.0318 | 0.00554 | 9.91E-09 |
| rs375826 | 4 | 124007236 | C | G | -0.0367 | 0.00649 | 1.59E-08 |
| rs13179493 | 5 | 39426307 | C | T | -0.0352 | 0.00609 | 7.55E-09 |
| rs40270 | 5 | 55804552 | C | A | 0.04011 | 0.00659 | 1.18E-09 |
| rs3936511 | 5 | 55860781 | G | A | 0.04631 | 0.00703 | 4.55E-11 |
| rs1187117 | 6 | 34169074 | T | G | -0.044 | 0.00729 | 1.68E-09 |
| rs9394951 | 6 | 43350753 | T | C | -0.0332 | 0.00559 | 2.78E-09 |
| rs744103 | 6 | 43805362 | A | T | -0.055 | 0.00597 | 3.12E-20 |
| rs4715227 | 6 | 51491884 | C | T | 0.03231 | 0.00556 | 6.32E-09 |
| rs7768973 | 6 | 109745325 | A | T | -0.0311 | 0.00563 | 3.28E-08 |
| rs4897160 | 6 | 126223944 | A | G | 0.03459 | 0.00554 | 4.42E-10 |
| rs941457 | 6 | 126292187 | C | G | 0.03436 | 0.00587 | 4.72E-09 |
| rs1490384 | 6 | 126851160 | T | C | 0.03855 | 0.00554 | 3.42E-12 |
| rs13199674 | 6 | 127185489 | A | G | 0.03494 | 0.00558 | 3.87E-10 |
| rs15116801 | 7 | 25642893 | G | C | -0.0407 | 0.00704 | 7.76E-09 |
| rs6461879 | 7 | 25698353 | T | C | 0.04777 | 0.00694 | 5.95E-12 |
| rs700753 | 7 | 46753684 | G | C | -0.0338 | 0.00583 | 6.67E-09 |
| rs17145750 | 7 | 73026378 | T | C | -0.0428 | 0.00754 | 1.36E-08 |
| rs8179 | 7 | 92236164 | C | T | -0.0381 | 0.00682 | 2.24E-08 |
| rs6464114 | 7 | 150539253 | G | A | 0.03193 | 0.00555 | 8.72E-09 |
| rs73728279 | 7 | 151411494 | T | G | -0.0422 | 0.00615 | 6.35E-12 |
| rs288762 | 7 | 155633556 | C | T | -0.0396 | 0.00576 | 5.99E-12 |
| rs1881008 | 7 | 156259409 | A | T | -0.0511 | 0.00792 | 1.16E-10 |
| rs2001433 | 8 | 10903475 | A | T | -0.0332 | 0.00554 | 2.01E-09 |
| rs13267474 | 8 | 23784587 | T | C | -0.0549 | 0.00564 | 2.05E-22 |
| rs34181781 | 9 | 96967504 | T | C | -0.0329 | 0.00596 | 3.35E-08 |
| rs68156080 | 10 | 63781673 | G | A | -0.0367 | 0.00622 | 3.71E-09 |
| rs12263369 | 10 | 94823343 | T | C | 0.03239 | 0.00564 | 9.35E-09 |
| rs10767873 | 11 | 30768678 | T | C | 0.03117 | 0.00556 | 2.01E-08 |
| rs479844 | 11 | 65551957 | G | A | -0.0322 | 0.00557 | 7.38E-09 |
| rs7128942 | 11 | 68387984 | A | G | -0.0422 | 0.0075 | 1.81E-08 |
| rs11056383 | 12 | 15320385 | A | G | 0.05525 | 0.00846 | 6.46E-11 |
| rs1042725 | 12 | 66358347 | T | C | -0.0438 | 0.00554 | 2.54E-15 |
| rs35952505 | 14 | 31775687 | T | C | 0.03141 | 0.00564 | 2.56E-08 |
| rs7145712 | 14 | 39524912 | T | C | 0.03305 | 0.00576 | 9.87E-09 |
| rs3095262 | 16 | 6538425 | T | C | -0.0331 | 0.00599 | 3.28E-08 |
| rs77924615 | 16 | 20392332 | A | G | 0.09645 | 0.007 | 3.42E-43 |
| rs74209810 | 16 | 20402466 | T | C | -0.0555 | 0.00894 | 5.15E-10 |
| rs62033406 | 16 | 53824226 | G | A | 0.0381 | 0.00564 | 1.42E-11 |
| rs8073894 | 17 | 59241155 | C | T | -0.0477 | 0.00705 | 1.33E-11 |
| rs9895661 | 17 | 59456589 | T | C | 0.0552 | 0.00741 | 9.10E-14 |

**Supplementary table 15:** MAGMA Gene-Set Analysis – Abdominal Subcutaneous fat

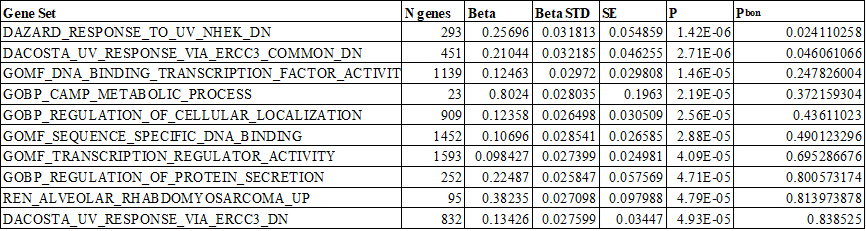

This table shows the top ten Gene-sets obtained using MAGMA Gene-Set analysis from FUMA. The columns are Gene Set: Name of the analysed gene set or GO term, N genes: Number of genes in the gene set included in the analysis, Beta: regression coefficient, Beta STD: Standardized regression coefficient for comparison across gene sets, SE: Standard error of the Beta, P: P-value for the significance of the association, Pbon: Bonferroni-adjusted P-value < 0.05.

**Supplementary table 16:** MAGMA Gene-Set Analysis – Visceral fat

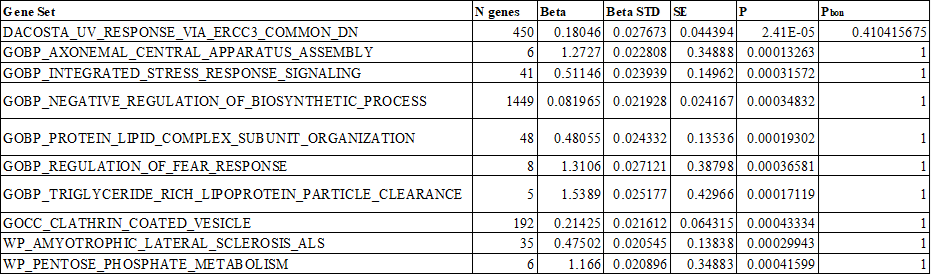

**Supplementary table 17:** MAGMA Gene-Set Analysis – Total muscle volume

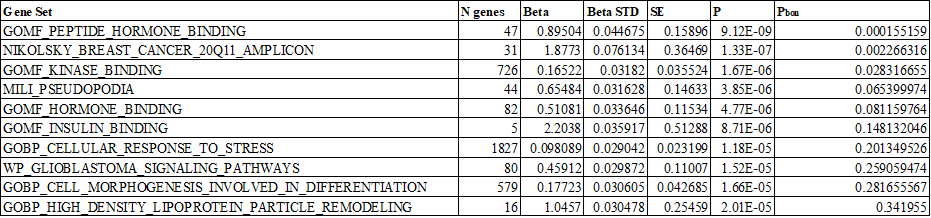

**Supplementary table 18:** MAGMA Gene-Set Analysis – Iliopsoas muscle volume
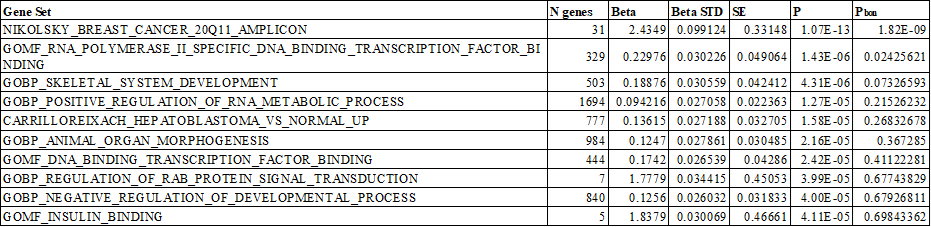

**Supplementary table 19:** MAGMA Gene-Set Analysis – Liver volume

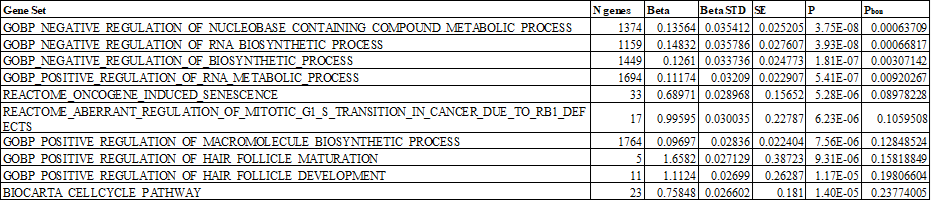

**Supplementary table 20:** MAGMA Gene-Set Analysis – Lung volume

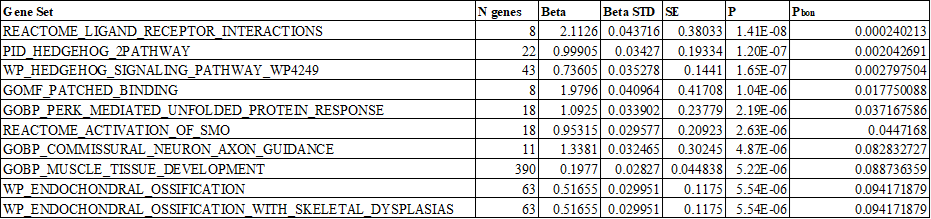

**Supplementary table 21:** MAGMA Gene-Set Analysis – Spleen volume

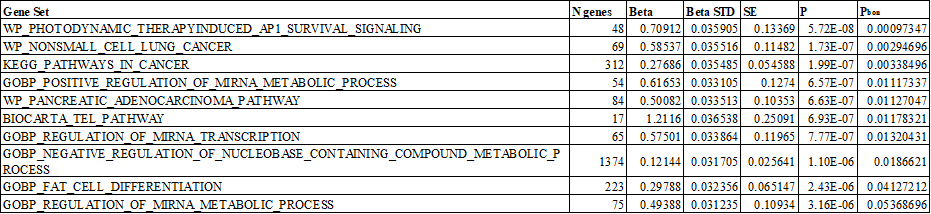

**Supplementary table 22:** MAGMA Gene-Set Analysis – Pancreas volume

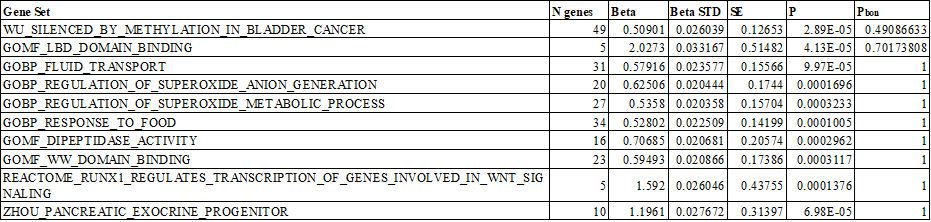

**Supplementary table 23:** MAGMA Gene-Set Analysis – Kidney volume

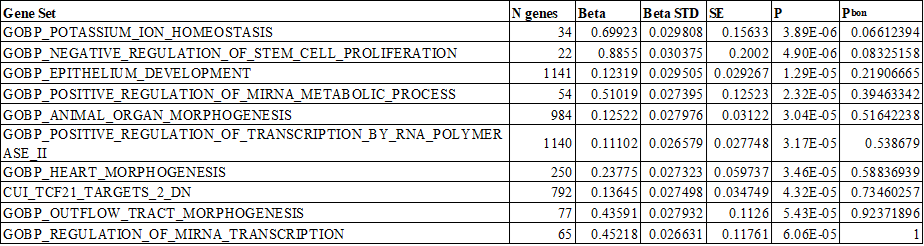

| **Supplementary table 24:** Biomarkers included in genetic correlation analyses: data sources and sample sizes. This table lists the anthropometric, metabolic, and insulin-related biomarkers used in the genetic correlation analyses with image-derived phenotypes (IDPs). For each biomarker, the table provides the corresponding sample size and the original GWAS data source or consortium reference. | | | | | |
| --- | --- | --- | --- | --- | --- |
| **Phenotype** | **PubMed ID** | **Sample size** | **Ethnicity** | **Reference** | **Data source** |
| Whole-body fat-free mass | NA | 454,850 | EUR | Elsworth B. 2018. | ukb-b-13354 |
| Adult BMI, waist/hip ratio (female), waist/hip ratio (male) | 30239722 | 806,834, 379,501, 315,284 | EUR | Pulit SL, et al., Human Molecular Genetics, 2019 | https://zenodo.org/records/1251813 |
| Childhood obesity | 31504550 | 24,160 | EUR | Bradfield JP, et al., Human Molecular Genetics, 2019 | http://egg-consortium.org/childhood-obesity-2019.html |
| Childhood BMI | 33045005 | 39,620 | EUR | Vogelezang S, Bradfield JP, Ahluwalia TS, Curtin JA, Lakka TA, Grarup N, et al., 2020 | http://egg-consortium.org/Childhood-body-mass-index-2020.html |
| BFP | NA | 454,633 | EUR | Elsworth B, 2018 | ukb-b-8909 |
| Birth weight | 31043758 | 298,142 | EUR | Warrington NM, et al., Nature Genetics, 2019 | http://egg-consortium.org/birth-weight-2019.html |
| Adult height | 36224396 | 4,080,687 | EUR | Yengo L, et al., Nature, 2022 | https://portals.broadinstitute.org/collaboration/giant/index.php/GIANT_consortium_data_files#Height_GWAS_Summary_Statistics_without_UK_Biobank |
| Proinsulin levels | 36693378 | 45,861 | EUR | Broadway et al. 2023 | http://magicinvestigators.org/downloads/ |
| Insulin sensitivity index | 37291194 | 55,535 and 55,172 (w/o diabetes) | EUR | Williamson et al. 2023 | http://magicinvestigators.org/downloads/ |
| Disposition index, corrected insulin response, insulin at 30 min, incremental insulin at 30 min | 24699409 | 5,318 | EUR | Prokopenko I, et al., PLoS Genetics, 2014 |  |
| HOMA-B, HOMA-IR | 20081858 | 46,186 | EUR | Dupuis J, et al., Nature Genetics, 2010 | http://magicinvestigators.org/downloads/ |
| HbA1c | 34059833 | 281,416 | EUR | Chen J, et al., Nature Genetics, 2021 | https://magicinvestigators.org/downloads/ |
| Fasting glucose, fasting insulin | 33558525 | 140,595, 98,210 | EUR | Lagou V, et al., Nature Communications, 2021 | https://magicinvestigators.org/downloads/ |
| Sex hormone-binding globulin (female) | NA | 214,989 | EUR | Richmond R., 2020. | ieu-b-4870 |
| Sex hormone-binding globulin (male) | NA | 185,221 | EUR | Richmond, R., 2020 | ieu-b-4871 |
| Leptin | 26833098 | 32,161 | EUR | Kilpeläinen TO, et al., Nature Communications, 2016 | ieu-a-1002 |
| C-reactive protein level | 30388399 | 204,402 | EUR | Ligthart S, AJHG, 2018 | ieu-b-35 |
| Adiponectin | 22479202 | 45,891 (AA *n* = 4,232, EAS *n* = 1,776, EUR *n* = 29,347) | AA, EAS, EUR | Dastani Z, et al., PLoS Genetics, 2012 | ieu-a-1 |
| Cytokines and growth factors | 27989323, 33491305 | 8,293 | EUR | Ahola-Olli AV, et al., AJHG, 2017; Kalaoja, M et al., Obesity, 2021 | https://data.bris.ac.uk/data/dataset/3g3i5smgghp0s2uvm1doflkx9x |
| Liver enzymes (ALP, ALT, GGT) | 33972514 | 437,438, 437,267, 437,194 | EUR | Pazoki R, et al., Nature Communications, 2021 | https://www.ebi.ac.uk/gwas/publications/33972514 |
| Metabolites | 35692035 | 115,078 | EUR | Borges CM, et al., BMC Medicine, 2022. | met-d-* |
| HDL, LDL, and non-HDL cholesterol, total cholesterol, triglycerides | 34887591, 36575460, 35931049 | 1,320,000 | EUR | Graham SE, et al., Nature, 2021; Kanoni S, et al., Genome Biology, 2022; Ramdas S, et al., AJHG, 2022 | https://csg.sph.umich.edu/willer/public/glgc-lipids2021/results/ancestry_specific/ |

| **Supplementary table 25:** Disease outcomes used in Mendelian randomisation analyses: GWAS data sources and sample sizes. This table summarises the published GWAS datasets used to perform Mendelian randomisation (MR) analyses of image-derived phenotypes (IDPs) on disease risk. For each disease outcome, the table provides the full phenotype name, sample size, data source. | | | | | | | | |
| --- | --- | --- | --- | --- | --- | --- | --- | --- |
| **Phenotype** | **n-case** | **n-control** | **sample size** | **year** | **Author** | **PMID** | **Population** | **Data source** |
| Alzheimer disease | 39106 | 46828 | 487511 | 2022 | Bellenguez C | 35379992 | European | ebi-a-GCST90027158 |
| Aortic aneurysm | 3230 | 475964 | 479194 | 2021 | Sakaue S | 34594039 | European | ebi-a-GCST90018783 |
| Asthma | 56167 | 352255 | 408442 | 2021 | Valette K | 34103634 | European | ebi-a-GCST90014325 |
| Atrial fibrillation | 60620 | 970216 | 1030836 | 2018 | Nielsen JB | 30061737 | European | ebi-a-GCST006414 |
| Cholelithiasis | 26122 | 461431 | 487553 | 2021 | Sakaue S | 34594039 | European | ebi-a-GCST90018819 |
| Chronic kidney disease | 41395 | 439303 | 480698 | 2019 | Wuttke M | 31152163 | European | https://ckdgen.imbi.uni-freiburg.de/datasets/Wuttke_2019 |
| Coronary heart disease | 42096 | 361 | 141217 | 2015 | Nikpay M | 26343387 | European | ebi-a-GCST003116 |
| Deep vein thrombosis | 9529 | 475069 | 484598 | 2021 | Dönertaş HM | 33959723 | NA | ebi-a-GCST90038615 |
| Depression | 13559 | 435855 | 449414 | 2021 | Sakaue S | 34594039 | European | ebi-a-GCST90018833 |
| Gastroesophageal reflux disease | 129080 | 473524 | 602604 | 2021 | Ong JS | 34187846 | European | ebi-a-GCST90000514 |
| Gout | 2115 | 67259 | 69374 | 2013 | Kottgen | 23263486 | European | ieu-a-1054 |
| Heart failure | 47309 | 930014 | 977323 | 2020 | Shah S | 31919418 | European | ebi-a-GCST009541 |
| Hip osteoarthritis | 15704 | 378169 | 393873 | 2019 | Tachmazidou I | 30664745 | European | ebi-a-GCST007091 |
| Hypertension | 129909 | 354689 | 484598 | 2021 | Dönertaş HM | 33959723 | NA | ebi-a-GCST90038604 |
| Knee osteoarthritis | 24955 | 378169 | 403124 | 2019 | Tachmazidou I | 30664745 | European | ebi-a-GCST007090 |
| MASLD | 8434 | 770180 | 778614 | 2021 | Ghodsian N | 34841290 | European | ebi-a-GCST90091033 |
| Myocardial infarction | 14825 | 2680 | 395795 | 2021 | Hartiala JA | 33532862 | European | ebi-a-GCST011364 |
| Osteoporosis | 7751 | 476847 | 484598 | 2021 | Dönertaş HM | 33959723 | NA | ebi-a-GCST90038656 |
| Parkinson's disease | 33674 | 449056 | 482730 | 2019 | Nalls MA | NA | European | ieu-b-7 |
| Peripheral artery disease | 7114 | 475964 | 483078 | 2021 | Sakaue S | 34594039 | European | ebi-a-GCST90018890 |
| Polycystic ovary syndrome | 797 | 140558 | 141355 | 2021 | Tyrmi JS | 34791234 | European | ebi-a-GCST90044902 |
| Psoriasis | 15967 | 28194 | 44161 | 2021 | Stuart PE | 34927100 | European | ebi-a-GCST90019017 |
| Pulmonary embolism | NA | NA | 407746 | 2021 | Mbatchou J | 34017140 | European | ebi-a-GCST90013937 |
| Rheumatoid arthritis | 14361 | 43923 | 58284 | 2020 | Ha E | 33310728 | European | ebi-a-GCST90013534 |
| Stroke | 34217 | 406111 | 440328 | 2018 | Malik R | 29531354 | European | ebi-a-GCST006908 |
| Type 2 diabetes | 180834 | 1159055 | 1339889 | 2022 | Mahajan | 35551307 | European | https://diagram-consortium.org/downloads.html |

| **Supplementary table 26:** FinnGen disease outcomes used in Mendelian randomisation analyses: sample sizes and data source. This table lists the disease | | | | |
| --- | --- | --- | --- | --- |
| **Phenotype** | **n-case** | **n-control** | **Sample size** | **Data source** |
| Alzheimer disease | 10520 | 401661 | 412181 | finngen_R10_G6_ALZHEIMER.gz |
| Aortic aneurysm | 8125 | 381977 | 390102 | finngen_R10_I9_AORTANEUR.gz |
| Asthma | 37760 | 219734 | 257494 | finngen_R10_J10_ASTHMA_MAIN_EXMORE.gz |
| Atrial fibrillation | 50743 | 210652 | 261395 | finngen_R10_I9_AF.gz |
| Cholelithiasis | 40191 | 361641 | 401832 | finngen_R10_K11_CHOLELITH.gz |
| Chronic kidney disease | 10039 | 396706 | 406745 | finngen_R10_N14_CHRONKIDNEYDIS.gz |
| Coronary heart disease | 16243 | 381977 | 398220 | finngen_R10_I9_ATHSCLE.gz |
| Deep vein thrombosis | 6501 | 357111 | 363612 | finngen_R10_I9_PHLETHROMBDVTLOW.gz |
| Depression | 54733 | 242809 | 297542 | finngen_R10_F5_DEPRESSION_DYSTHYMIA.gz |
| Gastroesophageal reflux disease | 28859 | 350064 | 378923 | finngen_R10_K11_REFLUX.gz |
| Gout | 9568 | 262844 | 272412 | finngen_R10_M13_GOUT.gz |
| Heart failure | 29672 | 382509 | 412181 | finngen_R10_I9_HEARTFAIL.gz |
| Hip osteoarthritis | 24255 | 262844 | 287099 | finngen_R10_M13_ARTHTROSIS_COX.gz |
| Hypertension | 122996 | 289117 | 412113 | finngen_R10_I9_HYPTENS.gz |
| Knee osteoarthritis | 48836 | 262844 | 311680 | finngen_R10_M13_ARTHROSIS_KNEE.gz |
| MASLD | 2568 | 409613 | 412181 | finngen_R10_NAFLD.gz |
| Myocardial infarction | 26060 | 343079 | 369139 | finngen_R10_I9_MI_STRICT.gz |
| Osteoporosis | 8017 | 391037 | 399054 | finngen_R10_M13_OSTEOPOROSIS.gz |
| Parkinson's disease | 4681 | 407500 | 412181 | finngen_R10_G6_PARKINSON.gz |
| Peripheral artery disease | 11924 | 288638 | 300562 | finngen_R7_I9_PAD.gz |
| Polycystic ovary syndrome | 1639 | 218970 | 220609 | finngen_R10_E4_PCOS.gz |
| Psoriasis | 10312 | 397564 | 407876 | finngen_R10_L12_PSORIASIS.gz |
| Pulmonary embolism | 10046 | 401128 | 411174 | finngen_R10_I9_PULMEMB.gz |
| Rheumatoid arthritis | 13621 | 262844 | 276465 | finngen_R10_M13_RHEUMA.gz |
| Stroke | 18720 | 281378 | 300098 | finngen_R7_I9_STR.gz |
| Type 2 diabetes | 65085 | 335112 | 400197 | finngen_R10_T2D.gz |
